# LLM-based reconstruction of longitudinal clinical trajectories in chronic liver disease

**DOI:** 10.64898/2026.02.10.26345124

**Authors:** Hania Paverd, Zeyu Gao, Golnar Mahani, Margarete A. Fabre, Sarah Burge, Matthew Hoare, Mireia Crispin-Ortuzar

**Author notes:** Contributing authors. These authors contributed equally to this work (Matthew Hoare, Mireia Crispin-Ortuzar).

## Abstract

Liver cancer primarily develops in patients with chronic liver disease (CLD), yet most cases are diagnosed at advanced stages with poor prognosis. While CLD surveillance generates extensive longitudinal data, its free-text nature hinders large-scale research. To address this, we developed a scalable framework using open-source LLMs with constrained decoding to process unstructured text across radiology, pathology, and transplant assessment domains. A calibration set comprising 507 reports from 30 patients was manually annotated to benchmark four LLMs against a regular expression baseline across 70 tasks. Llama-3.3-70B performed best, exceeding 90% accuracy on 59/70 tasks, outperforming Llama-3.1-8B (a smaller variant), OpenBioLLM-70B (a medically fine-tuned model), and DeepSeek-R1-8B. Constrained decoding achieved >99.9% format adherence, far surpassing unconstrained prompting (87.4%). Applied to the full cohort, the pipeline analysed 22,493 reports to generate a patient-level database of 29,225 datapoints (35 variables, 835 patients) without manual annotation. Further analysis confirmed known liver cancer risk factors (male sex, viral hepatitis, smoking, diabetes), and allowed for reconstruction of individualised disease timelines. This work provides a scalable blueprint for transforming real-world clinical free-text into structured formats and personalised patient trajectories. Future applications have the potential to accelerate data-driven research into early cancer detection within complex pre-cancerous diseases like CLD.

## Introduction

Chronic liver disease (CLD) is an increasing global health problem, affecting up to a third of the adult population worldwide [1]. CLD is the main driver of liver cancer (specifically, hepatocellular carcinoma), the third largest cause of cancer death globally [2].

Currently, CLD is most often diagnosed at an advanced stage, when transplantation is the only curative option [3]. Patients with recognised CLD undergo complex multi-disciplinary management with serial clinical investigations (including radiological imaging) performed throughout the disease course [4]. Investigations are performed to ensure recipients are fit enough for transplantation, but also to survey for worsening liver function or incident hepatocellular carcinoma (HCC) development. The diagnosis of HCC is based on imaging [5], interpreted according to the Liver Imaging Reporting and Data System (LIRADS) system [6], but tracking several lesions over time and through rounds of locoregional treatment can be complex.

Curation of longitudinal patient records encompassing multiple source modalities could in the future yield crucial clues for identifying predictive markers of disease progression and HCC development in CLD (Figure 1a). However, this data is mostly stored in unstructured formats - typically as free-text reports - and needs to be converted into tabular formats before any quantitative analysis can be performed. Manual curation of such data is time-consuming and prone to human error, limiting its scalability and routine use. This hurdle could be overcome with automated data extraction methods.

**Fig. 1:**
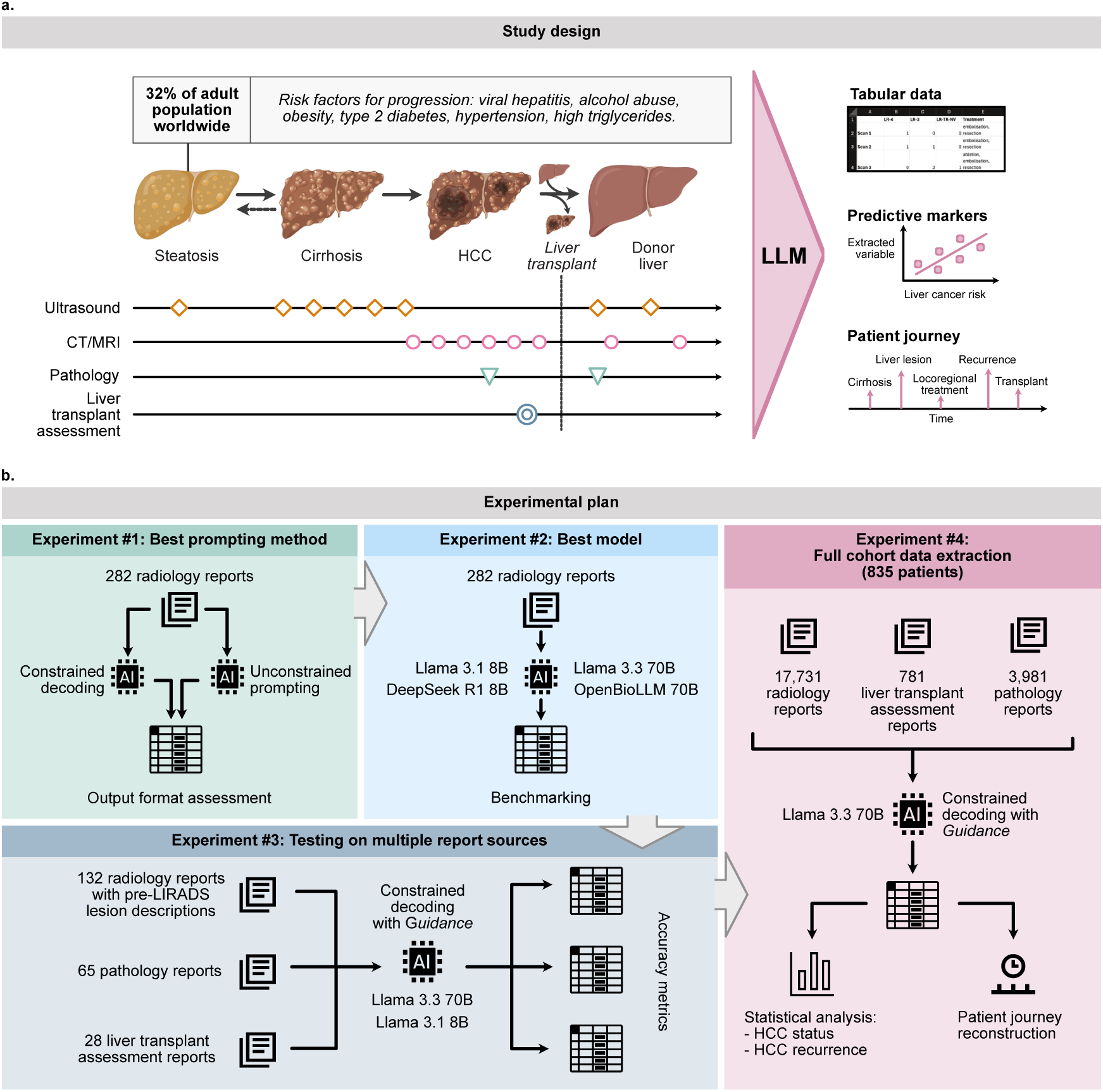
Study design and experimental workflow. **a.** Patients with chronic liver disease undergo complex, multi-modal assessment, including radiology, pathology, and transplant evaluation. In this paper, we validate LLMs performance for turning unstructured reports from these domains into structured tabular data, enabling reconstruction of patient timelines and identification of liver cancer correlates. This approach could facilitate population-scale retrospective research, biomarker development, and time-series modelling to better understand progression from steatosis to cirrhosis and hepatocellular carcinoma (HCC). **b.** Overview of the experimental workflow. In **Experiment 1**, we compared constrained decoding and unconstrained prompting, assessing the format of the outputs against pre-defined format requirements. In **Experiment 2**, we explored four open-source LLMs, assessing their performance against manually-curated ground truth in a set of radiology reports. In **Experiment 3**, we tested the two best-performing models on older radiology reports (before LIRADS system was implemented) as well as pathology reports and liver transplant assessment reports, assessing performance against manually curated ground truth. Finally, in **Experiment 4** we applied the best-performing model to the full cohort of liver transplant patients from the past decade in our institutions, analysing their radiology reports, pathology reports, and liver transplant assessment reports, and performed quantitative analysis of the extracted data.

Large Language Models (LLMs) have emerged as powerful tools for natural language processing and present an opportunity for large-scale automated data curation of complex data such as electronic healthcare records (EHR). LLMs have been successfully applied to extract tabular data from radiology reports [7, 8], pathology reports [9, 10], and clinical notes [11, 12], as well as to convert free-text radiology reports into structured formats [13–23].

Despite these promising results, several key aspects of LLM-based EHR data extraction remain underexplored. First, most studies focus on a single report modality [7, 9–25], overlooking the potential of multi-source records to provide complementary information. Second, studies often benchmark a single LLM [7, 8, 10–13, 15, 17–19, 23, 24], and only few compare LLMs against traditional NLP approaches [9, 11, 25]. Third, the majority of work relies on proprietary models — primarily those from the OpenAI GPT family [7, 9–11, 13–22] — rather than open-source alternatives such as Llama [23, 25] or Mistral [12, 24]. This can have implications to transparency, applicability to confidential datasets, and compatibility with downstream software.

Technical challenges also remain. One major concern is the tendency of LLMs to ”hallucinate” — i.e. to generate incorrect information not present in the input data [26]. Another concern is the lack of specialised medical knowledge in general-purpose LLMs [9]. While medical fine-tuning — training LLMs on curated clinical data — has been proposed to address this issue [27], evidence for its effectiveness remains mixed [28, 29].

Moreover, ensuring correct output formatting remains a practical hurdle, especially when extracting multiple fields simultaneously, as is done in most studies [7–13, 15–25]. Constrained decoding techniques have been proposed to enforce structured outputs to LLMs [30]. Although this approach has shown promise in generating structured radiology reports [23], it has yet to be widely applied to tabular data extraction from multi-source real-world patient datasets.

Finally, most existing research focuses on extracting data from a single time point [7–23], neglecting the longitudinal nature of patient records, which is critical for identifying early prognostic markers and analysing disease progression [24, 25].

In this paper, we investigate multiple open-source LLMs and constrained decoding methods on pre-operative radiology reports of liver transplant patients, extracting not only tumour-specific but also disease-specific information and general clinical markers. We then test the best-performing model and decoding method on other report sources: pathology reports and liver transplant assessment reports. Finally, we perform large-scale data extraction for all the patients transplanted in our institution over the past 10 years (835 patients) and perform quantitative analysis on the extracted tabular data, including pre-transplant timeline reconstruction (Figure 1b).

## Results

### Constrained decoding vs unconstrained prompting

We present a comprehensive evaluation of LLMs in tabular data extraction tasks, spanning technical implementation of output constraints, comparative performance of state-of-the-art LLMs, and large-scale application to real-world multi-source reports.

First, we tackle the challenge of ensuring correct output format by evaluating constrained decoding methods. We compared two software implementations of constrained decoding - one with the *Guidance* package, and one with *Ollama* package - against unconstrained template prompting, where multiple outputs are requested from the model with formatting instructions embedded in the prompt (Figure 2a). In contrast, constrained decoding methods use a programming language syntax to specify the output format which is then enforced onto the LLM output (Figure 2b).

**Fig. 2:**
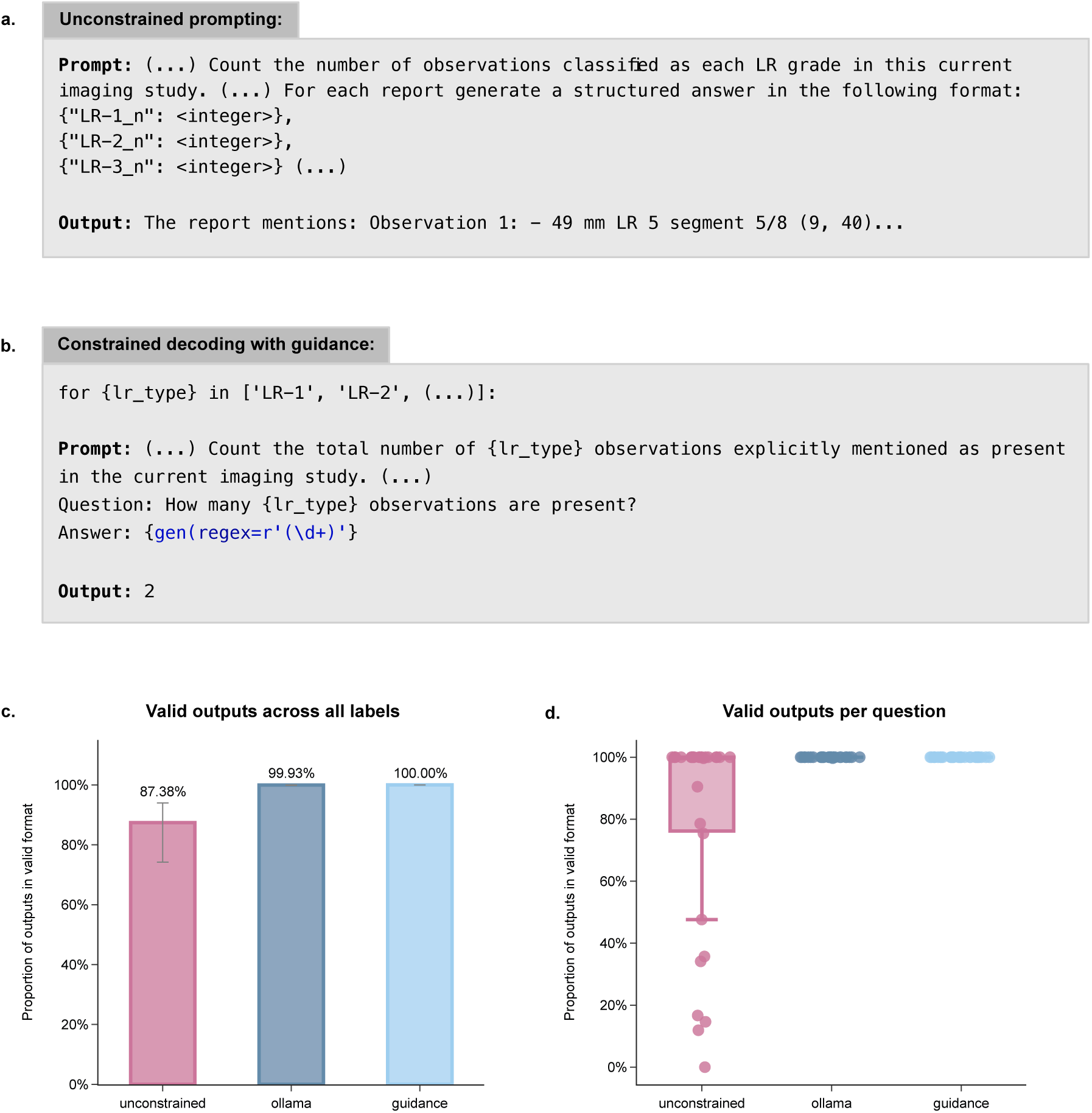
Assessment of constrained decoding. **a** Example of the unconstrained template prompt and LLM output. **b** Example of a prompt using constrained decoding with *Guidance* package (the package syntax is highlighted in blue). **c** Proportion of valid output formats across 5614 labels, shown with bootstrapped 95% confidence intervals (CI95%), for each of the three prompting methods. Format adherence for Unconstrained = 87.38% (CI95% 74.23-93.98), Ollama = 99.93% (CI95% 99.83-100), and Guidance = 100% (CI95% 100-100). **d** Proportion of valid output formats in each of the 31 prompts. LR-1, LR-2, LR-3, LR-5: grades of liver lesions assigned by the radiologists.

We assessed these methods by applying them to radiology reports extracted for a set of 20 liver transplant recipients from our institution (Calibration Set 1). This comprised 282 radiology reports, including 126 liver CT and MRI reports. We developed a set of 31 questions (Supplementary Table 1) with both categorical (e.g. binary yes/no, or multi-class categories) and numeric (e.g. a natural number, or a date) types, resulting in the total of 5614 question-answer pairs. LLMs outputs were assessed for format adherence against pre-defined format requirements (e.g. a natural number, or one of the specified categories). The same model (Llama 3.1 8B) was used across all three prompting methods.

Constrained decoding implementations achieved perfect format adherence with *Guidance* (0% invalid outputs) and near-perfect with *Ollama* (4/5614 invalid outputs, 0.07%, Figure 2c). Unconstrained prompting approach resulted in a 12.61% failure rate (708/5614 invalid outputs) due to format violations. The failure rate was higher among the numeric questions (230/1495 invalid outputs, 15.38%) compared to categorical questions (478/4119 outputs, 11.60%).

The failure rate was not evenly distributed across the 31 questions: seven questions demonstrated valid output rates below 50%, and further three between 50% and 100% (Figure 2d). In constrained decoding with *Guidance*, all questions achieved 100% valid outputs, whereas with *Ollama*, one question achieved 99.6% valid outputs, whilst the remaining 30 questions achieved 100% valid outputs.

To assess the computational cost of constrained decoding, we compared the latency of the extraction methods across Calibration Set 1. On identical hardware (a single NVIDIA A100-SXM4 80 GB GPU), unconstrained template prompting completed the extraction in 73 minutes. In contrast, constrained decoding via the Guidance package required 126.8 minutes - an approximate 1.7-fold increase in execution time. The Ollama implementation, evaluated separately on an Apple M3 Pro chip due to institutional cluster deployment constraints, completed the task in 183.4 minutes.

These results demonstrate that constrained decoding methods, albeit with increased latency, can reliably enforce output structure across diverse clinical question types - an important consideration for integrating LLMs into both research and clinical workflows that rely on consistent, analysable data.

### Benchmarking LLMs in radiology reports

Next, we explored the performance of multiple LLMs and regular expression search (regex) to curate data from retrospective, free-text radiology reports.

We used the same Calibration Set 1 as in the previous experiment, comprising 20 patients with 282 radiology reports, including 126 liver CT and MRI reports, and the same set of 31 questions (Supplementary Table 1). We tested the performance of four open-source LLMs: Llama 3.1 8B, Llama 3.3 70B, OpenBioLLM 70B, and DeepSeek R1 8B, in combination with constrained decoding with *Guidance* and/or *Ollama* packages, depending on the software compatibility. For comparison, regular expressions were developed to detect relevant phrases for each question and to generate answers using simple rule-based logic.

The performance of each model and regex was assessed against the manually curated ground truth with balanced accuracy for categorical questions (Figure 3a), and proportion of correct outputs for numeric questions (Figure 3b), with significance assessed using bootstrapped 95% confidence intervals (CI95%). Full results are provided in Supplementary Tables 2–9.

**Fig. 3:**
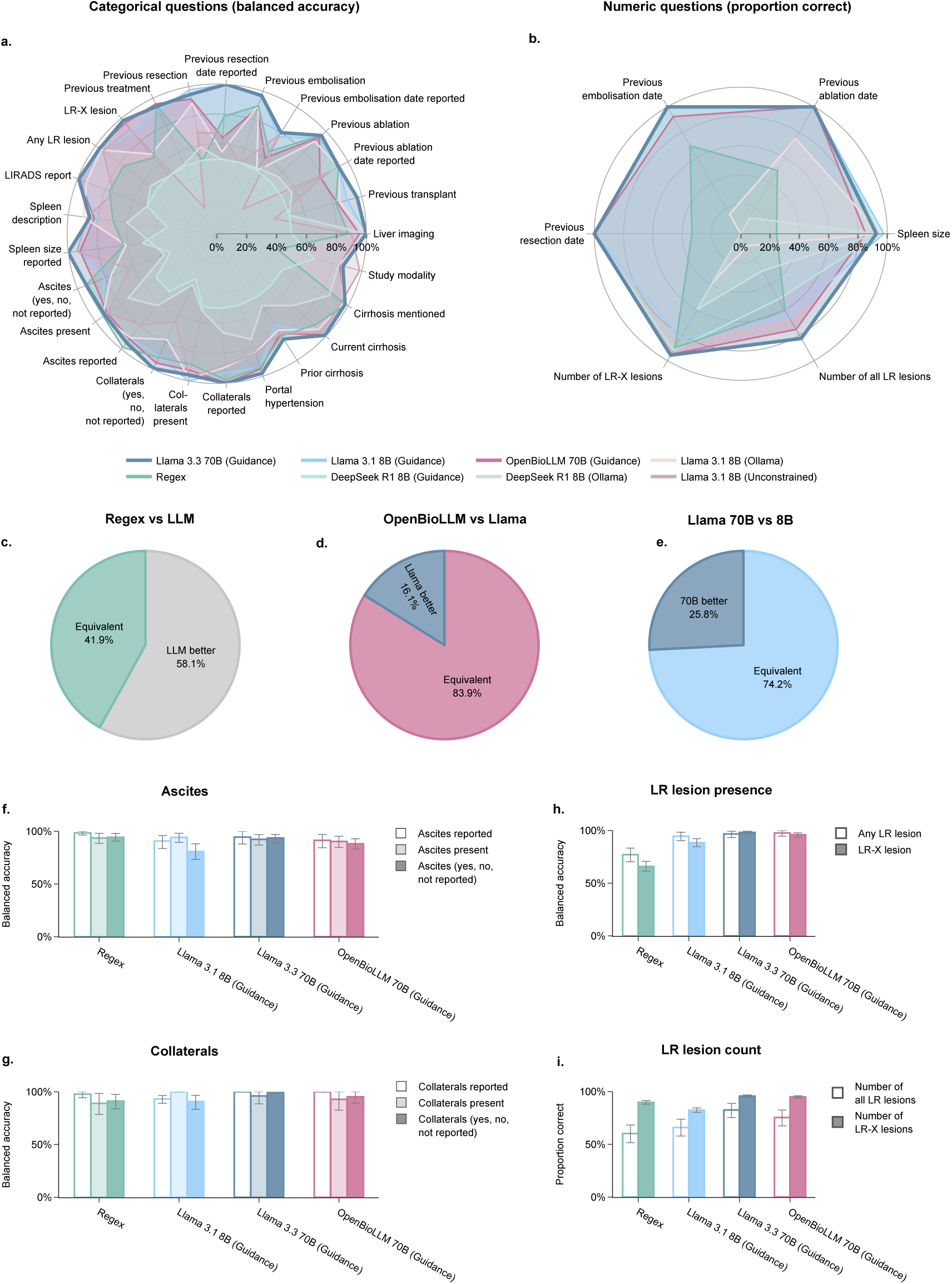
Benchmarking LLMs in radiology reports. **a & b** Performance metrics - balanced accuracy for categorical questions (**a**), and proportion correct outputs for numeric output formats (natural numbers and dates, **b**) - for all tested LLMs and regular expression search (regex). **c-e** Proportion of questions (out of 31 in total) where: **c** performance of regex was at least as good as the best LLM and where at least one LLM was significantly better than regex; **d** OpenBioLLM was at least as good as Llama 3.3 70B and where Llama 3.3 70B was significantly better than OpenBioLLM; **e** Llama 3.1 8B was at least as good as Llama 3.1 70B, and where Llama 3.3 70B was significantly better than Llama 3.1 8B. **f-i** Performance metrics across different question formulations for ascites (**i**), collaterals (**g**), LR lesion presence (**h**), and LR lesion count (**i**).

Across all models tested, DeepSeek R1 8B consistently achieved the lowest balanced accuracy for categorical questions, regardless of whether it was implemented with *Ollama* or *Guidance*. Performance improved for numeric questions when paired with *Guidance*, although the gains were less pronounced with *Ollama*.

An evaluation of extraction accuracy using Llama 3.1 8B revealed that while unconstrained prompting yielded a higher balanced accuracy for identifying study modality, it substantially underperformed on the majority of other tasks, including numeric extractions and previous treatment history.

Upon comparison of regex and LLMs, we found that regex matched or exceeded the top LLM in 13 of 31 questions (41.9%), but was significantly outperformed in the remaining 18 (58.1%; Figure 3c). Notably, regex tended to perform better on categorical questions, while LLMs showed superior performance on numeric questions (Supplementary Figure 1a–b).

We next assessed whether medical fine-tuning conferred any performance benefit by comparing the general-purpose Llama 3.3 70B with OpenBioLLM 70B (a medically fine-tuned model based on Llama 3.1 70B). Llama 3.3 70B significantly outperformed OpenBioLLM in 5 questions (16.1%; Figure 3d) and performed at least as well in the remaining 26 (83.9%). Importantly, OpenBioLLM did not outperform Llama 3.3 70B in any question, and this pattern held across both categorical and numeric questions (Supplementary Figure 1c–d).

Furthermore, we examined the trade-off between performance and model size by comparing Llama 3.1 8B with Llama 3.3 70B. The smaller Llama 3.1 8B was outperformed by the larger Llama 3.3 70B in 8 questions (25.8%; Figure 3e), but performed comparably in the remaining 23 (74.2%), with similar trends observed for both categorical and numeric questions (Supplementary Figure 1e–f).

Overall, Llama 3.3 70B with *Guidance* emerged as the best-performing model, with performance exceeding 90% on 26 of 31 questions, and maintained at ≥80% accuracy across all others. We examined the errors made by Llama 3.3 70B across its worst-performing extraction tasks (Supplementary Table 10). Errors in identifying study modality occurred primarily within complex procedure notes (e.g., misclassifying embolisation reports as standard diagnostic imaging) or during dual-modality pre-MDT reviews. Misidentifications of prior cirrhosis consisted exclusively of false positives; the model detected current cirrhotic features described by the radiologist, but failed to recognise the absence of a documented pre-scan diagnosis. Regarding spleen descriptions, 89% of errors occurred when the model incorrectly extracted it as ”not reported”, often failing to infer a ”normal” status from umbrella phrases like ”normal solid abdominal organs,” despite explicit prompt instructions permitting this deduction. When quantifying LR lesions, 82% of errors were overcounts. These likely stemmed from redundant extractions, where the model double-counted individual lesions referenced across multiple scans summarised within the same report, and from confounding previous LR classification with the currently reported one. Finally, errors in extracting previous embolisation dates were entirely false positives, frequently driven by the model confounding embolisation dates with those of alternative treatments (e.g., ablation) mentioned in the text.

Furthermore, we evaluated whether different question formulations affected LLM performance. For the assessment of reported ascites, we compared a single-question format (with three possible responses: ”yes”, ”no”, or ”not reported”) to a sequential two-step format: first asking whether ascites was mentioned at all, followed by a separate question about its presence (Supplementary Table 24). Llama 3.1 8B showed a trend towards better performance with the two-step formulation, although this was not statistically significant; this trend was not observed for the larger 70B variants (Figure 3f). A similar trend was observed in questions related to the presence of collaterals (Figure 3g). We also explored whether decomposing a question into individual components improved performance in lesion quantification tasks. Specifically, we compared asking about all LR lesion types collectively versus querying each type separately. While binary classification of LR lesion presence yielded comparable performance across both formulations (Figure 3h), lesion counting accuracy was significantly higher when each LR lesion type was queried individually (Figure 3i). Across all tested models, this decomposed approach yielded statistically significant gains: Llama 3.1 8B improved from 65.9% (CI95% 57.9-73.8) to 82.5% (CI95% 80.3-84.5); Llama 3.3 70B from 82.54% (CI95% 75.4-88.9) to 95.7% (CI95% 94.5-96.8); and OpenBioLLM from 75.40% (CI95% 67.5-82.5) to 94.92% (CI95% 93.7-96.0).

Our benchmarking shows that model selection, output constraint mechanisms, and prompt structure all significantly influence extraction performance, with Llama 3.3 70B in combination with *Guidance* emerging as the top-performing configuration, and narrowly focused prompts outperforming broader, multi-element formulations. These observations emphasise the need for deliberate and transparent methodological choices when developing LLM-based systems for clinical data curation.

### Testing on multiple report sources

We evaluated the two best-performing models, Llama 3.3 70B and Llama 3.1 8B, both implemented via *Guidance*, across three distinct clinical document types: radiology reports predating the implementation of the LIRADS system in our centre (i.e., prior to 2019), pathology reports, and liver transplant assessment (LTA) reports.

To assess performance on pre-LIRADS radiology reports, we curated an independent evaluation set (Calibration Set 2) comprising 132 radiology reports from 10 liver transplant recipients, including 68 liver CT and MRI scans. We manually annotated three novel questions addressing the presence, description, and number of liver lesions (Supplementary Table 11). Llama 3.3 70B achieved an accuracy of 96% in identifying lesion presence, 88% in lesion categorisation (“equivocal,” “probably HCC,” and “HCC”), and 95% in lesion count. Llama 3.1 8B performed comparably on the categorical questions but was substantially outperformed on the numeric lesion count task (Figure 4a–c; Supplementary Tables 12-13).

**Fig. 4:**
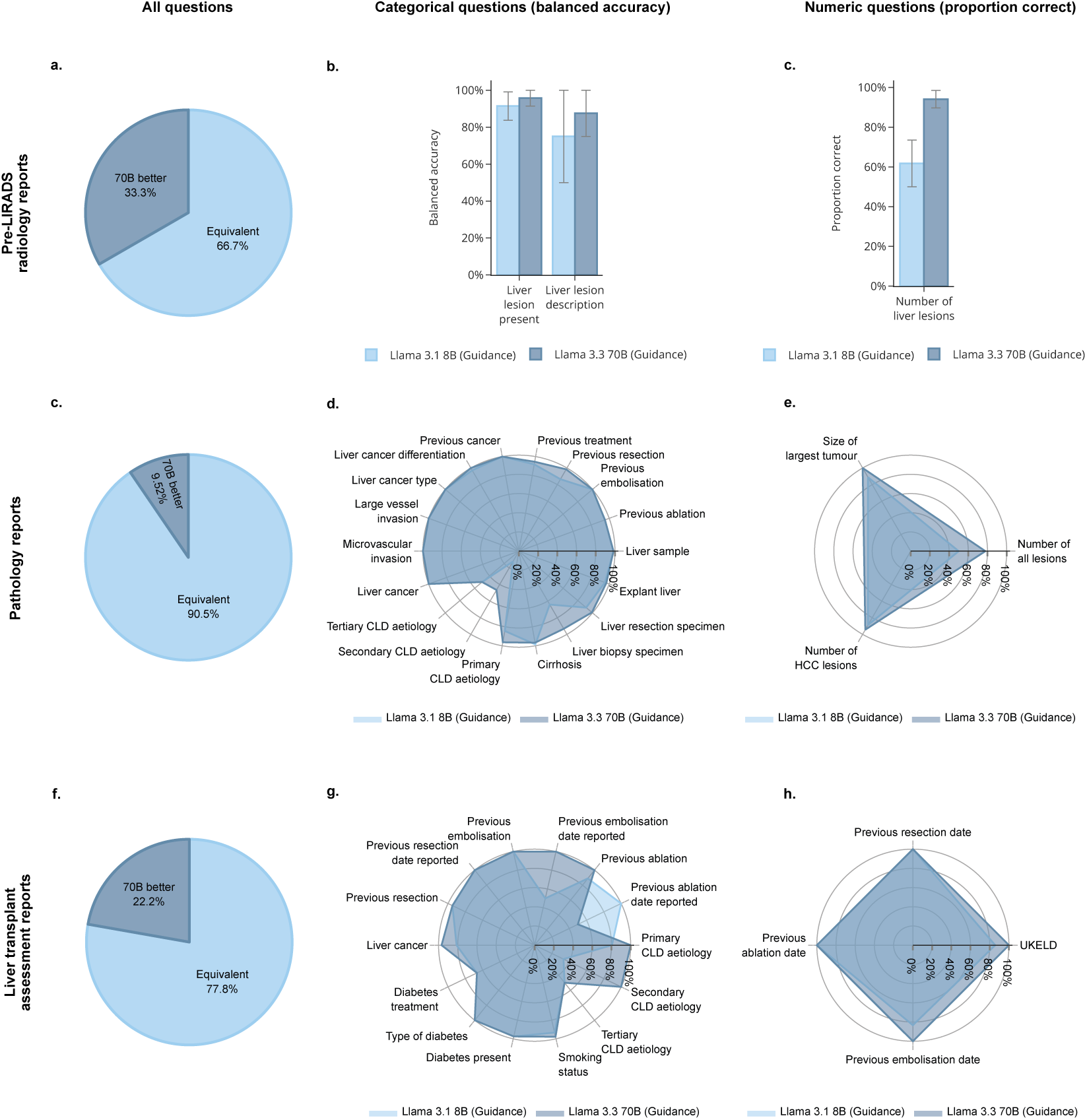
Testing LLMs on pre-LIRADS radiology reports, pathology reports and liver transplant assessment reports. **a** Performance of Llama-70B and Llama-8B for 3 questions across pre-LIRADS radiology reports. **b** Performance of Llama-70B and Llama-8B for 21 questions across pathology reports. **c** Performance of Llama-70B and Llama-8B for 18 questions across liver transplant assessment reports.

Pathology report evaluation was conducted using the combined Calibration Sets 1 and 2 (30 patients; 65 pathology reports). Here, we manually curated 21 questions addressing specimen type, characteristics of underlying liver disease, features of HCC, and treatment history (Supplementary Table 14). The performance of Llama 3.3 70B and Llama 3.1 8B was equally strong in most questions, with significantly lower accuracy of the smaller model observed only in identifying the current specimen as a resection and detecting mention of historical resection (Figures 4c–e; Supplementary Tables 15-16). Llama 3.3 70B achieved >90% accuracy on 18 of 21 questions, with lower accuracy observed on two categorical questions targeting secondary and tertiary CLD aetiologies (47% and 50%, respectively), and one numeric question concerning the total number of lesions (78% correct). Error analysis (Supplementary Table 17) revealed that all errors in lesion counting resulted from the model hallucinating additional lesions; however, the performance improved when the question was formulated to specifically count HCC lesions only (94% correct). For secondary and tertiary CLD aetiologies, all errors involved the LLM missing the respective aetiology (e.g., iron deposition or alpha-1 antitrypsin deficiency) and extracting it as “not reported”.

The evaluation of liver transplant assessment reports was again conducted using the combined Calibration Sets 1 and 2 (30 patients; 28 reports), with 18 manually curated questions addressing demographic data, comorbidities, liver disease and HCC status, and prior interventions (Supplementary Table 18). In this evaluation, Llama 3.3 70B showed superior performance compared to Llama 3.1 8B on four questions, with no statistically significant differences in the remaining 14 questions (Figures 4f–h, Supplementary Tables 19-20). Furthermore, Llama 3.3 achieved >90% accuracy on 15 out of 18 questions, with poorer performance noted in identifying tertiary CLD aetiology (50% accuracy), diabetes treatment method (67%), and whether the date of prior ablation was reported (50%). Error analysis (Supplementary Table 21) showed that the sole error in identifying tertiary CLD aetiology involved missing the diagnosis (similarly to the failure mode observed in pathology reports). The diabetes treatment error occurred when the LLM incorrectly assigned ‘tablet’ despite explicit text stating that metformin has been stopped and the patient was now diet-controlled. Furthermore, the model falsely reported the presence of an ablation date, perhaps confusing it with an embolisation date detailed in the subsequent sentence. However, performance for identifying the actual date of treatment was high (100% for all treatment types).

Overall, these findings demonstrate that Llama 3.3 70B maintains strong performance across heterogeneous clinical documents, suggesting that well-configured LLMs can generalise effectively beyond radiology to broader domains of clinical free-text reports.

### Full-scale data extraction across all report sources

We then applied the best-performing model Llama 3.3 70B, in combination with regex (where its performance was equal to the LLM) to the full cohort of liver transplant patients from our institution between 2014 and 2024, comprising 835 patients with a total of 17,731 pre-operative radiology reports, 3,981 pathology reports and 781 liver transplant assessment reports.

We created a per-patient summary dataframe by aggregating information extracted from multiple reports into a single result (Figure 5a). When multiple records existed per patient, binary variables (e.g., cirrhosis, previous locoregional therapies) were assigned as positive if any of the reports indicated the presence of the feature, whereas ordinal variables (e.g. LR lesion grade, cancer differentiation grade) were summarised by taking the most severe instance of the finding across all reports. We also indicated the source of each variable in the summary dataframe, and added additional sources of information from electronic healthcare records and manual annotations.

**Fig. 5:**
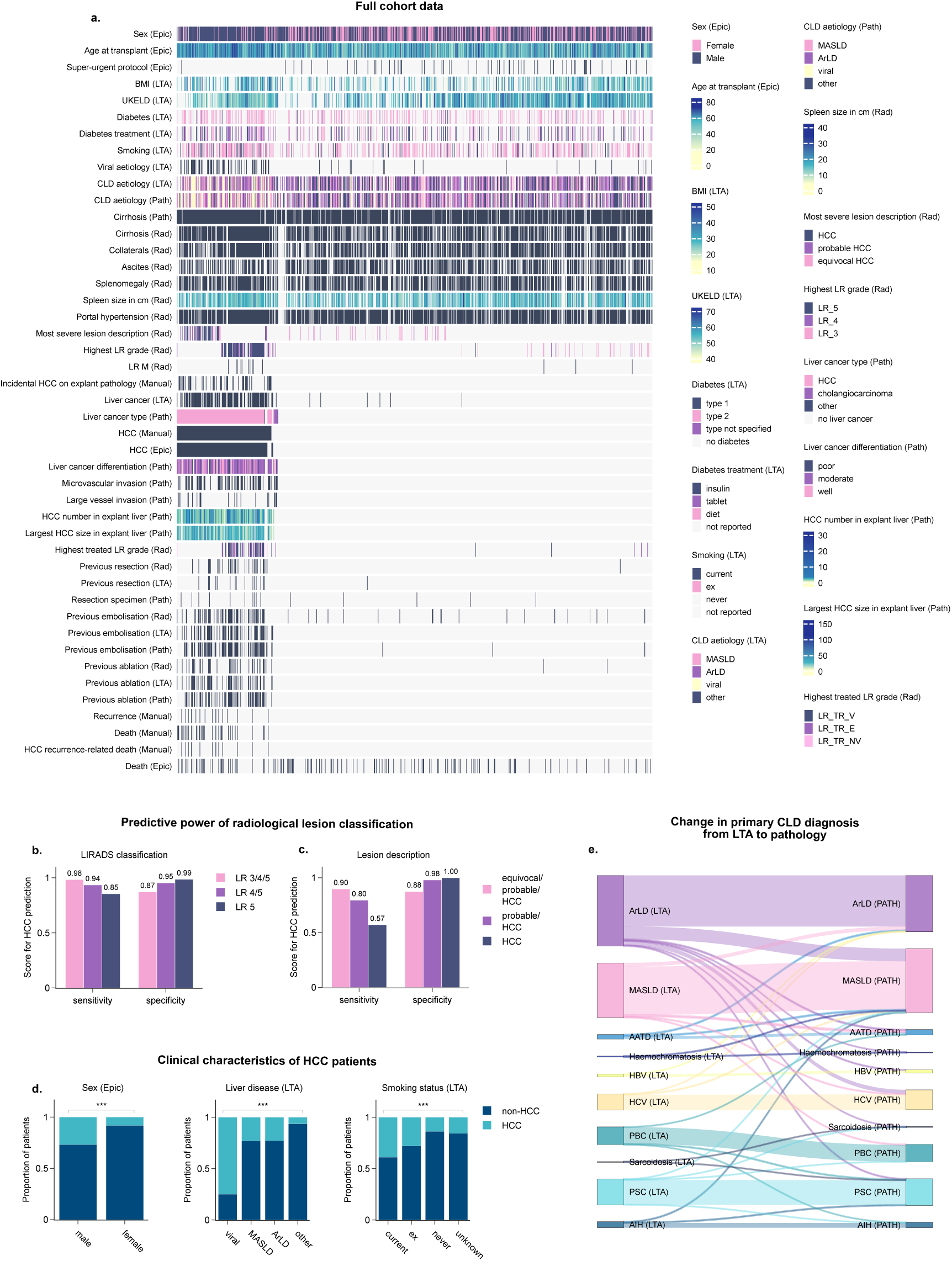
Full-scale data extraction across all report sources. **a** Summary of extracted data for 835 liver transplant patients (shown in columns), across all extracted sources (the source is indicated in brackets for each variable: radiology - ”Rad”, pathology - ”Path”, liver transplant assessment = ”LTA”, electronic healthcare records - ”Epic”, and manual annotations - ”Manual”). **b-c** Sensitivity and specificity of LR lesion classification (**b**) and descriptive lesion classification (**c**), extracted from radiology reports, for predicting pathology-confirmed HCC among patients with liver lesions (incidental HCCs are excluded). **d** Distribution of HCC and non-HCC patients across selected liver transplant assessment variables (the variables have been normalised), assessed for significance with Chi-squared test. **e** Change in primary chronic liver disease (CLD) diagnosis at the time of liver 12 transplant assessment (LTA) and post-transplant at the pathological examination of the explanted liver (PATH). The secondary CLD diagnosis is not shown.

We then analysed the extracted data focusing on several key aspects. First, we assessed the sensitivity and specificity of pre-transplant radiological lesion classification for predicting liver explant pathology-confirmed HCC among patients with liver lesions (incidental HCCs were excluded). Lesions classification as LR-3 or more severe (i.e. LR-3, LR-4, or LR-5) had 0.98 sensitivity and 0.87 specificity for predicting HCC (Figure 5b). As the LR grade increased, so did the specificity for predicting HCC, with LR-5 lesions achieving a near-perfect 0.99 specificity, with 0.87 sensitivity (Figure 5b), matching previously published data [31]. A similar pattern was observed for descriptive lesion classification, with lesions described as ”equivocal”, ”probable HCC” or ”HCC” achieving 0.90 sensitivity and 0.88 specificity. However, for lesions classified as ”HCC” only, the sensitivity dropped to 0.57, with specificity remaining high at 1.00 (Figure 5c).

Furthermore, we investigated the clinical characteristics of patients with and without HCC. We found that patients with HCC were more likely to be male (Chi-Squared test p<0.001), to have liver disease of viral aetiology (p<0.001), and to be smokers (p<0.001) compared to patients without HCC (Figure 5d). Furthermore, there was a higher proportion of diabetics among patients with HCC (p=0.035), and HCC patients had a significantly higher BMI (Mann-Whitney U test p<0.001) and were more advanced in age (p<0.001; Figure 2a).

Among 166 patients with HCC, we found that patients who developed post-transplant HCC recurrence were more likely to have larger lesions (Mann-Whitney U test p=0.039), and to have a poorly differentiated HCC (Chi-Squared test p=0.005) as per the explant pathology report (Figure 2b). HCC recurrence was the only variable significantly associated with mortality within the HCC cohort (Chi-Squared test p=0.001; Figure 3).

Finally, we examined the differences in CLD diagnoses between the liver transplant assessment (LTA) and the pathological examination of the explanted liver post-transplant (PATH; Figure 5e). The majority of diagnostic reclassifications involved patients initially diagnosed with alcohol-related liver disease (ArLD). Specifically, 38 patients (17.8% of those diagnosed with ArLD at LTA) were reclassified as having metabolic-associated steatotic liver disease (MASLD), 14 patients (6.6%) as viral hepatitis, and 9 patients (4.2%) as other aetiologies. In contrast, reclassification from MASLD to other aetiologies was less frequent, with 11 patients reclassified as ArLD (6.7% of those diagnosed with MASLD at LTA), 5 patients (3.0%) reclassified as alpha-1 antitrypsin deficiency and 4 patients (2.4%) reassigned to other aetiologies. Notably, our pipeline also captured several rare, highly divergent diagnostic reclassifications involving primary biliary cholangitis (PBC). To ensure pipeline integrity, we manually audited these cases against the source text (Supplementary Table 22) and confirmed that they accurately reflected the underlying clinical documentation rather than LLM hallucinations. These apparent reclassifications stemmed from several distinct clinical scenarios. In several cases, the discrepancy arose from diagnostic uncertainty at the time of LTA, with reports either explicitly stating the aetiology was unclear or listing multiple differential diagnoses (e.g., raising both MASLD and cholangiopathy as possibilities). In other cases, explant pathology revealed dual aetiologies where a non-PBC condition was deemed predominant, displacing the LTA’s primary diagnosis of PBC in the database. One case contained contradictory documentation within the LTA report itself, where both PSC and PBC were concurrently suggested, likely reflecting a data entry error or historical carryover. Finally, a single case represented a true diagnostic reclassification from MASLD at LTA to PBC on explant pathology, though we note the presence of positive anti-mitochondrial antibodies (AMA) prior to transplant.

Together, these findings demonstrate that LLMs can be used to accurately extract clinically meaningful insights from large patient cohorts spanning diverse sources of real-world clinical data.

### Patient timeline reconstruction

Patients with chronic disease frequently undergo multiple rounds of investigation and treatment that can be difficult to extract from unstructured clinical records. We reconstructed patient pre-transplant timelines focusing on two aspects: LR-reportable liver lesions and previous liver-directed treatments. LR timelines were reconstructed based on radiology reports, whereas the treatment timelines were reconstructed independently from two sources: radiology reports and liver transplant assessment (LTA) reports. Interestingly, we noted that in some cases LTA reports provided more precise information about previous treatment dates, whereas in radiology reports the dates needed to be interpolated from the report dates (Figure 6a).

**Fig. 6:**
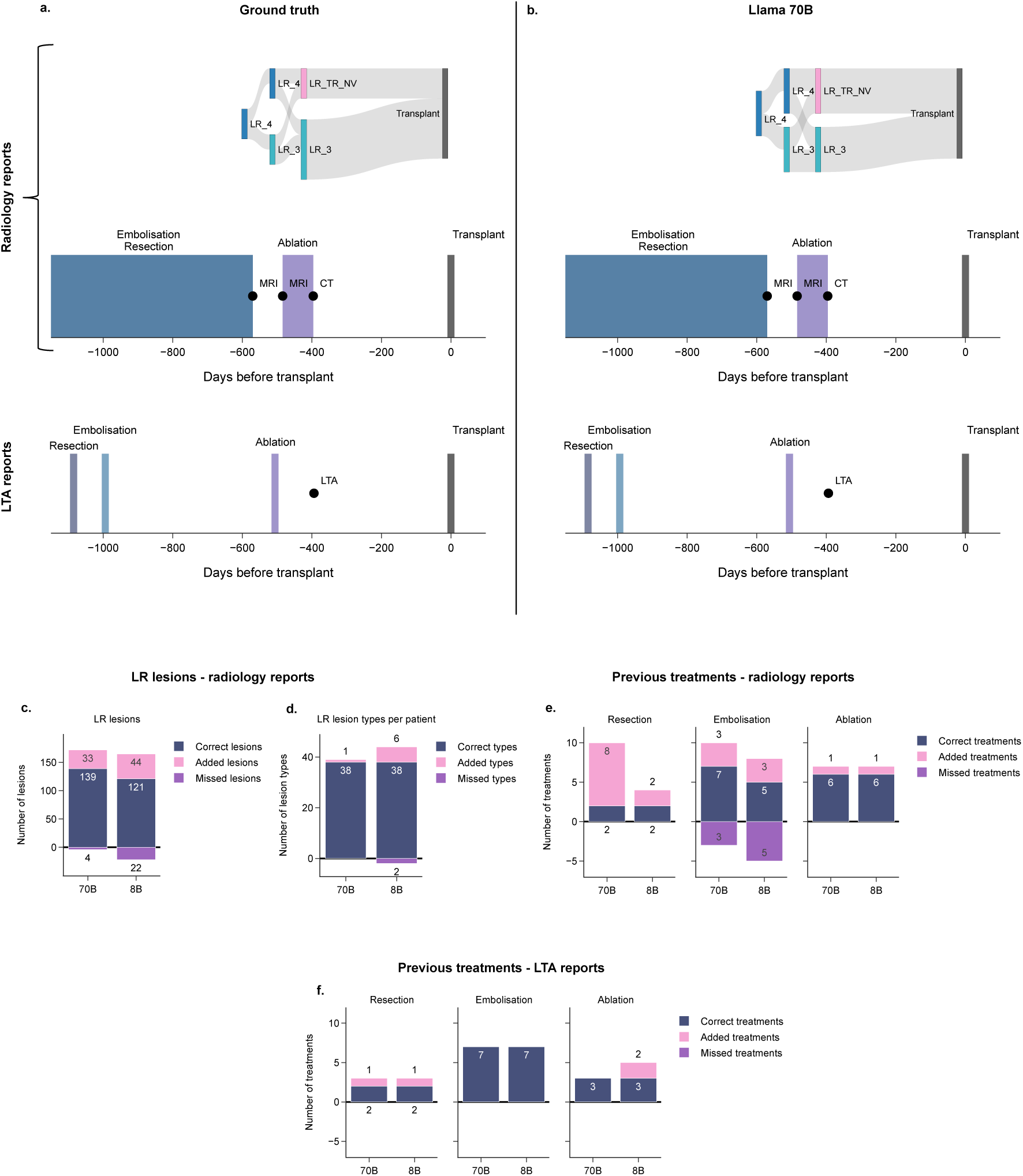
Pre-transplant timelines of patients with HCC. **a-b** Example timelines reconstructed for one patient from ground truth annotations (**a**) and Llama 3.3 70B outputs (**b**). The liver lesion (LR) timeline has been reconstructed from radiology reports data, and the treatment timelines have been reconstructed from radiology and liver transplant assessment (LTA) reports data separately. The flow connections in the LR lesion timelines visually represent aggregated category counts extracted per scan and are intended to show disease burden over time; they do not imply individual lesion-level tracking or linkage across time points. **c-d** Performance of Llama 3.1 8B and Llama 3.3 70B in reconstructing LR lesion timelines, assessed by counting correctly identified LR lesions, missed lesions, and hallucinated (added) lesions (**c**), and by counting the number of new lesion types introduced or missed by the model per patient (**d**). **e-f** Performance of Llama 3.1 8B and Llama 3.3 70B in reconstructing treatment timelines from radiology reports (**e**) and LTA reports (**f** ), assessed by counting correctly identified treatment events, missed events, and hallucinated events.

For evaluation, we reconstructed timelines for 20 patients from Calibration Set 1 (which includes 8 patients with HCC and 12 without) based on ground truth annotations as well as data extracted by Llama 3.1 8B and Llama 3.3 70B (Figure 6b). We assessed the performance by counting instances correctly reconstructed by the model (e.g. correctly identified LR lesions at a specific timepoint, or correctly identified previous treatment and its date), as well as instances which were missed or added (hallucinated).

In the classification of LR lesions, Llama 3.3 70B demonstrated superior performance, achieving sensitivity of 0.97 and precision of 0.81, while Llama 3.1 8B achieved sensitivity of 0.85 and precision of 0.73. This was due to differences in numbers of both correctly identified lesions (139 vs 121 for Llama 3.3 70B vs 3.1 8B) and hallucinated lesions (33 vs 44; Figure 6c). This performance gap was also evident at the individual level among the nine patients with documented LR lesions. Llama 3.3 70B achieved higher patient-level sensitivity (range of 0.86–1.00) and precision (0.75–1.00) compared to Llama 3.1 8B, which showed greater variability (sensitivity: 0.74–1.00; precision: 0.50–1.00). This difference in performance was only partially captured by the aggregated benchmarking metrics across Calibration Set 1: while Llama 3.3 70B showed significantly greater accuracy in distinguishing individual lesion types, both models exhibited similar accuracy in identifying any LR lesion or lesion counts (Figure 3a-b; Supplementary Figure 4). Importantly, while both models hallucinated lesions, Llama 3.3 70B introduced only one new lesion type in a single patient (Figure 6d). In contrast, Llama 3.1 8B added six new lesion types across five patients and failed to detect two lesion types documented in two separate cases (Figure 6d).

To better understand the nature of the errors, we conducted a qualitative review of the failure modes for Llama 3.3 70B (Supplementary Table 23). The model frequently extracted non-existent lesions when a report referenced a previous LI-RADS classification that had since evolved (e.g., erroneously extracting both an LR-4 and an LR-5 lesion from the phrase ”LR-5 (previously LR-4)”), or when a pre-treatment classification was mentioned alongside the post-treatment status (e.g., extracting an LR-5 lesion from ”Pretreatment category: LR-5” for a lesion currently graded as LR-TR-viable). Furthermore, hallucinations occurred when a single clinical note summarised multiple historical imaging studies, leading the model to double-count the same lesion discussed across different timepoints. Conversely, instances of missed lesions occasionally occurred when multiple distinct observations were syntactically grouped into a single descriptive sentence, causing the model to undercount the findings.

In the treatment timeline reconstruction task, Llama 3.3 70B outperformed Llama 3.1 8B in identifying instances of liver-directed embolisation from radiology reports (sensitivity: 0.77 vs. 0.67, respectively, both achieving 0.77 precision), while both models successfully identified all ablation events with one hallucinated instance (1.00 sensitivity, 0.86 precision) (Figure 6e). However, performance was less reliable for resection events; Llama 3.3 70B erroneously added eight resection instances across eight patients, causing precision to drop to 0.20 (with 1.00 sensitivity). In comparison, Llama 3.1 8B introduced only two hallucinated resection events, yielding precision of 0.50 (with 1.00 sensitivity).

Both models demonstrated improved accuracy when extracting treatment histories from LTA reports. In this context, Llama 3.3 70B achieved perfect precision and sensitivity (1.00) for both embolisation and ablation, adding only one spurious resection event (precision: 0.67, sensitivity: 1.00). While Llama 3.1 8B also maintained perfect sensitivity (1.00) across all three modalities, it introduced one resection and two ablation events not documented in the ground truth, resulting in a precision of 0.67 for resection and 0.60 for ablation (Figure 6f).

Our results illustrate that LLMs can be used not only to extract structured data but also to reconstruct temporally resolved patient trajectories - an essential step for enabling longitudinal analyses in chronic disease cohorts.

## Discussion

This study presents, to our knowledge, the first comprehensive evaluation of LLMs for curating and integrating multi-source datasets from real-world clinical documentation in the setting of chronic disease. We demonstrate that an open-source, general-domain LLM can accurately extract clinically relevant information from diverse document types, including radiology, pathology, and transplant assessment reports. Using LLM-derived data, we conducted institution-wide analyses spanning a decade of liver transplant cases, assessing lesion grading practices, uncovering predictors of cancer progression, and tracing reclassification patterns across clinical report types. We also showed that LLMs can support longitudinal patient profiling by reconstructing timelines of lesions and treatments, which is critical for characterising disease progression in chronic conditions.

We implemented several technical solutions to enhance the applicability of LLMs to real-world clinical cohorts. In particular, we demonstrated that constrained decoding successfully enforces correct output formatting, enabling a multi-step prompting approach. This design allows the output of one query to inform subsequent prompts, thereby reducing the risk of spurious or irrelevant generations — such as prompting for tumour size in patients without cancer — a critical consideration when working with heterogeneous clinical populations. Moreover, constrained decoding likely contributed to high performance of Llama models on numeric questions (in contrast to previously reported challenges [7]) by reducing formatting errors demonstrated with unconstrained prompting. However, our latency analysis revealed that this format reliability carries a computational cost, with Guidance requiring approximately 1.7 times the execution time of unconstrained prompting - a significant consideration for scaling these methods to larger datasets or real-time applications. Interestingly, DeepSeek R1 8B significantly underperformed Llama models on categorical questions, with numeric performance improving under *Ollama* but not *Guidance*. We hypothesise that this discrepancy may reflect incompatibilities between reasoning-focused models and the constrained decoding frameworks.

Beyond the choice of decoding framework, the formulation of the prompts themselves played a role in performance. While the benefits of narrowly targeted, step-wise prompts for simple classification tasks were limited to the smaller Llama 3.1 8B model, decomposing the more complex lesion counting task yielded significant improvements across all models. Asking models to count all lesion types collectively resulted in worse performance compared to querying specific lesion types individually, aligning with findings from [24]. We hypothesise that isolating variables in this manner effectively reduces the reasoning burden over unstructured text, making complex extraction tasks more tractable even for highly capable models.

Overall, Llama 3.3 70B emerged as the best-performing model compared to other LLMs, with no clear advantage derived from using a medically fine-tuned model. A possible explanation is a mismatch between the benchmark tasks: medical benchmarks often include multiple-choice questions as opposed to real-world clinical data extraction [32], and recent studies suggest that the benefits diminish when models are evaluated on more diverse set of benchmarks [29]. Alternatively, the observed differences may stem from base-model generation: OpenBioLLM is built on the earlier Llama 3.1 architecture, and the newer Llama 3.3 70B may have benefited from architectural improvements and training on more recent data. Benchmarking models of different sizes revealed that Llama 3.1 8B performed comparably to the 70B model in most tasks, however the larger model had a performance advantage in specific scenarios across all three clinical report types. Interestingly, regular expression search was also comparable with Llama 70B on many categorical questions (contrary to previous studies comparing LLMs to traditional NLP methods [9, 11, 25]), although it lagged behind on numeric questions. Both the smaller LLM and regex offer practical advantages over the larger model: they are faster and more cost-effective, and can be deployed on standard hospital infrastructure without requiring specialised hardware or risk data leakage through cloud access.

The use of LLM-extracted data enabled comprehensive analyses across the entire cohort of 835 liver transplant patients, demonstrating the potential for scalable, institution-wide studies of disease patterns and diagnostic practices. Leveraging this structured dataset, we assessed the sensitivity and specificity of radiological lesion classification for predicting pathology-confirmed HCC, finding diagnostic performance at our center to be on par with published data [31]. We also identified clinical variables associated with HCC development in our cohort — including viral aetiology, male sex, smoking, diabetes, higher BMI, and older age — in line with established literature [33, 34]. Additionally, we examined changes in chronic liver disease diagnoses between transplant assessment and post-transplant pathology, with most reclassifications shifting from ArLD to alternative aetiologies.

This highlights the challenge of diagnostic bias in the presence of overlapping risk factors and the pathological similarity between ArLD and MASLD [35–37].

Finally, we reconstructed longitudinal patient timelines from LLM-extracted data, including the identification of liver lesions and prior treatments. The larger Llama 3.3 70B model outperformed the smaller 8B variant in identifying LR lesions; however, both models struggled to accurately identify prior resections, particularly in patients with HCC who had undergone multiple treatments. These errors likely stem from a known limitation of LLMs: the tendency to hallucinate, particularly when information is implicit or distributed across the text [26]. It is worth noting that aggregate benchmarking metrics indicated high performance for both models - however, this was calculated across all reports, including a high volume of true negatives (e.g., lesion-free scans). Conversely, timeline reconstruction was evaluated at the event level, assessing errors against true-positive events only. This highlights the necessity of employing task-specific, event-level evaluations to accurately gauge a model’s readiness for potential clinical applications. Interestingly, extraction of prior treatment dates was more accurate for liver transplant assessment reports compared to radiology reports, possibly due to their more standardised structure.

Our in-depth error analysis across both benchmarking and timeline tasks revealed that many of extraction failures stemmed from the model’s limited capacity to interpret nuanced context across temporal, syntactic and clinical dimensions. Temporally, it struggled with relative changes (e.g., ”reduction in volume”), discontinued treatments (e.g., diabetes medication), and historical references (e.g., ”previously LR-4”). Syntactically, we observed contextual misattribution, with the model misassigning treatment dates to nearby procedures, or double-counting lesions summarised across multiple scans or report sections. In the clinical dimension, the LLM failed to deduce implicit normalcy (e.g., inferring a normal spleen from ”normal abdominal viscera”). Crucially, the clinical impact varies. Hallucinations (e.g., added lesions) carry severe consequences by improperly upstaging disease severity. Conversely, omissions (e.g., missing minor secondary aetiologies or implicit normalcies) reduce dataset granularity but are less likely to introduce dangerous clinical entries.

Several strategies are directly applicable to clinical data extraction tasks to mitigate hallucinations in future work. Data-level approaches include filtering out low-quality reports and enforcing structured input formats [26], aligning with broader trends toward structured reporting in radiology [38]. Inference-level strategies include self-verification, where the model cross-checks extracted events against the source text or external knowledge bases, and ensemble confirmation methods that reconcile information across multiple report types (e.g., radiology and LTA reports) [26]. Acceptable error thresholds, as well as the required level of data granularity, will necessarily depend on the downstream use of LLM-derived trajectories. For exploratory, cohort-level analyses, patient-level summaries (e.g., aggregate lesion counts) are appropriate and may tolerate minor errors. In contrast, direct clinical applications (e.g., surgical planning) would require near-zero fabrication, as well as much finer temporal and spatial granularity, such as tracking individual lesions across sequential scans. While further research is needed to achieve such granularity, our framework demonstrates LLMs’ potential for longitudinal data curation in chronic diseases — a key step toward incorporating the temporal dimension into patient data analysis and predictive modelling, with the aim of improving understanding of disease progression and treatment outcomes over time.

This study has several important limitations. First, our results reflect a highly tailored approach: a single model (Llama 3.3 70B) employed on a focused single-institution cohort, with prompts designed by a medical domain expert. As the extraction performance will likely vary across different clinical contexts, institutions and tasks, further validation across diverse datasets remains essential to ensure reliability in other settings. Furthermore, all ground truth annotations for the calibration sets were created by a single individual, making it difficult to definitively distinguish between true pipeline errors and annotator subjectivity, particularly when interpreting ambiguous free-text clinical descriptions. Second, although we included patients both with and without HCC, our cohort was still cancer-enriched and liver transplant–specific, which limits the generalisability of our findings. Because transplant selection relies on strict clinical criteria and multidisciplinary judgment, downstream analyses — such as identifying variables associated with HCC development — must be interpreted with caution and may not apply to broader chronic liver disease populations. Furthermore, the higher prevalence of HCC in the calibration datasets compared to the full dataset may have introduced internal bias in our model evaluation metrics due to differences in the underlying population distributions. Third, some of the extraction tasks involved rare events, such as identifying specific treatment dates. These events only occur in a small subset of patients (e.g., those with HCC who were treated and whose treatment date was explicitly documented in the report), which constrained the number of examples available for model evaluation and likely reduced statistical power. This low event frequency is particularly relevant for timeline reconstruction tasks, where the presence, type, and timing of interventions must all be clearly stated in the text to enable accurate extraction. While our study leaves the performance of proprietary models unassessed, our decision to evaluate exclusively open-source models was driven by the direct privacy constraints governing our clinical datasets. Similar constraints are inherent to most real-world clinical environments, where utilising proprietary APIs requires transmitting Protected Health Information to third-party commercial servers. Because data governance frameworks often prohibit this without complex and costly enterprise agreements — which were unavailable at our institution — locally deployed open-source models often remain the most pragmatic and privacy-compliant approach for institution-wide clinical data curation.

Several directions for future work emerge from this study. Expanding the application of LLM-based extraction to other cancer types and disease areas is key, particularly for conditions with complex, longitudinal disease trajectories. Furthermore, broader benchmarking across LLM families is needed, as few studies have systematically compared models from different providers or architectural lineages [21, 39]. In parallel, the development of standardised evaluation datasets and shared benchmarks would help ensure consistency across studies and facilitate reproducibility. While our findings highlight the utility of constrained decoding, future research should also explore the trade-offs between output reliability and the reasoning flexibility required for more complex inferential tasks. Finally, further work on lightweight models that can run securely on local infrastructure without sacrificing performance could support clinical deployment, especially in settings with limited computational resources or strict data privacy requirements.

In summary, this study demonstrates the feasibility and utility of using large language models to extract structured data from real-world, multimodal clinical documentation in a chronic disease context. Through comprehensive evaluation across document types, extraction tasks, and model configurations, we show that open-source LLMs can support scalable analyses of disease risk factors, lesion grading, diagnostic consistency, and longitudinal patient trajectories. Importantly, our findings highlight that performance gains must be weighed against practical deployment considerations, with smaller models and regex-based approaches offering viable, cost-effective alternatives for certain use cases.

Our work serves as a proof-of-concept showcasing that LLMs can enable large-scale data curation without manual annotation, opening the door to institution-wide retrospective studies and timeline reconstruction — approaches that are fundamentally extensible to other chronic disease cohorts and healthcare settings. As LLM capabilities continue to advance, their integration into clinical research pipelines has the potential to transform the scale, speed, and scope of real-world evidence generation.

## Methods

This study was performed under ethical approval of the Health and Social Care Research Ethics Committee A, REC reference 20/NI/0109, IRAS 285521. All the patients agreed to the use of their anonymised data for research purposes by signing a written consent form.

### Datasets

We curated a multi-source dataset of clinical reports from liver transplant patients treated at our institution between 2014 and 2024 (Supplementary Figure 5). The final cohort consisted of 835 patients, with a total of 17,731 radiology reports, 3,981 pathology reports and 781 liver transplant assessment reports (Supplementary Figure 6).

Radiology reports were noted to be of variable structure and detail. Some followed a standardised template for liver imaging, typically including information liver lesions (with LIRADS classification and information on previous treatment, if relevant), chronic liver disease features (e.g. portal hypertension, spleen size), and assessment of biliary and vascular findings (Supplementary Figure 7a). Others were less structured and significantly more abbreviated (Supplementary Figure 7b), including imaging summaries written for multidisciplinary team review, particularly when the primary scan was performed externally and has already been fully reported elsewhere. These reports often focused on key findings without full protocol details or background context. The number of radiology reports per patient ranged from 3 to 107, with a median of 19 (Supplementary Figure 6a).

Pathology reports generally consist of four free-text sections: Clinical Information (clinical details provided by the referrer), Gross Description (e.g. liver size, nodularity, lesion number and location), Microscopic Description (e.g. tumour morphology and grade, background liver changes, vascular invasion), and Final Diagnosis (summary of findings for each specimen). Where hepatocellular carcinoma (HCC) was identified, a structured Synoptic Report is included, providing a standardised summary of key pathological features (e.g. tumour size and number, margins, treatment response, TNM staging, background aetiology; Supplementary Figure 8a). The number of pathology reports per patient ranged from 1 to 52, with a median of 4 (Supplementary Figure 6b).

Liver transplant assessment reports are multidisciplinary documents summarising a patient’s suitability for liver transplantation, incorporating input from hepatology, surgery, anaesthetics, cardiopulmonary medicine, dietetics, and transplant coordination teams. Each report includes routine demographic and clinical metadata (e.g., age, diagnosis, blood group, UKELD score), followed by detailed narrative sections, including liver disease history (including aetiology and any locoregional therapies), medication and social history, clinical examination findings, investigations (including laboratory tests and imaging), specialist reviews, transplant coordinator notes, and a summary and listing decision section (Supplementary Figure 8b). Liver transplant assessment reports were available for 779 patients and missing for 56 patients (Supplementary Figure 6c).

To explore the performance of large language models (LLMs) on tabular data extraction from these reports, we created two calibration sets (Table 1). Calibration Set 1 consisted of 20 patients and included 282 radiology reports (126 liver CT and MRI). To support model calibration, the 20 patients were selected using stratified sampling based on known HCC status (8 with, 12 without), ensuring representation from both earlier and later entries in the cohort. This approach ensured that a sufficient number of ground truth labels were available for evaluating HCC-related questions. Report contents were not reviewed at the time of selection. The number of radiology reports per patient ranged between 5 and 52, with median of 17 (Supplementary Figure 6d). Calibration Set 2 consisted of 10 patients, all of whom underwent transplant prior to the implementation of the LIRADS system in 2019, and included 132 radiology reports (68 liver CT and MRI), ranging between 8 and 51 reports per patient with median of 21. All patients in Calibration Set 2 had known HCC, selected to assess LR lesion information extraction tasks in radiology reports. For multi-source testing, both calibration sets were used, with a total of 65 pathology reports (range between 1 and 7, median of 3 reports per patient; Supplementary Figure 6e) and 28 liver transplant assessment reports (available for 28 patients, missing for 2 patients; Supplementary Figure 6f). The size of the calibration sets was determined pragmatically based on the time required to generate ground-truth annotations, balancing the need for sufficient evaluation examples with study feasibility. Due to the enrichment strategy used to assess HCC-related questions, the calibration dataset contained a higher prevalence of HCC-positive patients compared to the full dataset (Table 1).

**Table 1:**
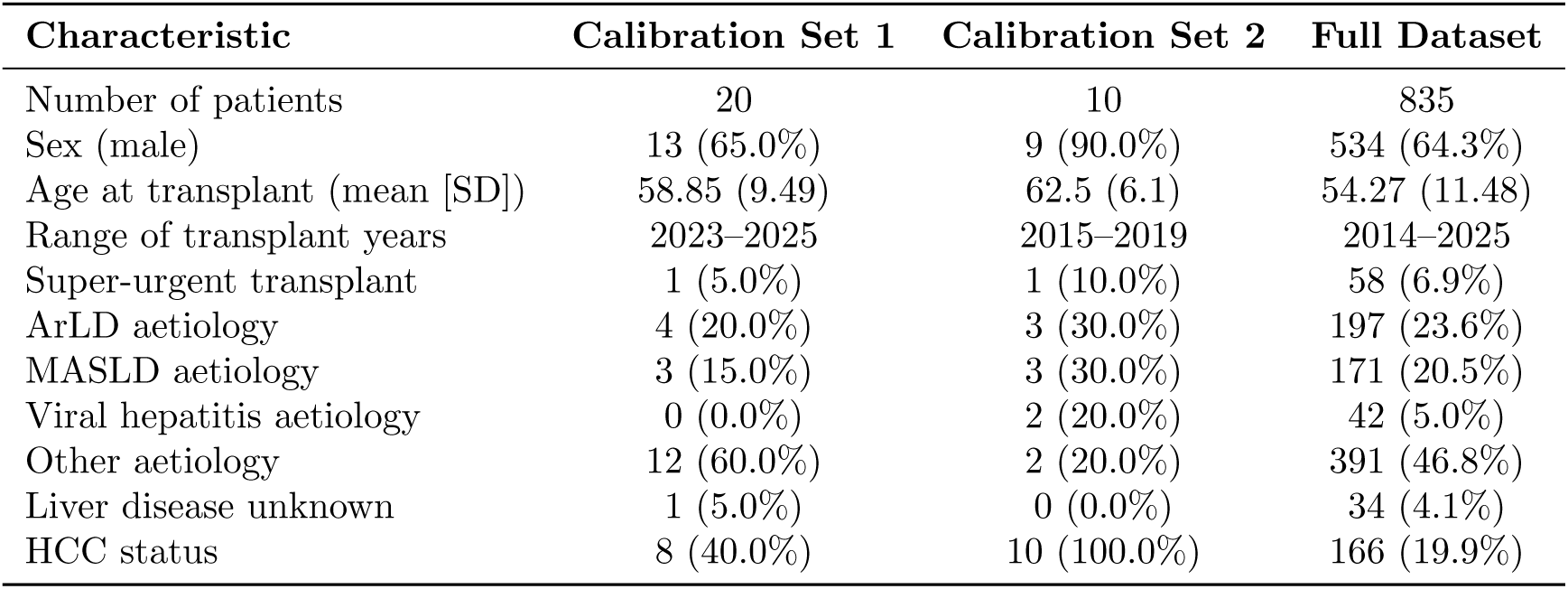
Summary statistics for calibration sets and full dataset.

| Characteristic | Calibration Set 1 | Calibration Set 2 | Full Dataset |
| --- | --- | --- | --- |
| Number of patients | 20 | 10 | 835 |
| Sex (male) | 13 (65.0%) | 9 (90.0%) | 534 (64.3%) |
| Age at transplant (mean [SD]) | 58.85 (9.49) | 62.5 (6.1) | 54.27 (11.48) |
| Range of transplant years | 2023–2025 | 2015–2019 | 2014–2025 |
| Super-urgent transplant | 1 (5.0%) | 1 (10.0%) | 58 (6.9%) |
| ArLD aetiology | 4 (20.0%) | 3 (30.0%) | 197 (23.6%) |
| MASLD aetiology | 3 (15.0%) | 3 (30.0%) | 171 (20.5%) |
| Viral hepatitis aetiology | 0 (0.0%) | 2 (20.0%) | 42 (5.0%) |
| Other aetiology | 12 (60.0%) | 2 (20.0%) | 391 (46.8%) |
| Liver disease unknown | 1 (5.0%) | 0 (0.0%) | 34 (4.1%) |
| HCC status | 8 (40.0%) | 10 (100.0%) | 166 (19.9%) |

Manual ground truth annotations were created for each report type by a resident doctor (HP) with 3-years experience in clinical radiology and 3-years experience in general medicine. For radiology reports, 31 clinically relevant questions were defined across categorical (binary and multi-class) and numeric (natural numbers and dates; Supplementary Table 1). Three additional questions were added for pre-LIRADS radiology reports, covering lesion presence, size, and description (Supplementary Table 11). For pathology reports, we developed a tailored set of 21 questions (Supplementary Table 14) and for liver transplant assessment reports, we curated 18 questions (Supplementary Table 18). The ground truth labels were created in a hierarchical manner, with subsequent questions dependent on the answers to previous questions; e.g. if there were no liver lesions in the report, the subsequent questions about lesion size or description were labelled as ”not applicable” (N/A).

### LLM data extraction

We explored four large language models: Llama 3.1 8B, Llama 3.3 70B, OpenBioLLM 70B, and DeepSeek R1 8B, as well as two constrained decoding methods: *Guidance* package (v0.2.0) and *Ollama* package (v0.5.1).

Llama 3.1 8B and DeepSeek R1 8B represent compact models that can be executed on a local desktop or even a high-end laptop, with DeepSeek R1 specifically designed for reasoning. In contrast, Llama 3.3 70B and OpenBioLLM 70B have larger parameter scales and are designed to provide stronger performance and enhanced biomedical domain coverage, while still being feasible to deploy on a single GPU. We deliberately restricted our scope to open-source and locally deployable models and tools, as hospital environments often require on-premise solutions that are independent of commercial cloud services due to strict privacy and security regulations.

All four LLMs were tested with the *Guidance* package; they were accessed through HuggingFace instructs [40–43]. LLM extraction was performed in Python (v3.12.4), running on a single NVIDIA A100 GPU with 80GB of memory.

The 8B-parameter models (Llama 3.1 8B and DeepSeek R1 8B) were also tested using the *Ollama* package. They were accessed via the *Ollama* model repository ([44, 45]) and run locally in Python (v3.13.5) on an Apple M3 Pro chip.

In constrained decoding, each question was prompted separately, with the model instructed to provide a single answer in a specific format (full prompts are shown in Supplementary Tables 24, 25, 26, and 27). For categorical questions, the model was instructed to provide an answer from a predefined set of options, whereas for numeric questions, the model was instructed to provide a single number or date matching a defined regex pattern.

Unconstrained template prompting included questions which combined multiple outputs in a single prompt, with the model instructed to provide answers in a specific format.

To ensure transparency and reproducibility, the complete codebase used for the LLM-based data extraction is publicly available at https://github.com/hmpaverd/LLM-clinical-extraction.

### Model evaluation

We evaluated the performance of the LLMs both in terms of output format and content. Model evaluation and all downstream analyses were performed in Python (v3.13.5) using scipy (v1.15.3) and scikit-klearn (v1.6.1) packages.

To assess format validity, we assessed the output of one model - Llama 3.1 8B - in combination with *Guidance* package, the *Ollama* package, and with no constrained decoding (”unconstrained template prompting”). We used Calibration Set 1, which included 20 patients and 282 radiology reports, with a total of 5614 question-answer pairs. The output was considered valid if it matched the expected format; the format we evaluated the output against was the same as defined in the prompt. We calculated the proportion of valid outputs across all 5614 question-answer pairs, as well as for each individual question. We evaluated outputs only for questions deemed applicable to the report: as per the pipeline’s hierarchical structure, dependent questions (e.g., the date of a previous ablation) were assessed only if their prerequisite conditions were met (e.g., previous ablation was present). Consequently, the final number of evaluated question-answer pairs (n=5,614) is lower than the theoretical maximum of 8,742 (282 reports × 31 questions).

When assessing output content, we calculated the performance metrics for each question for all six model-constrained decoding combinations, as well as uncosntrained template prompting for Llama 3.1 8B. The LLM’s output was evaluated against the manually curated ground truth with each report contributing equally to the final score. Reports without assigned ground truth labels for a given question - i.e. where the question was not applicable - were excluded from the evaluation for that specific question. For categorical questions, we calculated balanced accuracy, specificity, sensitivity, precision, and F1 score; for numeric questions, we calculated the proportion of correct outputs, defined as the number of answers exactly matching the ground truth divided by the total number of answers. Model ’hallucinations’ - instances where the LLM extracted a finding, lesion, or treatment not present in the source text - were penalised as incorrect. For categorical questions, these were recorded as false positives, which directly reduced balanced accuracy and specificity. For numeric extraction tasks, hallucinated counts were evaluated as failing to exactly match the ground truth, thereby reducing the overall proportion of correct answers. The results were then bootstrapped to calculate 95% confidence intervals (CI95%) for each question. Models were compared against each other using bootstrapped confidence intervals, with model A being considered significantly better than model B if the lower limit of Model A’s CI95% was above the upper limit of Model B’s CI95%. While this approach is statistically conservative for paired evaluations, this high threshold ensures that a model is only deemed ’significantly better’ if it demonstrates a robust and unambiguous performance advantage, thereby mitigating the risk of selecting more computationally expensive models based on marginal or noisy gains.

### Full scale data analysis

We applied the best-performing model, Llama 3.3 70B with *Guidance* package, to the full cohort of 835 patients. The extraction pipeline was designed to ensure that the questions were answerable based on the information available in the report. This was achieved by including ”if” statements based on the answers to previous questions, allowing the model to skip questions that were not applicable; for example, if the model identified that there were no liver lesions in the report, it would not be prompted to answer questions about lesion size or description.

The extracted data was then summarised per patient, with binary variables (e.g. cirrhosis, previous locoregional therapies) assigned as positive if any of the reports indicated the presence of the feature, and ordinal variables (e.g. LR lesion grade, cancer differentiation grade) summarised by taking the most severe instance of the finding across all reports. The source of each variable in the summary dataframe was indicated, and additional sources of information from electronic healthcare records and manual annotations were added.

We then performed quantitative analysis of the extracted data. We calculated the sensitivity and specificity of radiological lesion classification for predicting pathology-confirmed HCC among patients with liver lesions, excluding incidental HCCs, including both LR lesion classification (LR-3 or more severe, LR-4 or more severe, LR-5) and descriptive lesion classification (equivocal or more severe, probable HCC or more severe, HCC).

We compared patient characteristics between those with and without HCC (n=166 vs. 611), excluding super-urgent transplants. Within the HCC group, we further examined associations with HCC recurrence (n=15 vs. 151) and mortality (n=35 vs. 131). Categorical variables were assessed using Chi-squared tests and continuous variables with Mann–Whitney U tests. Multiple comparisons correction was applied using the Bonferroni method, with a significance threshold of p<0.05 after correction.

Finally, we assessed the change in primary chronic liver disease (CLD) diagnosis at the time of liver transplant assessment (LTA) and post-transplant at the pathological examination of the explanted liver (PATH). The primary CLD diagnosis was taken as the answer to the first prompt about CLD aetiology, and our analysis did not include secondary and tertiary CLD diagnoses.

### Timeline reconstruction

We reconstructed patient timelines for Calibration Set 1 patients, focusing on two aspects: LR lesions and previous treatments. We compared the performance of Llama 3.3 70B and Llama 3.1 8B, both with *Guidance* package, to the manually curated ground truth.

LR timelines were reconstructed based on radiology reports, with each timepoint on the timeline corresponding to the date of the scan (relative to the transplant date) and annotated with the number of lesions of each LR type extracted from that scan’s report. We compared LLM-extracted and ground truth extracted timelines by calculating the number of lesions correctly identified by the model at each timepoint, as well as the number of lesions missed and added by the model. Additionally, we computed type-level discrepancies: model-added or model-missed LIRADS categories across the full timeline for each patient.

Treatment timelines were reconstructed separately from two sources: radiology reports and liver transplant assessment (LTA) reports. For each treatment type (resection, ablation, embolisation), reported dates were converted to relative day offsets from the transplant date. If no treatment date was available but treatment was reported in a radiology report, we assigned a range spanning from the previous report to the current report date; this was done only for the first report which documented the treatment, and subsequent report were assumed to refer to the same instance unless the date was specified. The resulting treatment timeline was then compared to the manually curated ground truth, with each treatment instance (consisting of treatment type and treatment date or date range) being classified as either correctly identified, missed, or added by the model.

## Data Availability

The data underlying this article cannot be published due to confidentiality considerations.

## Supplementary information

Supplementary figures and tables are included in the ”Supplementary information” file.

## Data availability

The data supporting this study’s findings cannot be made publicly available due to patient confidentiality considerations.

## Code availability

The code and processing pipelines used for LLM data extraction are publicly available in the GitHub repository: https://github.com/hmpaverd/LLM-clinical-extraction.

## Acknowledgements

The authors gratefully acknowledge AstraZeneca UK Limited and the NIHR BioResource (G127831) for funding this work. We thank Jakub Jaworski from the Cambridge University Hospitals NHS Foundation Trust for his work extracting clinical data for this study. H.P. was supported by AstraZeneca UK Limited and the NIHR BioResource (G127831). M.H. was supported by a CRUK Programme Foundation Award (DRCPFA-Jun22/100001) and an MRC research grant (MR/X00970X/1). M.C.O. was supported by the Joseph Mitchell Cancer Research Fund, the Academy of Medical Sciences (G117526) and NIHR (NIHR206092). We thank all the patients and families who participated in the study.

## Author contributions

Conceptualisation: M.C.O., M.H. Design: H.P., M.H., M.C.O. Data Acquisition: H.P., S.B. Software: H.P., Z.G., G.M. Analysis: H.P. Writing - Original Draft: H.P. Writing - Review and Editing: H.P., Z.G., G.M., M.F., S.B., M.H., M.C.O. Supervision: M.H., M.C.O.

## Declaration of generative AI and AI-assisted technologies in the writing process

During the preparation of this work the authors used generative AI in order to format and proof-read the article. After using this tool, the authors reviewed and edited the content as needed and take full responsibility for the content of the publication.

## Abbreviations

A1AT: Alpha-1 Antitrypsin
AIH: Autoimmune Hepatitis
ArLD: Alcohol-related Liver Disease
BMI: Body Mass Index
CI: Confidence Interval
CLD: Chronic Liver Disease
CT: Computed Tomography
DM: Diabetes Mellitus
EHR: Electronic Healthcare Records
FNH: Focal Nodular Hyperplasia
GPT: Generative Pre-trained Transformer
HCC: Hepatocellular Carcinoma
HCV: Hepatitis C Virus
LIRADS: Liver Imaging Reporting and Data System
LLM: Large Language Model
LR: Liver Imaging Reporting and Data System (LIRADS) Grade
LTA: Liver Transplant Assessment
LVI: Large Vessel Invasion
MASH: Metabolic-associated Steatohepatitis
MASLD: Metabolic-associated Steatotic Liver Disease
MRI: Magnetic Resonance Imaging
MVI: Microvascular Invasion
NAFLD: Non-alcoholic Fatty Liver Disease
NASH: Non-alcoholic Steatohepatitis
NLP: Natural Language Processing
OLT: Orthotopic Liver Transplant
PATH: Pathological examination of the explanted liver
PBC: Primary Biliary Cholangitis
PET: Positron Emission Tomography
PSC: Primary Sclerosing Cholangitis
Regex: Regular Expression
RFA: Radiofrequency Ablation
SD: Standard Deviation
T1DM: Type 1 Diabetes Mellitus
T2DM: Type 2 Diabetes Mellitus
TACE: Transarterial Chemoembolisation
TAE: Transarterial Embolisation
TIV: Tumour in Vein
TNM: Tumour, Node, Metastasis (Staging system)
UKELD: United Kingdom Model for End-Stage Liver Disease
US: Ultrasound
VTT: Venous Tumour Thrombus

## Supplementary figures

**Supplementary Figure 1:**
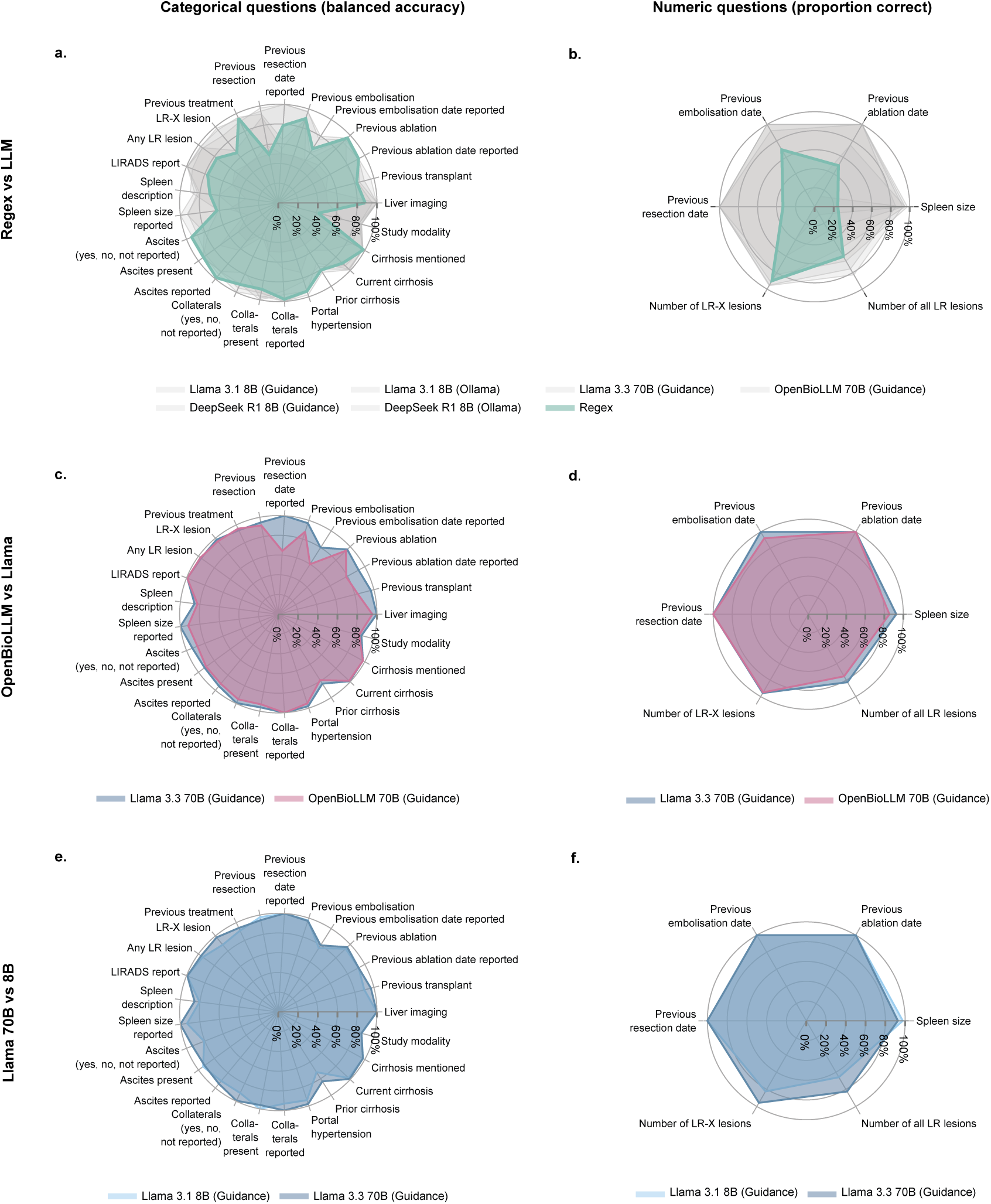
Benchmarking LLMs in radiology reports - model comparison. **a-b** Comparison of regex and LLMs’ performance. Panel **a** shows balanced accuracy and panel **b** proportion correct outputs of regex (in green) vs all other LLMs in grey is shown. **c-d** Comparison of Llama 3.3 70B and its medically fine-tuned version, OpenBioLLM 70B. Panel **c** shows balanced accuracy and panel **d** proportion correct outputs of Llama 3.3 70B vs OpenBioLLM. **e-f** Comparison of Llama 3.3 70B and Llama 3.1 8B. Panel **e** shows balanced accuracy and panel **f** proportion correct outputs of Llama 3.1 8B vs Llama 3.1 70B.

**Supplementary Figure 2:**
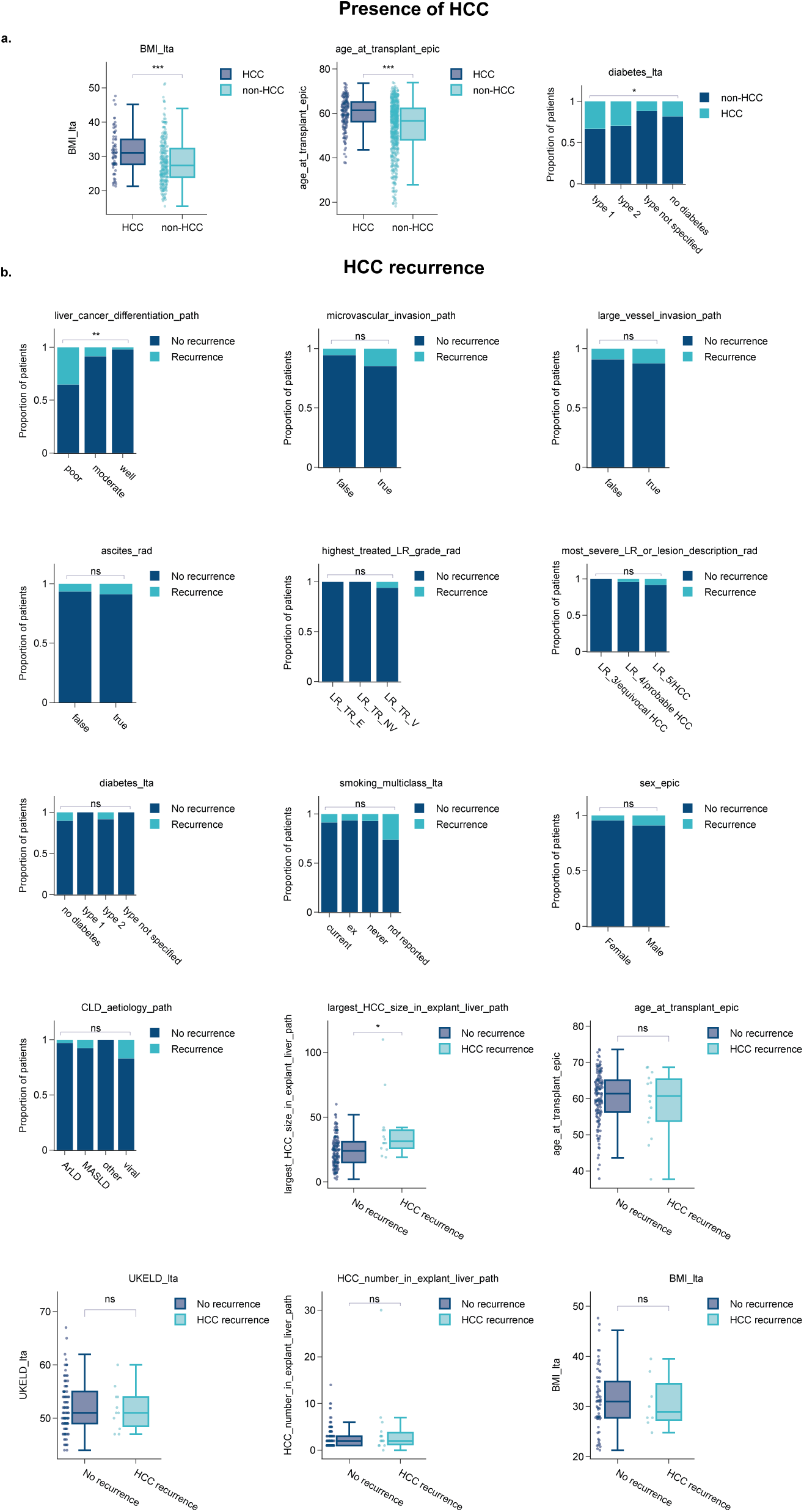
Characteristics of patients with HCC development and recurrence. a. Characteristics of patients with HCC development among the full 835 patient cohort. b. Characteristics of patients with HCC recurrence among the 166 patients with HCC. ***: p<0.001; *: p<0.01; ns: not significant.

**Supplementary Figure 3:**
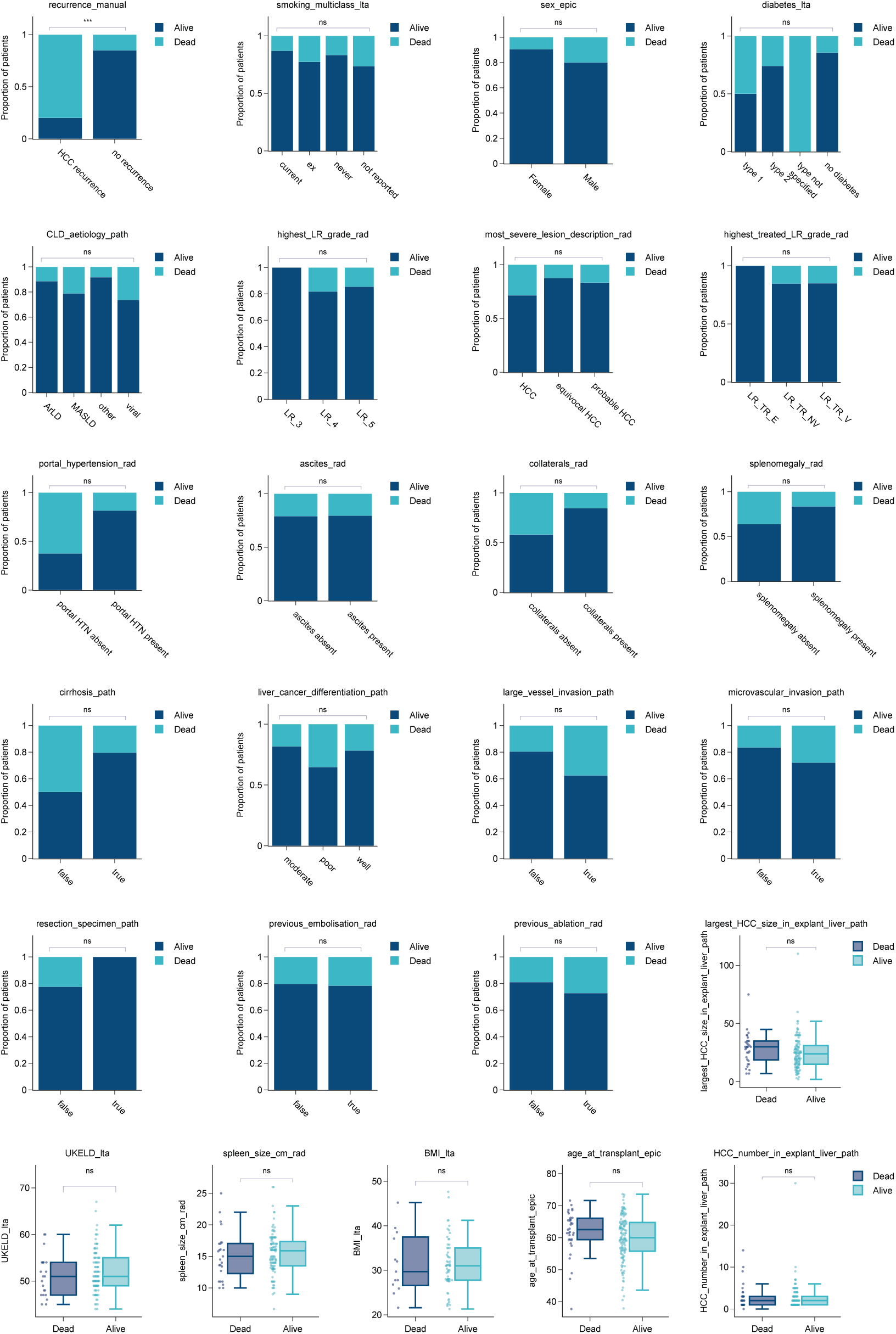
Characteristics associated with mortality in patients with HCC. Comparison of clinical features between patients who were alive versus deceased at the latest follow-up among the 166 patients with HCC. ***: p<0.001; ns: not significant.

**Supplementary Figure 4:**
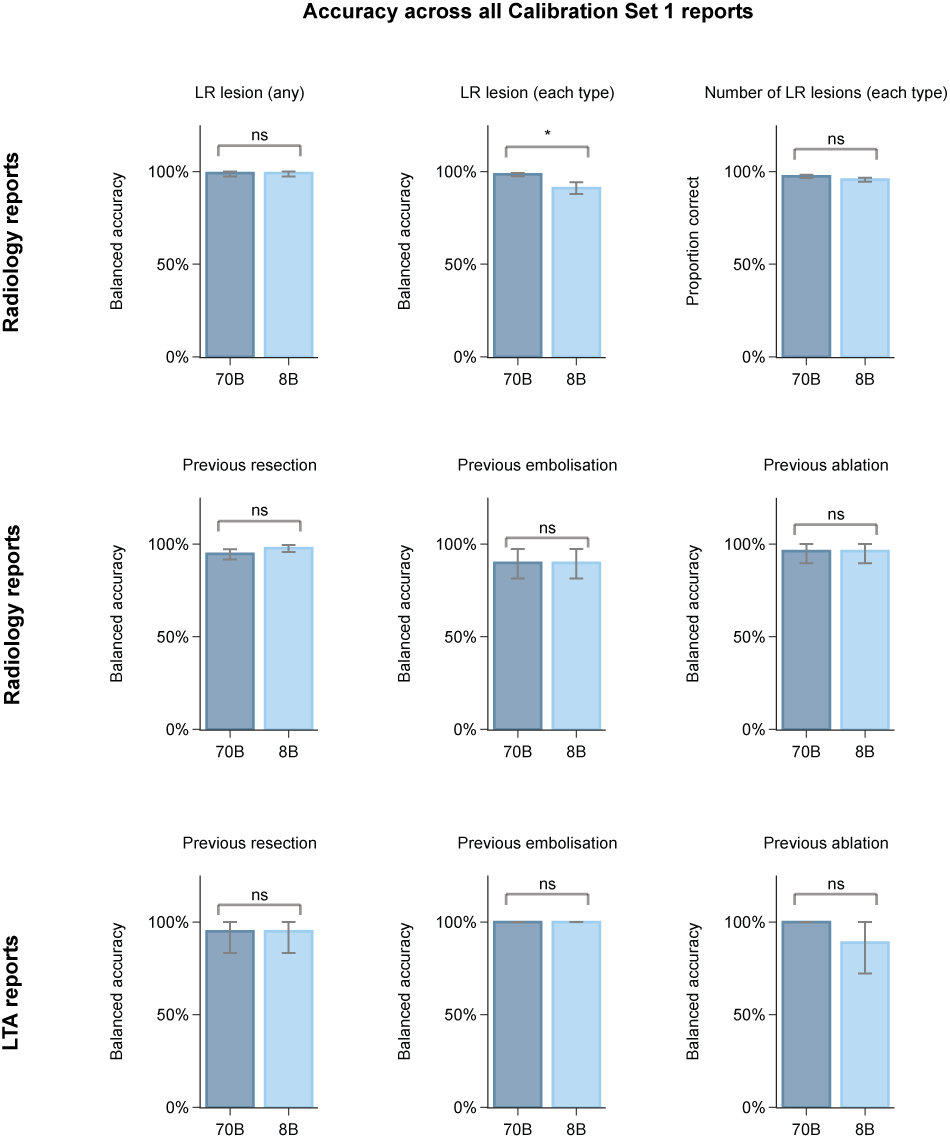
Performance of Llama 3.3 70B and Llama 3.1 8B across all Calibration Set 1 radiology and LTA reports for LR lesion and treatment-related questions.

**Supplementary Figure 5:**
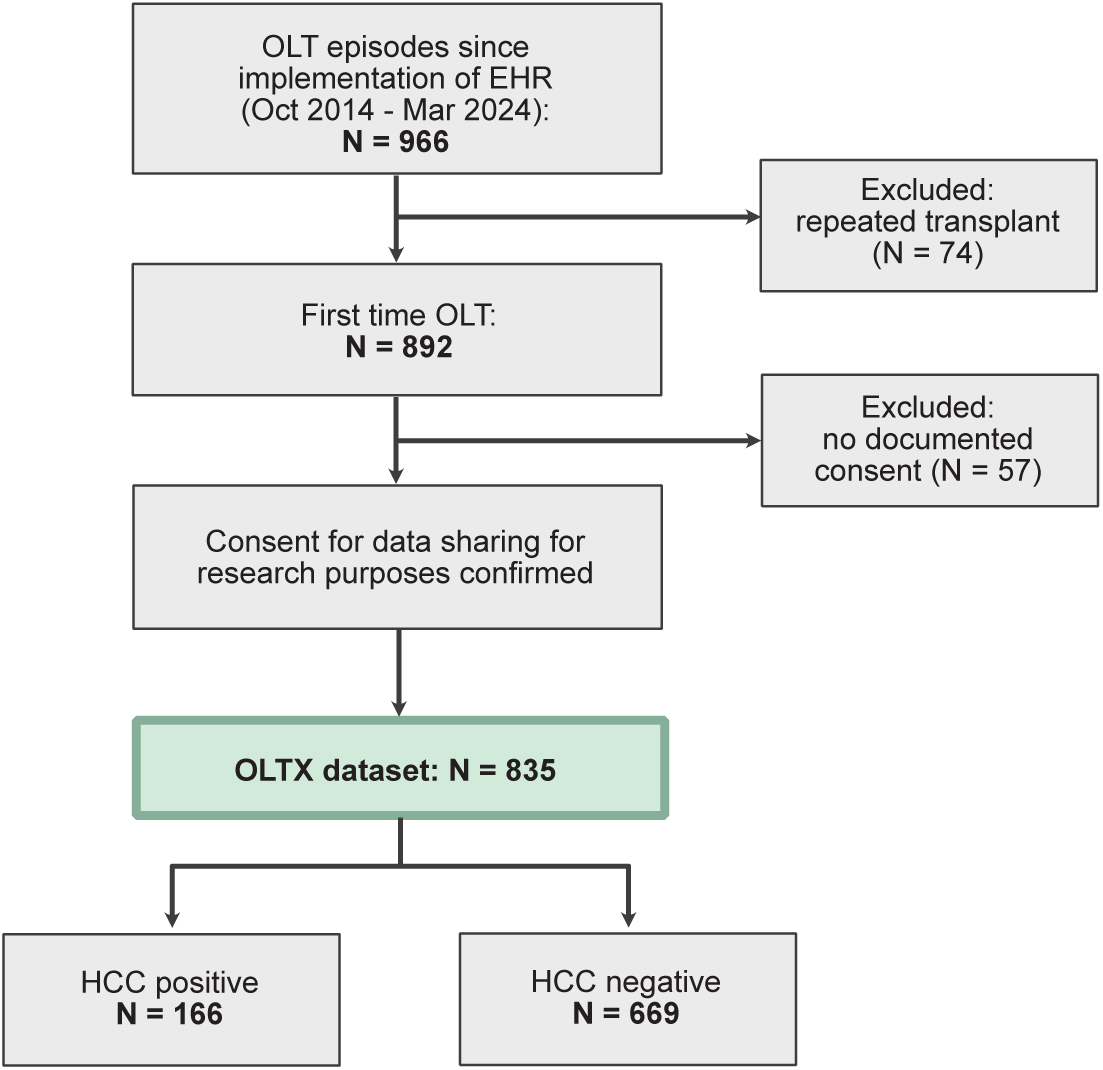
Patient cohort selection flowchart. OLT(X): orthotopic liver transplant. EHR: Electronic Healthcare Record. HCC: hepatocellular carcinoma.

**Supplementary Figure 6:**
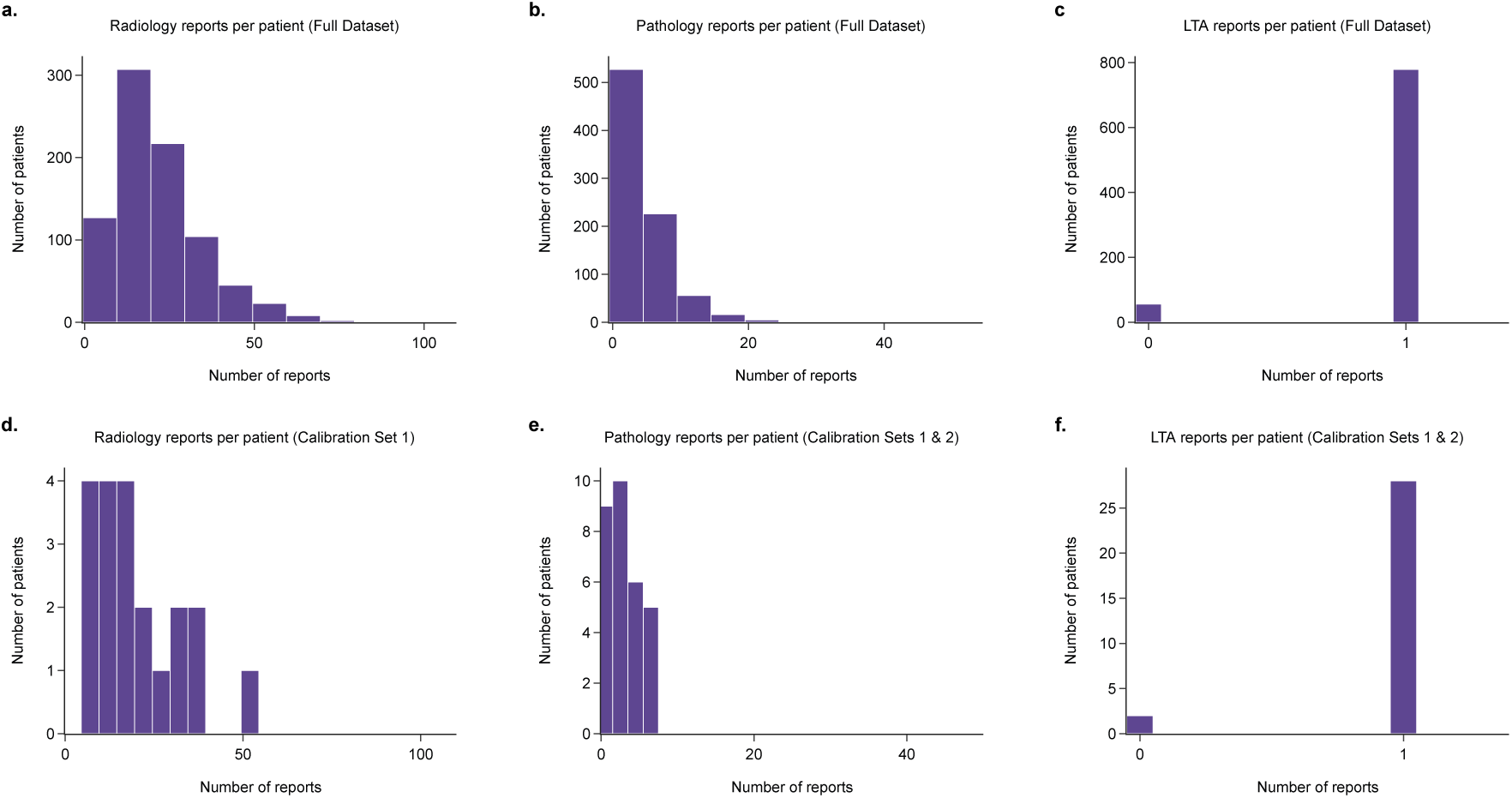
Distribution of report numbers across the patient cohort for (a) radiology, (b) pathology, and (c) liver transplant assessment reports.

**Supplementary Figure 7:**
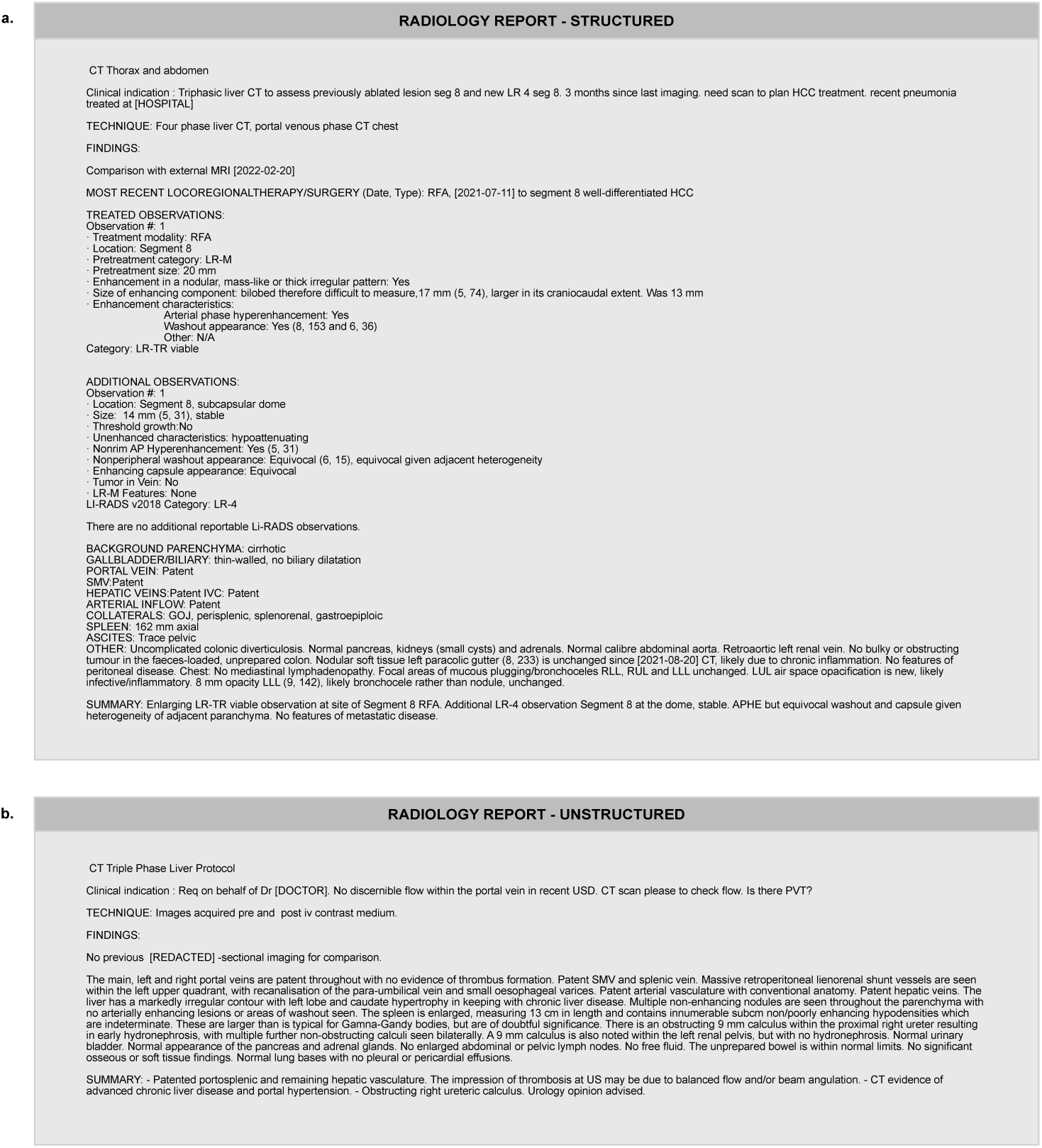
Examples of well structured (a) and unstructured (b) radiology report. The dates have been pseudonymised to preserve patient confidentiality.

**Supplementary Figure 8:**
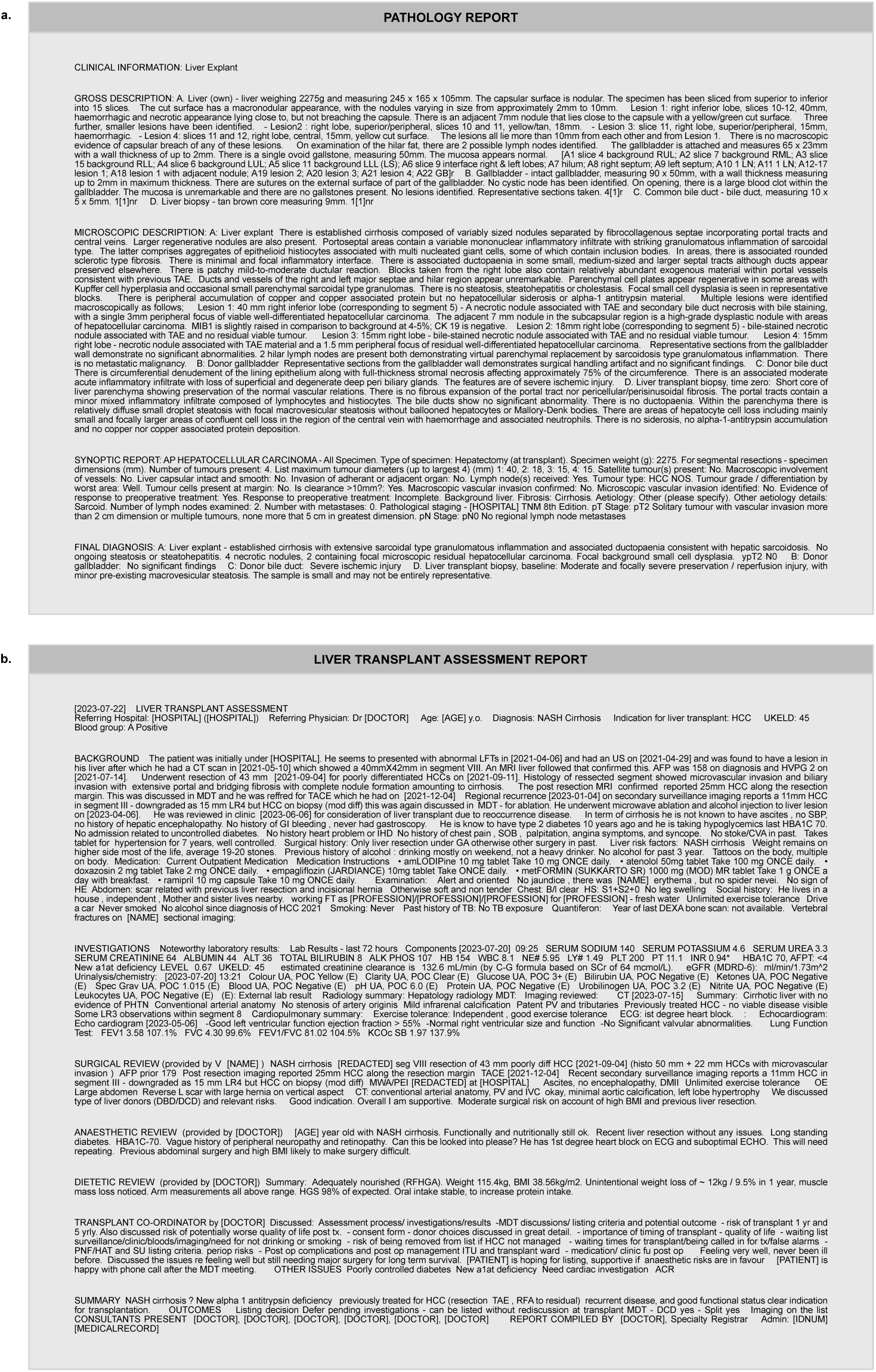
Examples of pathology report (a) and liver transplant assessment report (b). The dates have been pseudonymised to preserve patient confidentiality.

## Supplementary tables

**Supplementary Table 1:** Radiology questions.

| Question | Possible answers |
| --- | --- |
| Does this report describe liver imaging? | "yes", "no" |
| What kind of imaging is described in this report? | "CT", "MRI", "US", "transjugular biopsy", "embolisation", "angio", "cholangiogram", "nuclear medicine", "other" |
| Is cirrhosis explicitly mentioned in this report? | "yes", "no" |
| Is there current diagnosis of cirrhosis? | "yes", "no" |
| Was there a prior diagnosis of cirrhosis? | "yes", "no" |
| Is portal hypertension explicitly diagnosed in this report? | "yes", "no" |
| Is there an explicit comment on presence or absence of collateral vessels? | "yes", "no" |
| Are "collaterals", "shunt vessels" or "varices" explicitly reported as present? | "yes", "no" |
| Are there collateral vessels present? | "yes", "no", "not reported" |
| Is there an explicit comment on the presence or absence of ascites? | "yes", "no" |
| Is ascites explicitly reported as present? | "yes", "no" |
| Is there ascites present? | "yes", "no", "not reported" |
| Is the spleen size explicitly reported in this report? | "yes", "no" |
| What is the size of the spleen? | \(\d+\.?\d*\) |
| What is the description of the spleen? | "splenomegaly", "normal", "not reported" |
| Does this report use LIRADS system? | "yes", "no" |
| Are there any LIRADS observations present? | "yes", "no" |
| How many LIRADS observations are present? | \(\d+\) |
| Are there any LR-X observations present? | "yes", "no" |
| How many LR-X observations are present? | \(\d+\) |
| Does this report describe any previous liver cancer treatment? | "yes", "no" |
| Did the patient have a previous resection for liver cancer? | "yes", "no" |
| Is the date of the previous resection explicitly reported? | "yes", "no" |
| What is the date of the previous resection the patient underwent for liver cancer? | \(\d{4}-\d{2}-\d{2}\) |
| Did the patient have a previous embolisation for liver cancer? | "yes", "no" |
| Is the date of the previous embolisation explicitly reported in this report? | "yes", "no" |
| What is the date of the previous embolisation the patient underwent for liver cancer? | \(\d{4}-\d{2}-\d{2}\) |
| Did the patient have a previous ablation for liver cancer? | "yes", "no" |
| Is the date of the previous ablation mentioned in this report? | "yes", "no" |
| What is the date of the previous ablation the patient underwent for liver cancer? | \(\d{4}-\d{2}-\d{2}\) |
| Did the patient have a previous liver transplant? | "yes", "no" |

**Supplementary Table 2:** Performance metrics for radiology questions - Llama 3.3 70B (Guidance).

| Task | Balanced accuracy | Precision | Sensitivity | Specificity | F1 score | Proportion correct |
| --- | --- | --- | --- | --- | --- | --- |
| Liver imaging | 0.998<br>[0.993, 1.000] | 1.000<br>[1.000, 1.000] | 0.996<br>[0.986, 1.000] | 1.000<br>[1.000, 1.000] | 0.998<br>[0.993, 1.000] | – |
| Study modality | 0.870<br>[0.825, 0.875] | 0.950<br>[0.916, 0.977] | 0.965<br>[0.940, 0.982] | Not defined<br>(multi-class) | 0.956<br>[0.925, 0.980] | – |
| Cirrhosis mentioned | 0.979<br>[0.952, 1.000] | 1.000<br>[1.000, 1.000] | 0.957<br>[0.905, 1.000] | 1.000<br>[1.000, 1.000] | 0.978<br>[0.950, 1.000] | – |
| Current cirrhosis | 0.992<br>[0.972, 1.000] | 1.000<br>[1.000, 1.000] | 0.983<br>[0.943, 1.000] | 1.000<br>[1.000, 1.000] | 0.991<br>[0.971, 1.000] | – |
| Prior cirrhosis | 0.833<br>[0.788, 0.881] | 0.484<br>[0.356, 0.617] | 1.000<br>[1.000, 1.000] | 0.667<br>[0.576, 0.763] | 0.652<br>[0.525, 0.763] | – |
| Portal hypertension | 0.980<br>[0.929, 1.000] | 1.000<br>[1.000, 1.000] | 0.960<br>[0.857, 1.000] | 1.000<br>[1.000, 1.000] | 0.980<br>[0.923, 1.000] | – |
| Collaterals reported | 1.000<br>[1.000, 1.000] | 1.000<br>[1.000, 1.000] | 1.000<br>[1.000, 1.000] | 1.000<br>[1.000, 1.000] | 1.000<br>[1.000, 1.000] | – |
| Collaterals present | 0.961<br>[0.886, 1.000] | 0.984<br>[0.948, 1.000] | 0.984<br>[0.948, 1.000] | 0.938<br>[0.789, 1.000] | 0.984<br>[0.958, 1.000] | – |
| Collaterals (yes, no, not reported) | 0.995<br>[0.983, 1.000] | 0.993<br>[0.979, 1.000] | 0.992<br>[0.976, 1.000] | Not defined<br>(multi-class) | 0.992<br>[0.977, 1.000] | – |
| Ascites reported | 0.944<br>[0.880, 1.000] | 0.971<br>[0.931, 1.000] | 1.000<br>[1.000, 1.000] | 0.889<br>[0.760, 1.000] | 0.985<br>[0.964, 1.000] | – |
| Ascites present | 0.923<br>[0.870, 0.968] | 1.000<br>[1.000, 1.000] | 0.846<br>[0.739, 0.936] | 1.000<br>[1.000, 1.000] | 0.917<br>[0.850, 0.967] | – |
| Ascites (yes, no, not reported) | 0.936<br>[0.892, 0.970] | 0.940<br>[0.910, 0.971] | 0.929<br>[0.881, 0.968] | Not defined<br>(multi-class) | 0.929<br>[0.881, 0.968] | – |
| Spleen size reported | 1.000<br>[1.000, 1.000] | 1.000<br>[1.000, 1.000] | 1.000<br>[1.000, 1.000] | 1.000<br>[1.000, 1.000] | 1.000<br>[1.000, 1.000] | – |
| Spleen size | – | – | – | – | – | 0.926<br>[0.864, 0.975] |
| Spleen description | 0.855<br>[0.776, 0.931] | 0.849<br>[0.798, 0.920] | 0.795<br>[0.681, 0.909] | Not defined<br>(multi-class) | 0.791<br>[0.660, 0.908] | – |
| LIRADS report | 0.992<br>[0.973, 1.000] | 1.000<br>[1.000, 1.000] | 0.984<br>[0.945, 1.000] | 1.000<br>[1.000, 1.000] | 0.992<br>[0.972, 1.000] | – |
| Any LR lesion | 0.967<br>[0.932, 0.992] | 1.000<br>[1.000, 1.000] | 0.934<br>[0.863, 0.984] | 1.000<br>[1.000, 1.000] | 0.966<br>[0.926, 0.992] | – |
| LR-X lesion | 0.981<br>[0.965, 0.992] | 0.822<br>[0.750, 0.886] | 0.981<br>[0.952, 1.000] | 0.980<br>[0.972, 0.988] | 0.895<br>[0.846, 0.933] | – |
| Number of all LR lesions | – | – | – | – | – | 0.825<br>[0.754, 0.889] |
| Number of LR-X lesions | – | – | – | – | – | 0.957<br>[0.945, 0.968] |
| Previous treatment | 0.940<br>[0.902, 0.973] | 0.808<br>[0.686, 0.909] | 1.000<br>[1.000, 1.000] | 0.881<br>[0.805, 0.946] | 0.894<br>[0.814, 0.952] | – |
| Previous resection | 0.947<br>[0.917, 0.972] | 0.520<br>[0.318, 0.714] | 1.000<br>[1.000, 1.000] | 0.894<br>[0.835, 0.945] | 0.684<br>[0.483, 0.833] | – |
| Previous resection date reported | 1.000<br>[1.000, 1.000] | 1.000<br>[1.000, 1.000] | 1.000<br>[1.000, 1.000] | 1.000<br>[1.000, 1.000] | 1.000<br>[1.000, 1.000] | – |
| Previous resection date | – | – | – | – | – | 1.000<br>[1.000, 1.000] |
| Previous embolisation | 0.970<br>[0.945, 0.990] | 0.812<br>[0.667, 0.939] | 1.000<br>[1.000, 1.000] | 0.940<br>[0.890, 0.980] | 0.897<br>[0.800, 0.969] | – |
| Previous embolisation date reported | 0.800<br>[0.643, 0.950] | 0.789<br>[0.588, 0.950] | 1.000<br>[1.000, 1.000] | 0.600<br>[0.286, 0.900] | 0.882<br>[0.741, 0.974] | – |
| Previous embolisation date | – | – | – | – | – | 1.000<br>[1.000, 1.000] |
| Previous ablation | 0.959<br>[0.931, 0.982] | 0.654<br>[0.461, 0.833] | 1.000<br>[1.000, 1.000] | 0.917<br>[0.861, 0.964] | 0.791<br>[0.632, 0.909] | – |
| Previous ablation date reported | 0.929<br>[0.750, 1.000] | 0.889<br>[0.667, 1.000] | 1.000<br>[1.000, 1.000] | 0.857<br>[0.500, 1.000] | 0.941<br>[0.800, 1.000] | – |
| Previous ablation date | – | – | – | – | – | 1.000<br>[1.000, 1.000] |
| Previous transplant | 0.969<br>[0.939, 0.993] | 0.902<br>[0.812, 0.976] | 1.000<br>[1.000, 1.000] | 0.938<br>[0.878, 0.986] | 0.948<br>[0.897, 0.988] | – |

**Supplementary Table 3:**
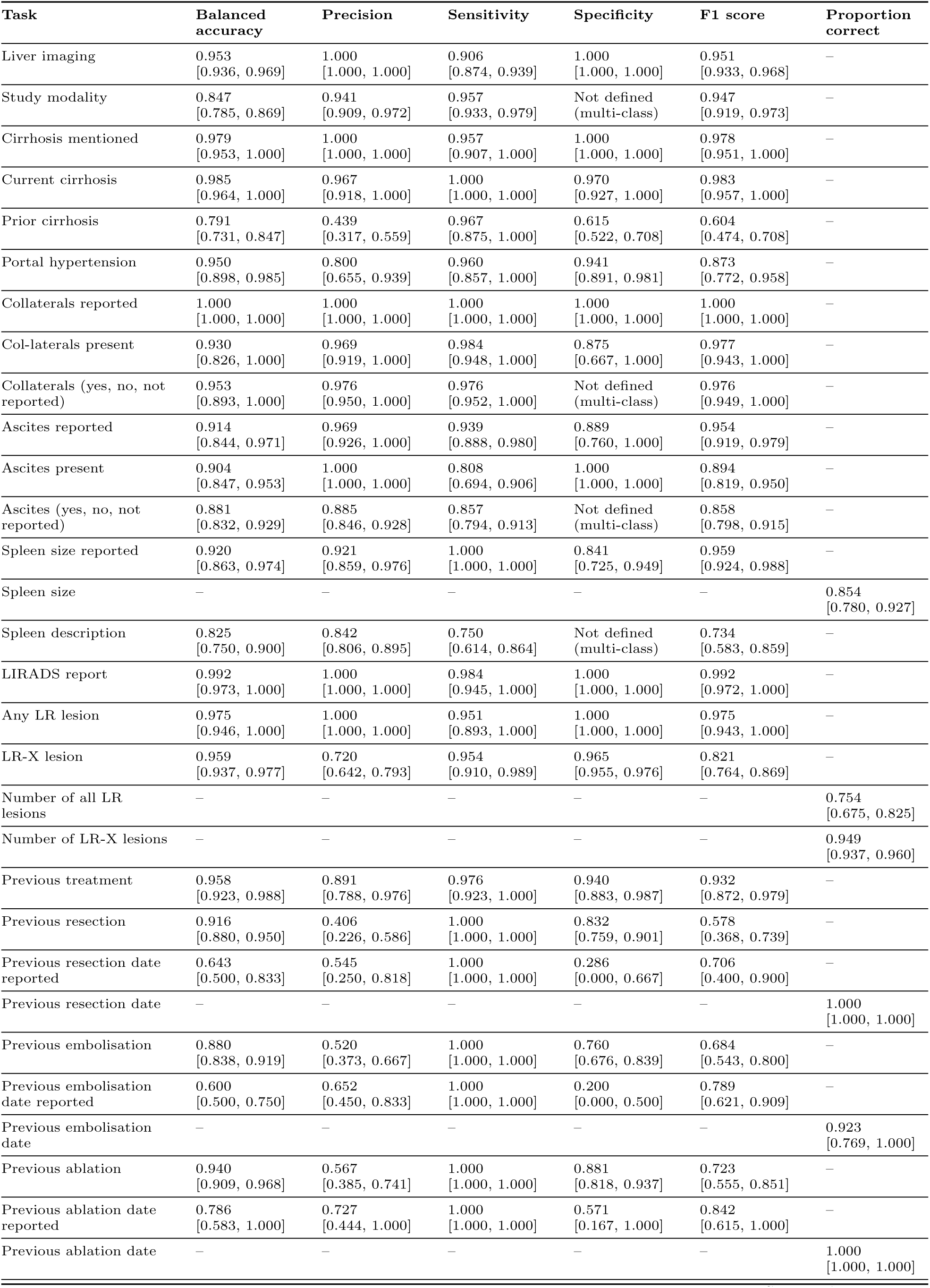
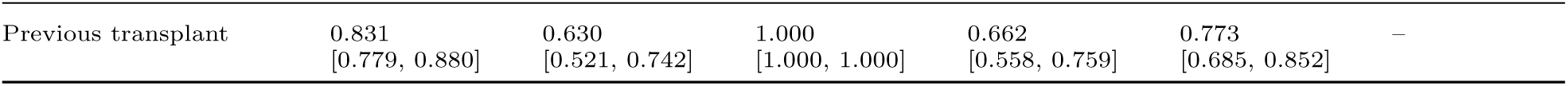
Performance metrics for radiology questions - OpenBioLLM 70B (Guidance).

**Supplementary Table 4:** Performance metrics for radiology questions - Llama 3.1 8B (Guidance).

| Task | Balanced accuracy | Precision | Sensitivity | Specificity | F1 score | Proportion correct |
| --- | --- | --- | --- | --- | --- | --- |
| Liver imaging | 1.000<br>[1.000, 1.000] | 1.000<br>[1.000, 1.000] | 1.000<br>[1.000, 1.000] | 1.000<br>[1.000, 1.000] | 1.000<br>[1.000, 1.000] | – |
| Study modality | 0.872<br>[0.827, 0.875] | 0.960<br>[0.935, 0.983] | 0.968<br>[0.947, 0.986] | Not defined<br>(multi-class) | 0.962<br>[0.938, 0.984] | – |
| Cirrhosis mentioned | 0.979<br>[0.950, 1.000] | 1.000<br>[1.000, 1.000] | 0.957<br>[0.900, 1.000] | 1.000<br>[1.000, 1.000] | 0.978<br>[0.947, 1.000] | – |
| Current cirrhosis | 0.978<br>[0.949, 1.000] | 0.952<br>[0.891, 1.000] | 1.000<br>[1.000, 1.000] | 0.955<br>[0.898, 1.000] | 0.975<br>[0.942, 1.000] | – |
| Prior cirrhosis | 0.723<br>[0.658, 0.782] | 0.367<br>[0.260, 0.473] | 0.967<br>[0.882, 1.000] | 0.479<br>[0.384, 0.583] | 0.532<br>[0.407, 0.637] | – |
| Portal hypertension | 0.940<br>[0.865, 1.000] | 1.000<br>[1.000, 1.000] | 0.880<br>[0.731, 1.000] | 1.000<br>[1.000, 1.000] | 0.936<br>[0.844, 1.000] | – |
| Collaterals reported | 0.931<br>[0.891, 0.965] | 1.000<br>[1.000, 1.000] | 0.863<br>[0.782, 0.931] | 1.000<br>[1.000, 1.000] | 0.926<br>[0.878, 0.964] | – |
| Col-laterals present | 1.000<br>[1.000, 1.000] | 1.000<br>[1.000, 1.000] | 1.000<br>[1.000, 1.000] | 1.000<br>[1.000, 1.000] | 1.000<br>[1.000, 1.000] | – |
| Collaterals (yes, no, not reported) | 0.908<br>[0.834, 0.967] | 0.934<br>[0.898, 0.970] | 0.929<br>[0.881, 0.968] | Not defined<br>(multi-class) | 0.928<br>[0.878, 0.968] | – |
| Ascites reported | 0.907<br>[0.837, 0.959] | 0.978<br>[0.942, 1.000] | 0.889<br>[0.825, 0.949] | 0.926<br>[0.800, 1.000] | 0.931<br>[0.890, 0.966] | – |
| Ascites present | 0.942<br>[0.894, 0.982] | 1.000<br>[1.000, 1.000] | 0.885<br>[0.787, 0.964] | 1.000<br>[1.000, 1.000] | 0.939<br>[0.881, 0.981] | – |
| Ascites (yes, no, not reported) | 0.808<br>[0.734, 0.882] | 0.850<br>[0.801, 0.903] | 0.825<br>[0.754, 0.889] | Not defined<br>(multi-class) | 0.823<br>[0.754, 0.889] | – |
| Spleen size reported | 0.932<br>[0.875, 0.979] | 0.932<br>[0.874, 0.979] | 1.000<br>[1.000, 1.000] | 0.864<br>[0.750, 0.957] | 0.965<br>[0.933, 0.989] | – |
| Spleen size | – | – | – | – | – | 0.976<br>[0.939, 1.000] |
| Spleen description | 0.805<br>[0.703, 0.894] | 0.778<br>[0.677, 0.882] | 0.727<br>[0.591, 0.864] | Not defined<br>(multi-class) | 0.724<br>[0.577, 0.860] | – |
| LIRADS report | 0.992<br>[0.973, 1.000] | 1.000<br>[1.000, 1.000] | 0.984<br>[0.945, 1.000] | 1.000<br>[1.000, 1.000] | 0.992<br>[0.972, 1.000] | – |
| Any LR lesion | 0.946<br>[0.904, 0.982] | 0.909<br>[0.833, 0.970] | 0.984<br>[0.945, 1.000] | 0.908<br>[0.831, 0.969] | 0.945<br>[0.898, 0.982] | – |
| LR-X lesion | 0.883<br>[0.846, 0.922] | 0.717<br>[0.636, 0.795] | 0.796<br>[0.721, 0.875] | 0.970<br>[0.960, 0.980] | 0.754<br>[0.693, 0.818] | – |
| Number of all LR lesions | – | – | – | – | – | 0.659<br>[0.579, 0.738] |
| Number of LR-X lesions | – | – | – | – | – | 0.825<br>[0.803, 0.845] |
| Previous treatment | 0.911<br>[0.858, 0.955] | 0.759<br>[0.640, 0.868] | 0.976<br>[0.923, 1.000] | 0.845<br>[0.759, 0.921] | 0.854<br>[0.769, 0.926] | – |
| Previous resection | 0.978<br>[0.957, 0.995] | 0.722<br>[0.500, 0.926] | 1.000<br>[1.000, 1.000] | 0.956<br>[0.913, 0.991] | 0.839<br>[0.667, 0.962] | – |
| Previous resection date reported | 1.000<br>[1.000, 1.000] | 1.000<br>[1.000, 1.000] | 1.000<br>[1.000, 1.000] | 1.000<br>[1.000, 1.000] | 1.000<br>[1.000, 1.000] | – |
| Previous resection date | – | – | – | – | – | 1.000<br>[1.000, 1.000] |
| Previous embolisation | 0.975<br>[0.953, 0.995] | 0.839<br>[0.704, 0.962] | 1.000<br>[1.000, 1.000] | 0.950<br>[0.905, 0.990] | 0.912<br>[0.826, 0.980] | – |
| Previous embolisation date reported | 0.767<br>[0.633, 0.900] | 1.000<br>[1.000, 1.000] | 0.533<br>[0.267, 0.800] | 1.000<br>[1.000, 1.000] | 0.696<br>[0.421, 0.889] | – |
| Previous embolisation date | – | – | – | – | – | 1.000<br>[1.000, 1.000] |
| Previous ablation | 0.936<br>[0.903, 0.966] | 0.548<br>[0.368, 0.714] | 1.000<br>[1.000, 1.000] | 0.872<br>[0.806, 0.933] | 0.708<br>[0.538, 0.833] | – |
| Previous ablation date reported | 0.929<br>[0.750, 1.000] | 0.889<br>[0.667, 1.000] | 1.000<br>[1.000, 1.000] | 0.857<br>[0.500, 1.000] | 0.941<br>[0.800, 1.000] | – |
| Previous ablation date | – | – | – | – | – | 1.000<br>[1.000, 1.000] |
| Previous transplant | 0.900<br>[0.851, 0.939] | 0.742<br>[0.634, 0.846] | 1.000<br>[1.000, 1.000] | 0.800<br>[0.701, 0.878] | 0.852<br>[0.776, 0.917] | – |

**Supplementary Table 5:** Performance metrics for radiology questions - Llama 3.1 8B (Ollama).

| Task | Balanced accuracy | Precision | Sensitivity | Specificity | F1 score | Proportion correct |
| --- | --- | --- | --- | --- | --- | --- |
| Liver imaging | 1.000<br>[1.000, 1.000] | 1.000<br>[1.000, 1.000] | 1.000<br>[1.000, 1.000] | 1.000<br>[1.000, 1.000] | 1.000<br>[1.000, 1.000] | – |
| Study modality | 0.861<br>[0.801, 0.875] | 0.956<br>[0.926, 0.980] | 0.965<br>[0.940, 0.986] | Not defined<br>(multi-class) | 0.958<br>[0.930, 0.982] | – |
| Cirrhosis mentioned | 0.964<br>[0.929, 0.992] | 1.000<br>[1.000, 1.000] | 0.929<br>[0.859, 0.985] | 1.000<br>[1.000, 1.000] | 0.963<br>[0.924, 0.992] | – |
| Current cirrhosis | 0.932<br>[0.885, 0.972] | 1.000<br>[1.000, 1.000] | 0.864<br>[0.771, 0.943] | 1.000<br>[1.000, 1.000] | 0.927<br>[0.871, 0.971] | – |
| Prior cirrhosis | 0.764<br>[0.696, 0.828] | 0.418<br>[0.303, 0.537] | 0.933<br>[0.829, 1.000] | 0.594<br>[0.505, 0.689] | 0.577<br>[0.452, 0.690] | – |
| Portal hypertension | 0.835<br>[0.740, 0.929] | 0.944<br>[0.833, 1.000] | 0.680<br>[0.500, 0.864] | 0.990<br>[0.969, 1.000] | 0.791<br>[0.634, 0.917] | – |
| Collaterals reported | 0.894<br>[0.848, 0.935] | 1.000<br>[1.000, 1.000] | 0.787<br>[0.695, 0.870] | 1.000<br>[1.000, 1.000] | 0.881<br>[0.820, 0.931] | – |
| Col-laterals present | 0.984<br>[0.960, 1.000] | 1.000<br>[1.000, 1.000] | 0.969<br>[0.919, 1.000] | 1.000<br>[1.000, 1.000] | 0.984<br>[0.958, 1.000] | – |
| Collaterals (yes, no, not reported) | 0.782<br>[0.687, 0.870] | 0.861<br>[0.793, 0.925] | 0.865<br>[0.802, 0.921] | Not defined<br>(multi-class) | 0.861<br>[0.791, 0.919] | – |
| Ascites reported | 0.902<br>[0.832, 0.955] | 0.978<br>[0.935, 1.000] | 0.879<br>[0.806, 0.938] | 0.926<br>[0.806, 1.000] | 0.926<br>[0.881, 0.960] | – |
| Ascites present | 0.942<br>[0.897, 0.981] | 1.000<br>[1.000, 1.000] | 0.885<br>[0.793, 0.962] | 1.000<br>[1.000, 1.000] | 0.939<br>[0.885, 0.981] | – |
| Ascites (yes, no, not reported) | 0.761<br>[0.677, 0.837] | 0.812<br>[0.737, 0.881] | 0.778<br>[0.706, 0.849] | Not defined<br>(multi-class) | 0.784<br>[0.712, 0.852] | – |
| Spleen size reported | 0.611<br>[0.524, 0.704] | 0.750<br>[0.632, 0.857] | 0.585<br>[0.481, 0.688] | 0.636<br>[0.490, 0.787] | 0.658<br>[0.559, 0.744] | – |
| Spleen size | – | – | – | – | – | 0.951<br>[0.902, 0.988] |
| Spleen description | 0.898<br>[0.811, 0.967] | 0.904<br>[0.838, 0.970] | 0.864<br>[0.750, 0.955] | Not defined<br>(multi-class) | 0.874<br>[0.772, 0.956] | – |
| LIRADS report | 0.937<br>[0.889, 0.977] | 0.921<br>[0.846, 0.984] | 0.951<br>[0.891, 1.000] | 0.923<br>[0.855, 0.985] | 0.935<br>[0.882, 0.977] | – |
| Any LR lesion | 0.953<br>[0.917, 0.985] | 0.923<br>[0.852, 0.982] | 0.984<br>[0.945, 1.000] | 0.923<br>[0.852, 0.983] | 0.952<br>[0.910, 0.985] | – |
| LR-X lesion | 0.690<br>[0.668, 0.711] | 0.133<br>[0.109, 0.160] | 0.972<br>[0.939, 1.000] | 0.408<br>[0.380, 0.437] | 0.235<br>[0.195, 0.274] | – |
| Number of all LR lesions | – | – | – | – | – | 0.095<br>[0.048, 0.151] |
| Number of LR-X lesions | – | – | – | – | – | 0.385<br>[0.358, 0.411] |
| Previous treatment | 0.744<br>[0.693, 0.801] | 0.494<br>[0.383, 0.598] | 1.000<br>[1.000, 1.000] | 0.488<br>[0.386, 0.602] | 0.661<br>[0.554, 0.748] | – |
| Previous resection | 0.885<br>[0.842, 0.921] | 0.333<br>[0.184, 0.477] | 1.000<br>[1.000, 1.000] | 0.770<br>[0.684, 0.842] | 0.500<br>[0.311, 0.646] | – |
| Previous resection date reported | 0.548<br>[0.263, 0.833] | 0.500<br>[0.143, 0.857] | 0.667<br>[0.250, 1.000] | 0.429<br>[0.000, 0.833] | 0.571<br>[0.200, 0.834] | – |
| Previous resection date | – | – | – | – | – | 0.000<br>[0.000, 0.000] |
| Previous embolisation | 0.900<br>[0.858, 0.939] | 0.565<br>[0.412, 0.714] | 1.000<br>[1.000, 1.000] | 0.800<br>[0.716, 0.878] | 0.722<br>[0.583, 0.833] | – |
| Previous embolisation date reported | 0.500<br>[0.344, 0.667] | 0.600<br>[0.389, 0.810] | 0.800<br>[0.588, 1.000] | 0.200<br>[0.000, 0.500] | 0.686<br>[0.500, 0.842] | – |
| Previous embolisation date | – | – | – | – | – | 0.154<br>[0.000, 0.385] |
| Previous ablation | 0.899<br>[0.859, 0.936] | 0.436<br>[0.279, 0.591] | 1.000<br>[1.000, 1.000] | 0.798<br>[0.718, 0.873] | 0.607<br>[0.436, 0.743] | – |
| Previous ablation date reported | 0.929<br>[0.750, 1.000] | 0.889<br>[0.667, 1.000] | 1.000<br>[1.000, 1.000] | 0.857<br>[0.500, 1.000] | 0.941<br>[0.800, 1.000] | – |
| Previous ablation date | – | – | – | – | – | 0.750<br>[0.500, 1.000] |
| Previous transplant | 0.850<br>[0.799, 0.898] | 0.657<br>[0.551, 0.769] | 1.000<br>[1.000, 1.000] | 0.700<br>[0.597, 0.795] | 0.793<br>[0.710, 0.870] | – |

**Supplementary Table 6:**
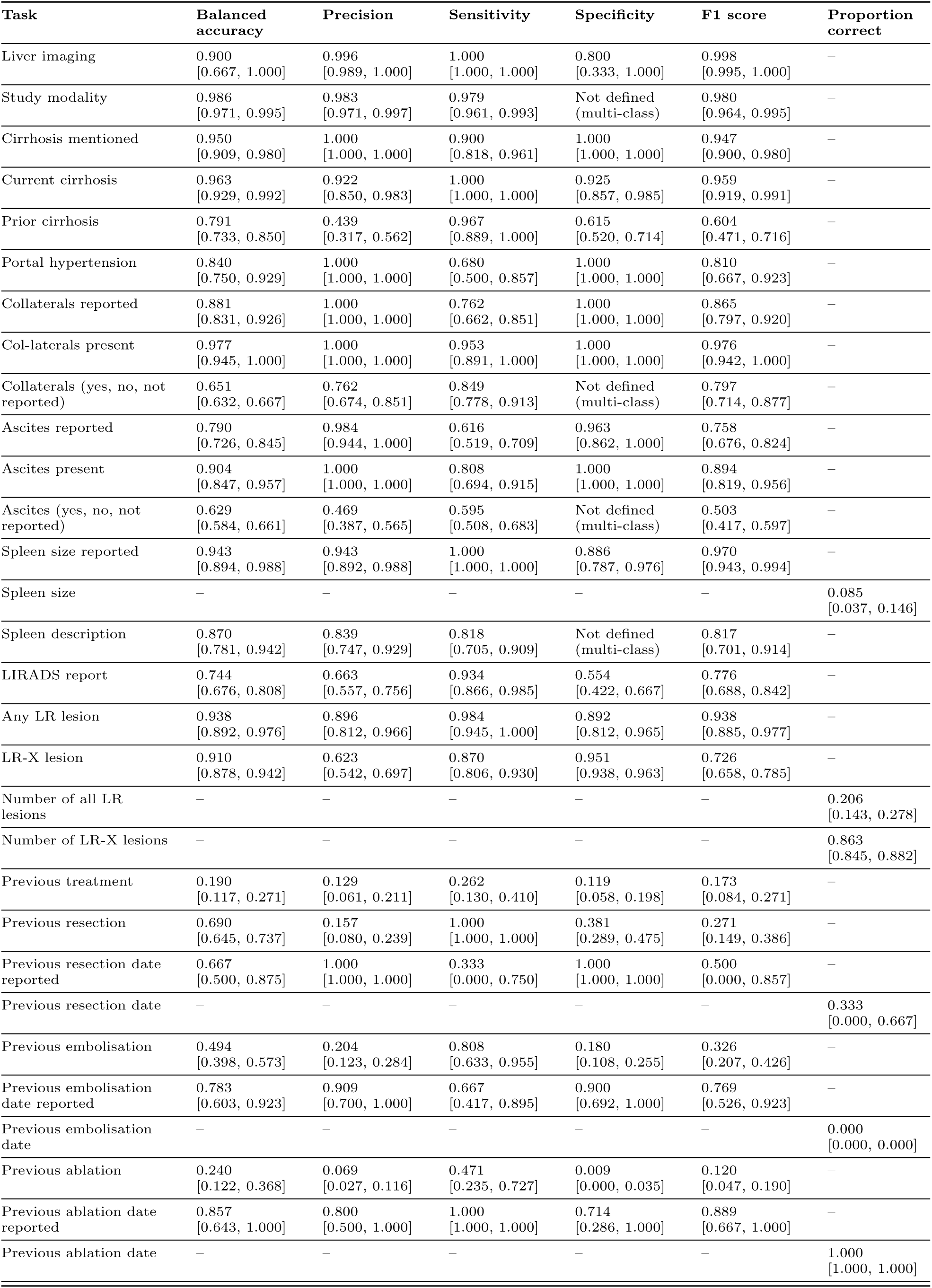
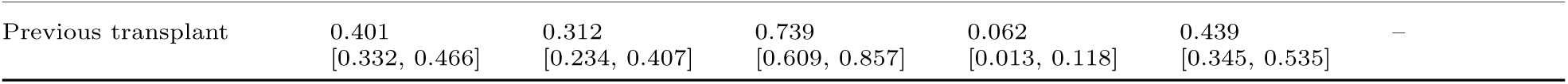
Performance metrics for radiology questions - Llama 3.1 8B (Unconstrained).

**Supplementary Table 7:**
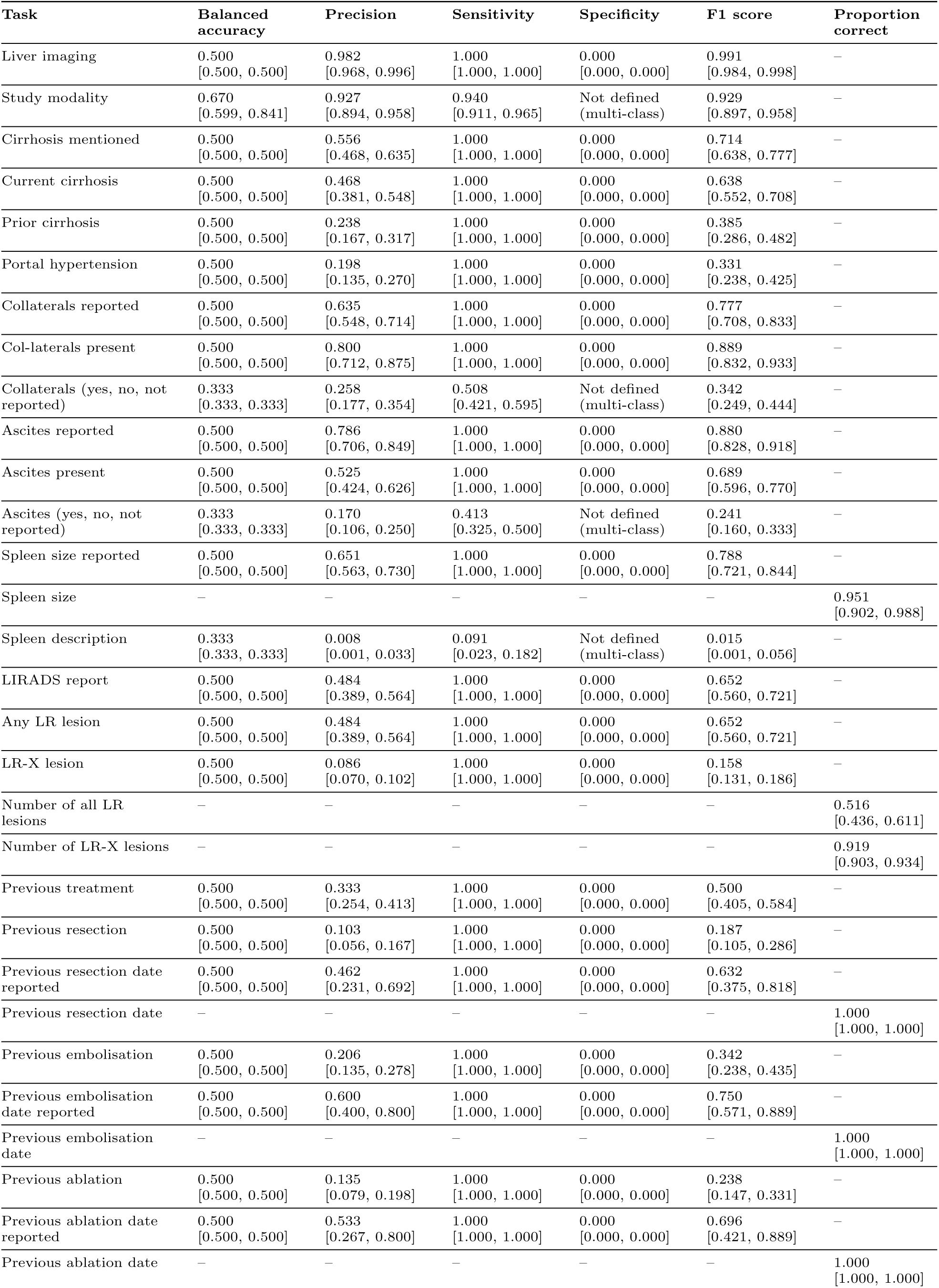
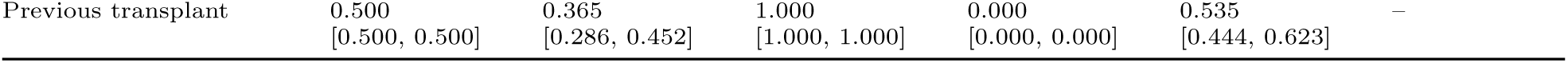
Performance metrics for radiology questions - DeepSeek R1 8B (Guidance).

**Supplementary Table 8:** Performance metrics for radiology questions - Ollama DeepSeek R1 8B.

| Task | Balanced accuracy | Precision | Sensitivity | Specificity | F1 score | Proportion correct |
| --- | --- | --- | --- | --- | --- | --- |
| Liver imaging | 0.998<br>[0.995, 1.000] | 1.000<br>[1.000, 1.000] | 0.996<br>[0.989, 1.000] | 1.000<br>[1.000, 1.000] | 0.998<br>[0.994, 1.000] | – |
| Study modality | 0.841<br>[0.776, 0.860] | 0.923<br>[0.890, 0.954] | 0.936<br>[0.908, 0.961] | Not defined<br>(multi-class) | 0.927<br>[0.895, 0.956] | – |
| Cirrhosis mentioned | 0.857<br>[0.787, 0.919] | 0.844<br>[0.754, 0.921] | 0.929<br>[0.857, 0.985] | 0.786<br>[0.661, 0.893] | 0.884<br>[0.821, 0.936] | – |
| Current cirrhosis | 0.700<br>[0.641, 0.761] | 0.598<br>[0.500, 0.696] | 0.983<br>[0.944, 1.000] | 0.418<br>[0.300, 0.534] | 0.744<br>[0.662, 0.814] | – |
| Prior cirrhosis | 0.568<br>[0.536, 0.602] | 0.265<br>[0.183, 0.348] | 1.000<br>[1.000, 1.000] | 0.135<br>[0.072, 0.204] | 0.420<br>[0.309, 0.517] | – |
| Portal hypertension | 0.742<br>[0.652, 0.814] | 0.355<br>[0.242, 0.477] | 0.880<br>[0.737, 1.000] | 0.604<br>[0.505, 0.697] | 0.506<br>[0.376, 0.627] | – |
| Collaterals reported | 0.661<br>[0.591, 0.738] | 0.722<br>[0.638, 0.806] | 0.975<br>[0.938, 1.000] | 0.348<br>[0.209, 0.500] | 0.830<br>[0.765, 0.886] | – |
| Col-laterals present | 0.523<br>[0.483, 0.600] | 0.808<br>[0.718, 0.886] | 0.984<br>[0.952, 1.000] | 0.062<br>[0.000, 0.211] | 0.887<br>[0.827, 0.938] | – |
| Collaterals (yes, no, not reported) | 0.377<br>[0.349, 0.415] | 0.586<br>[0.457, 0.685] | 0.556<br>[0.468, 0.643] | Not defined<br>(multi-class) | 0.438<br>[0.334, 0.542] | – |
| Ascites reported | 0.623<br>[0.539, 0.716] | 0.832<br>[0.761, 0.895] | 0.949<br>[0.904, 0.990] | 0.296<br>[0.136, 0.482] | 0.887<br>[0.838, 0.927] | – |
| Ascites present | 0.733<br>[0.643, 0.822] | 0.717<br>[0.594, 0.831] | 0.827<br>[0.717, 0.927] | 0.638<br>[0.500, 0.774] | 0.768<br>[0.674, 0.850] | – |
| Ascites (yes, no, not reported) | 0.376<br>[0.333, 0.425] | 0.275<br>[0.174, 0.392] | 0.437<br>[0.349, 0.516] | Not defined<br>(multi-class) | 0.297<br>[0.206, 0.386] | – |
| Spleen size reported | 0.448<br>[0.388, 0.515] | 0.623<br>[0.526, 0.710] | 0.805<br>[0.714, 0.890] | 0.091<br>[0.020, 0.186] | 0.702<br>[0.618, 0.772] | – |
| Spleen size | – | – | – | – | – | 0.927<br>[0.878, 0.976] |
| Spleen description | 0.606<br>[0.487, 0.710] | 0.809<br>[0.721, 0.915] | 0.455<br>[0.318, 0.614] | Not defined<br>(multi-class) | 0.489<br>[0.330, 0.651] | – |
| LIRADS report | 0.500<br>[0.500, 0.500] | 0.484<br>[0.389, 0.564] | 1.000<br>[1.000, 1.000] | 0.000<br>[0.000, 0.000] | 0.652<br>[0.560, 0.721] | – |
| Any LR lesion | 0.546<br>[0.515, 0.586] | 0.508<br>[0.412, 0.595] | 1.000<br>[1.000, 1.000] | 0.092<br>[0.029, 0.172] | 0.674<br>[0.583, 0.746] | – |
| LR-X lesion | 0.500<br>[0.500, 0.500] | 0.086<br>[0.070, 0.102] | 1.000<br>[1.000, 1.000] | 0.000<br>[0.000, 0.000] | 0.158<br>[0.131, 0.186] | – |
| Number of all LR lesions | – | – | – | – | – | 0.294<br>[0.214, 0.373] |
| Number of LR-X lesions | – | – | – | – | – | 0.597<br>[0.572, 0.625] |
| Previous treatment | 0.512<br>[0.500, 0.530] | 0.339<br>[0.260, 0.419] | 1.000<br>[1.000, 1.000] | 0.024<br>[0.000, 0.061] | 0.506<br>[0.413, 0.591] | – |
| Previous resection | 0.588<br>[0.555, 0.626] | 0.123<br>[0.065, 0.194] | 1.000<br>[1.000, 1.000] | 0.177<br>[0.109, 0.252] | 0.218<br>[0.122, 0.326] | – |
| Previous resection date reported | 0.500<br>[0.500, 0.500] | 0.462<br>[0.231, 0.692] | 1.000<br>[1.000, 1.000] | 0.000<br>[0.000, 0.000] | 0.632<br>[0.375, 0.818] | – |
| Previous resection date | – | – | – | – | – | 0.000<br>[0.000, 0.000] |
| Previous embolisation | 0.500<br>[0.500, 0.500] | 0.206<br>[0.135, 0.278] | 1.000<br>[1.000, 1.000] | 0.000<br>[0.000, 0.000] | 0.342<br>[0.238, 0.435] | – |
| Previous embolisation date reported | 0.517<br>[0.410, 0.643] | 0.609<br>[0.409, 0.800] | 0.933<br>[0.786, 1.000] | 0.100<br>[0.000, 0.333] | 0.737<br>[0.562, 0.878] | – |
| Previous embolisation date | – | – | – | – | – | 0.000<br>[0.000, 0.000] |
| Previous ablation | 0.560<br>[0.530, 0.591] | 0.150<br>[0.089, 0.217] | 1.000<br>[1.000, 1.000] | 0.119<br>[0.061, 0.182] | 0.262<br>[0.164, 0.357] | – |
| Previous ablation date reported | 0.571<br>[0.500, 0.722] | 0.571<br>[0.308, 0.800] | 1.000<br>[1.000, 1.000] | 0.143<br>[0.000, 0.444] | 0.727<br>[0.471, 0.889] | – |
| Previous ablation date | – | – | – | – | – | 0.125<br>[0.000, 0.375] |
| Previous transplant | 0.500<br>[0.500, 0.500] | 0.365<br>[0.286, 0.452] | 1.000<br>[1.000, 1.000] | 0.000<br>[0.000, 0.000] | 0.535<br>[0.444, 0.623] | – |

**Supplementary Table 9:** Performance metrics for radiology questions - regular expression search.

| Task | Balanced accuracy | Precision | Sensitivity | Specificity | F1 score | Proportion correct |
| --- | --- | --- | --- | --- | --- | --- |
| Liver imaging | 0.880<br>[0.634, 0.989] | 0.996<br>[0.989, 1.000] | 0.960<br>[0.935, 0.982] | 0.800<br>[0.319, 1.000] | 0.978<br>[0.963, 0.991] | – |
| Study modality | 0.419<br>[0.334, 0.566] | 0.795<br>[0.746, 0.838] | 0.745<br>[0.688, 0.794] | Not defined<br>(multi-class) | 0.739<br>[0.680, 0.790] | – |
| Cirrhosis mentioned | 0.986<br>[0.963, 1.000] | 1.000<br>[1.000, 1.000] | 0.971<br>[0.926, 1.000] | 1.000<br>[1.000, 1.000] | 0.986<br>[0.962, 1.000] | – |
| Current cirrhosis | 0.892<br>[0.839, 0.942] | 0.836<br>[0.736, 0.919] | 0.949<br>[0.889, 1.000] | 0.836<br>[0.740, 0.918] | 0.889<br>[0.822, 0.940] | – |
| Prior cirrhosis | 0.807<br>[0.762, 0.854] | 0.448<br>[0.324, 0.567] | 1.000<br>[1.000, 1.000] | 0.615<br>[0.525, 0.708] | 0.619<br>[0.489, 0.724] | – |
| Portal hypertension | 0.940<br>[0.872, 0.990] | 0.852<br>[0.704, 0.968] | 0.920<br>[0.789, 1.000] | 0.960<br>[0.919, 0.991] | 0.885<br>[0.776, 0.967] | – |
| Collaterals reported | 0.978<br>[0.944, 1.000] | 0.976<br>[0.936, 1.000] | 1.000<br>[1.000, 1.000] | 0.957<br>[0.889, 1.000] | 0.988<br>[0.967, 1.000] | – |
| Col-laterals present | 0.891<br>[0.785, 0.985] | 0.954<br>[0.899, 1.000] | 0.969<br>[0.917, 1.000] | 0.812<br>[0.600, 1.000] | 0.961<br>[0.922, 0.992] | – |
| Collaterals (yes, no, not reported) | 0.913<br>[0.840, 0.975] | 0.945<br>[0.908, 0.978] | 0.944<br>[0.905, 0.976] | Not defined<br>(multi-class) | 0.944<br>[0.904, 0.977] | – |
| Ascites reported | 0.985<br>[0.965, 1.000] | 1.000<br>[1.000, 1.000] | 0.970<br>[0.931, 1.000] | 1.000<br>[1.000, 1.000] | 0.985<br>[0.964, 1.000] | – |
| Ascites present | 0.937<br>[0.887, 0.980] | 0.911<br>[0.827, 0.981] | 0.981<br>[0.935, 1.000] | 0.894<br>[0.800, 0.977] | 0.944<br>[0.891, 0.982] | – |
| Ascites (yes, no, not reported) | 0.944<br>[0.906, 0.979] | 0.942<br>[0.907, 0.977] | 0.937<br>[0.889, 0.976] | Not defined<br>(multi-class) | 0.936<br>[0.888, 0.976] | – |
| Spleen size reported | 0.628<br>[0.583, 0.675] | 1.000<br>[1.000, 1.000] | 0.256<br>[0.167, 0.349] | 1.000<br>[1.000, 1.000] | 0.408<br>[0.286, 0.518] | – |
| Spleen size | – | – | – | – | – | 0.244<br>[0.159, 0.341] |
| Spleen description | 0.698<br>[0.667, 0.746] | 0.772<br>[0.254, 0.818] | 0.568<br>[0.409, 0.705] | Not defined<br>(multi-class) | 0.457<br>[0.292, 0.636] | – |
| LIRADS report | 0.770<br>[0.703, 0.833] | 1.000<br>[1.000, 1.000] | 0.541<br>[0.407, 0.667] | 1.000<br>[1.000, 1.000] | 0.702<br>[0.578, 0.800] | – |
| Any LR lesion | 0.770<br>[0.703, 0.833] | 1.000<br>[1.000, 1.000] | 0.541<br>[0.407, 0.667] | 1.000<br>[1.000, 1.000] | 0.702<br>[0.578, 0.800] | – |
| LR-X lesion | 0.659<br>[0.613, 0.707] | 0.494<br>[0.388, 0.611] | 0.352<br>[0.264, 0.444] | 0.966<br>[0.956, 0.976] | 0.411<br>[0.318, 0.500] | – |
| Number of all LR lesions | – | – | – | – | – | 0.603<br>[0.516, 0.683] |
| Number of LR-X lesions | – | – | – | – | – | 0.898<br>[0.881, 0.915] |
| Previous treatment | 0.946<br>[0.901, 0.987] | 0.929<br>[0.842, 1.000] | 0.929<br>[0.848, 1.000] | 0.964<br>[0.919, 1.000] | 0.929<br>[0.868, 0.977] | – |
| Previous resection | 0.500<br>[0.500, 0.500] | 0.103<br>[0.056, 0.167] | 1.000<br>[1.000, 1.000] | 0.000<br>[0.000, 0.000] | 0.187<br>[0.105, 0.286] | – |
| Previous resection date reported | 0.786<br>[0.583, 1.000] | 0.667<br>[0.333, 1.000] | 1.000<br>[1.000, 1.000] | 0.571<br>[0.167, 1.000] | 0.800<br>[0.500, 1.000] | – |
| Previous resection date | – | – | – | – | – | 0.333<br>[0.000, 0.833] |
| Previous embolisation | 0.899<br>[0.814, 0.974] | 0.955<br>[0.842, 1.000] | 0.808<br>[0.643, 0.958] | 0.990<br>[0.966, 1.000] | 0.875<br>[0.756, 0.963] | – |
| Previous embolisation date reported | 0.650<br>[0.500, 0.812] | 0.682<br>[0.478, 0.857] | 1.000<br>[1.000, 1.000] | 0.300<br>[0.000, 0.625] | 0.811<br>[0.647, 0.923] | – |
| Previous embolisation date | – | – | – | – | – | 0.692<br>[0.385, 0.923] |
| Previous ablation | 0.961<br>[0.897, 1.000] | 0.889<br>[0.722, 1.000] | 0.941<br>[0.812, 1.000] | 0.982<br>[0.954, 1.000] | 0.914<br>[0.800, 1.000] | – |
| Previous ablation date reported | 0.929<br>[0.750, 1.000] | 0.889<br>[0.636, 1.000] | 1.000<br>[1.000, 1.000] | 0.857<br>[0.500, 1.000] | 0.941<br>[0.778, 1.000] | – |
| Previous ablation date | – | – | – | – | – | 0.500<br>[0.125, 0.875] |
| Previous transplant | 0.822<br>[0.765, 0.881] | 0.638<br>[0.528, 0.753] | 0.957<br>[0.891, 1.000] | 0.688<br>[0.580, 0.784] | 0.765<br>[0.679, 0.846] | – |

**Supplementary Table 10:**
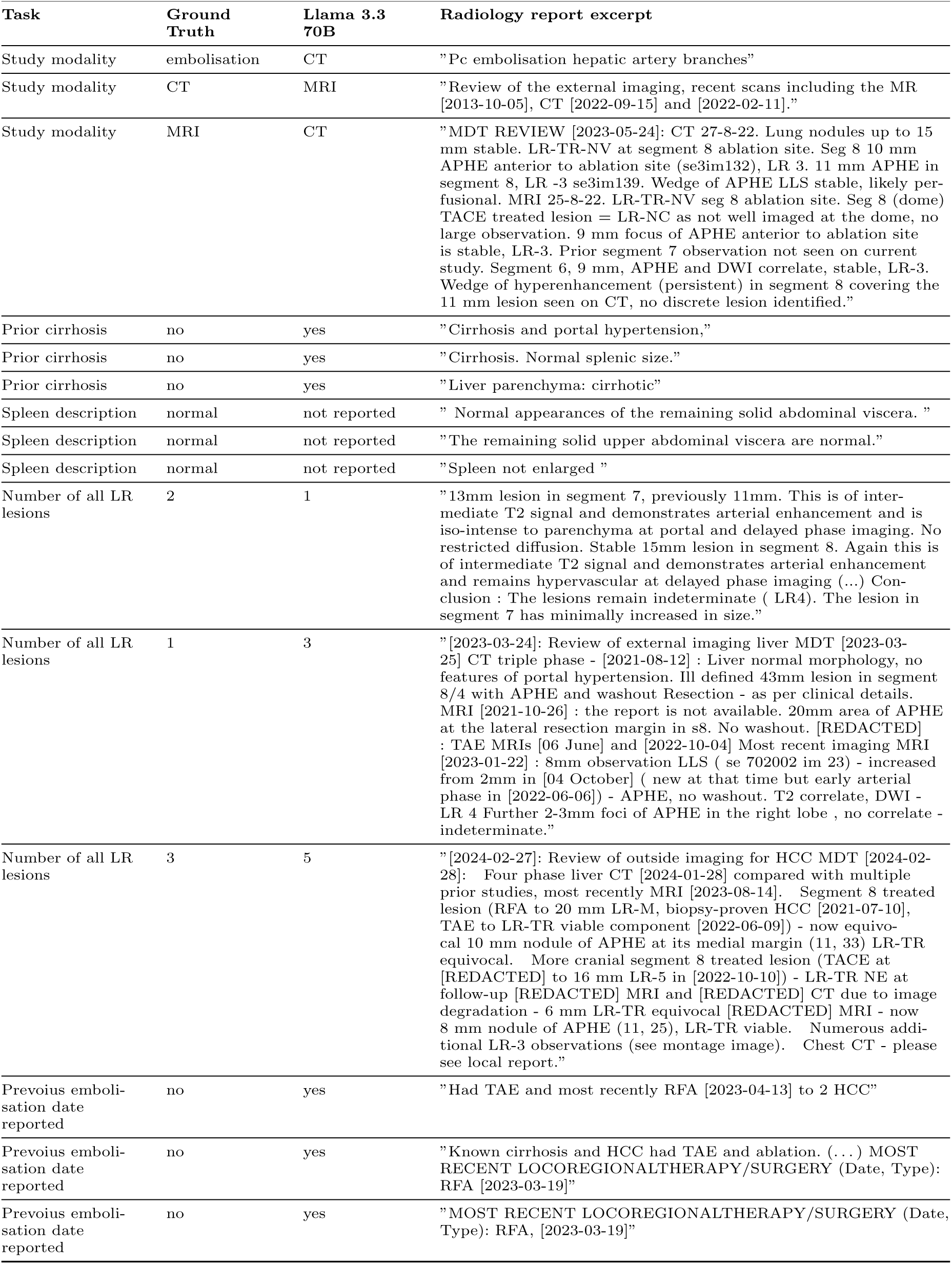
Error analysis of Llama 3.3 70B on radiology report extraction tasks. The table shows up to three examples from the worst-performing tasks (accuracy or proportion correct <90%), including relevant excerpts from radiology reports.

| Task | Ground Truth | Llama 3.3 70B | Radiology report excerpt |
| --- | --- | --- | --- |
| Study modality | embolisation | CT | "Pc embolisation hepatic artery branches" |
| Study modality | CT | MRI | "Review of the external imaging, recent scans including the MR [2013-10-05], CT [2022-09-15] and [2022-02-11]." |
| Study modality | MRI | CT | "MDT REVIEW [2023-05-24]: CT 27-8-22. Lung nodules up to 15 mm stable. LR-TR-NV at segment 8 ablation site. Seg 8 10 mm APHE anterior to ablation site (se3im132), LR 3. 11 mm APHE in segment 8, LR -3 se3im139. Wedge of APHE LLS stable, likely per-fusional. MRI 25-8-22. LR-TR-NV seg 8 ablation site. Seg 8 (dome) TACE treated lesion = LR-NC as not well imaged at the dome, no large observation. 9 mm focus of APHE anterior to ablation site is stable, LR-3. Prior segment 7 observation not seen on current study. Segment 6, 9 mm, APHE and DWI correlate, stable, LR-3. Wedge of hyperenhancement (persistent) in segment 8 covering the 11 mm lesion seen on CT, no discrete lesion identified." |
| Prior cirrhosis | no | yes | "Cirrhosis and portal hypertension," |
| Prior cirrhosis | no | yes | "Cirrhosis. Normal splenic size." |
| Prior cirrhosis | no | yes | "Liver parenchyma: cirrhotic" |
| Spleen description | normal | not reported | " Normal appearances of the remaining solid abdominal viscera. " |
| Spleen description | normal | not reported | "The remaining solid upper abdominal viscera are normal." |
| Spleen description | normal | not reported | "Spleen not enlarged " |
| Number of all LR lesions | 2 | 1 | "13mm lesion in segment 7, previously 11mm. This is of intermediate T2 signal and demonstrates arterial enhancement and is iso-intense to parenchyma at portal and delayed phase imaging. No restricted diffusion. Stable 15mm lesion in segment 8. Again this is of intermediate T2 signal and demonstrates arterial enhancement and remains hypervascular at delayed phase imaging (...) Conclusion : The lesions remain indeterminate ( LR4). The lesion in segment 7 has minimally increased in size." |
| Number of all LR lesions | 1 | 3 | "[2023-03-24]: Review of external imaging liver MDT [2023-03-25] CT triple phase - [2021-08-12] : Liver normal morphology, no features of portal hypertension. Ill defined 43mm lesion in segment 8/4 with APHE and washout Resection - as per clinical details. MRI [2021-10-26] : the report is not available. 20mm area of APHE at the lateral resection margin in s8. No washout. [REDACTED] : TAE MRIs [06 June] and [2022-10-04] Most recent imaging MRI [2023-01-22] : 8mm observation LLS ( se 702002 im 23) - increased from 2mm in [04 October] ( new at that time but early arterial phase in [2022-06-06]) - APHE, no washout. T2 correlate, DWI - LR 4 Further 2-3mm foci of APHE in the right lobe , no correlate - indeterminate." |
| Number of all LR lesions | 3 | 5 | "[2024-02-27]: Review of outside imaging for HCC MDT [2024-02-28]: Four phase liver CT [2024-01-28] compared with multiple prior studies, most recently MRI [2023-08-14]. Segment 8 treated lesion (RFA to 20 mm LR-M, biopsy-proven HCC [2021-07-10], TAE to LR-TR viable component [2022-06-09]) - now equivocal 10 mm nodule of APHE at its medial margin (11, 33) LR-TR equivocal. More cranial segment 8 treated lesion (TACE at [REDACTED] to 16 mm LR-5 in [2022-10-10]) - LR-TR NE at follow-up [REDACTED] MRI and [REDACTED] CT due to image degradation - 6 mm LR-TR equivocal [REDACTED] MRI - now 8 mm nodule of APHE (11, 25), LR-TR viable. Numerous additional LR-3 observations (see montage image). Chest CT - please see local report." |
| Prevoius embolisation date reported | no | yes | "Had TAE and most recently RFA [2023-04-13] to 2 HCC" |
| Prevoius embolisation date reported | no | yes | "Known cirrhosis and HCC had TAE and ablation. (...) MOST RECENT LOCOREGIONALTHERAPY/SURGERY (Date, Type): RFA [2023-03-19]" |
| Prevoius embolisation date reported | no | yes | "MOST RECENT LOCOREGIONALTHERAPY/SURGERY (Date, Type): RFA, [2023-03-19]" |

**Supplementary Table 11:** Pre-LIRADS radiology reports questions.

| Task | Possible answers |
| --- | --- |
| Are there any liver lesions which could develop into HCC in this report? | "yes", "no" |
| How many liver lesions which could develop into HCC are described in this report? | "0", "1", "2", "3", "4", "5", "6", "7", "8", "9" |
| What's the description of the liver lesion which could develop into HCC in this report? | "HCC", "probable HCC", "equivocal", "not reported" |

**Supplementary Table 12:** Performance metrics for pre-LIRADS questions - Llama 3.3 70B (Guidance).

| Task | Balanced accuracy | Precision | Sensitivity | Specificity | F1 score | Proportion correct |
| --- | --- | --- | --- | --- | --- | --- |
| Liver lesion present | 0.957<br>[0.874, 1.000] | 0.933<br>[0.769, 1.000] | 0.933<br>[0.769, 1.000] | 0.981<br>[0.940, 1.000] | 0.933<br>[0.815, 1.000] | – |
| Number of liver lesions | – | – | – | – | – | 0.941<br>[0.882, 0.985] |
| Liver lesion description | 0.875<br>[0.750, 1.000] | 0.942<br>[0.667, 1.000] | 0.933<br>[0.800, 1.000] | Not defined<br>(multi-class) | 0.924<br>[0.716, 1.000] | – |

**Supplementary Table 13:** Performance metrics for pre-LIRADS questions - Llama 3.1 8B (Guidance).

| Task | Balanced accuracy | Precision | Sensitivity | Specificity | F1 score | Proportion correct |
| --- | --- | --- | --- | --- | --- | --- |
| Liver lesion present | 0.914<br>[0.810, 0.991] | 0.867<br>[0.666, 1.000] | 0.867<br>[0.667, 1.000] | 0.962<br>[0.900, 1.000] | 0.867<br>[0.692, 0.973] | – |
| Number of liver lesions | – | – | – | – | – | 0.618<br>[0.500, 0.735] |
| Liver lesion description | 0.750<br>[0.667, 1.000] | 0.787<br>[0.571, 1.000] | 0.867<br>[0.667, 1.000] | Not defined<br>(multi-class) | 0.817<br>[0.593, 1.000] | – |

**Supplementary Table 14:**
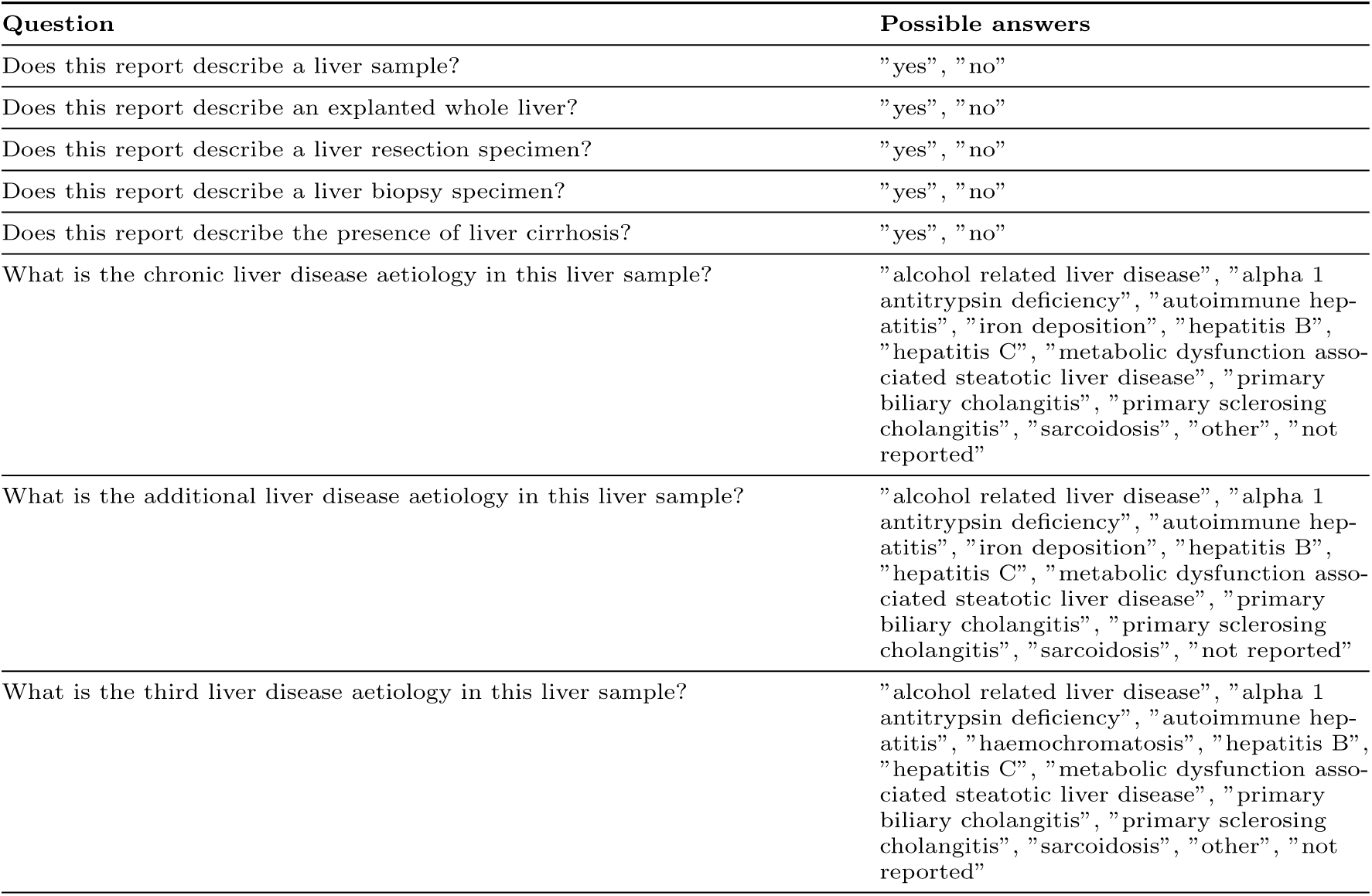
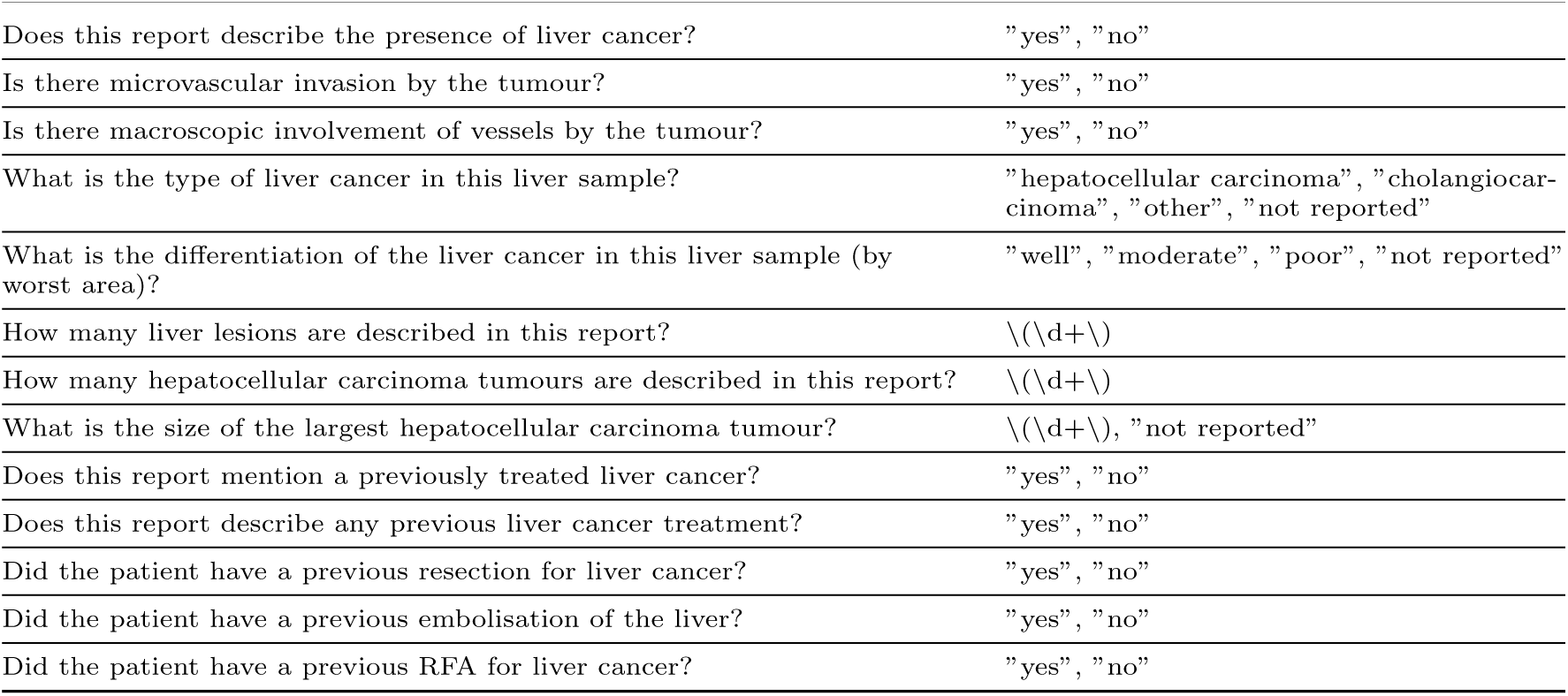
Pathology questions.

**Supplementary Table 15:**
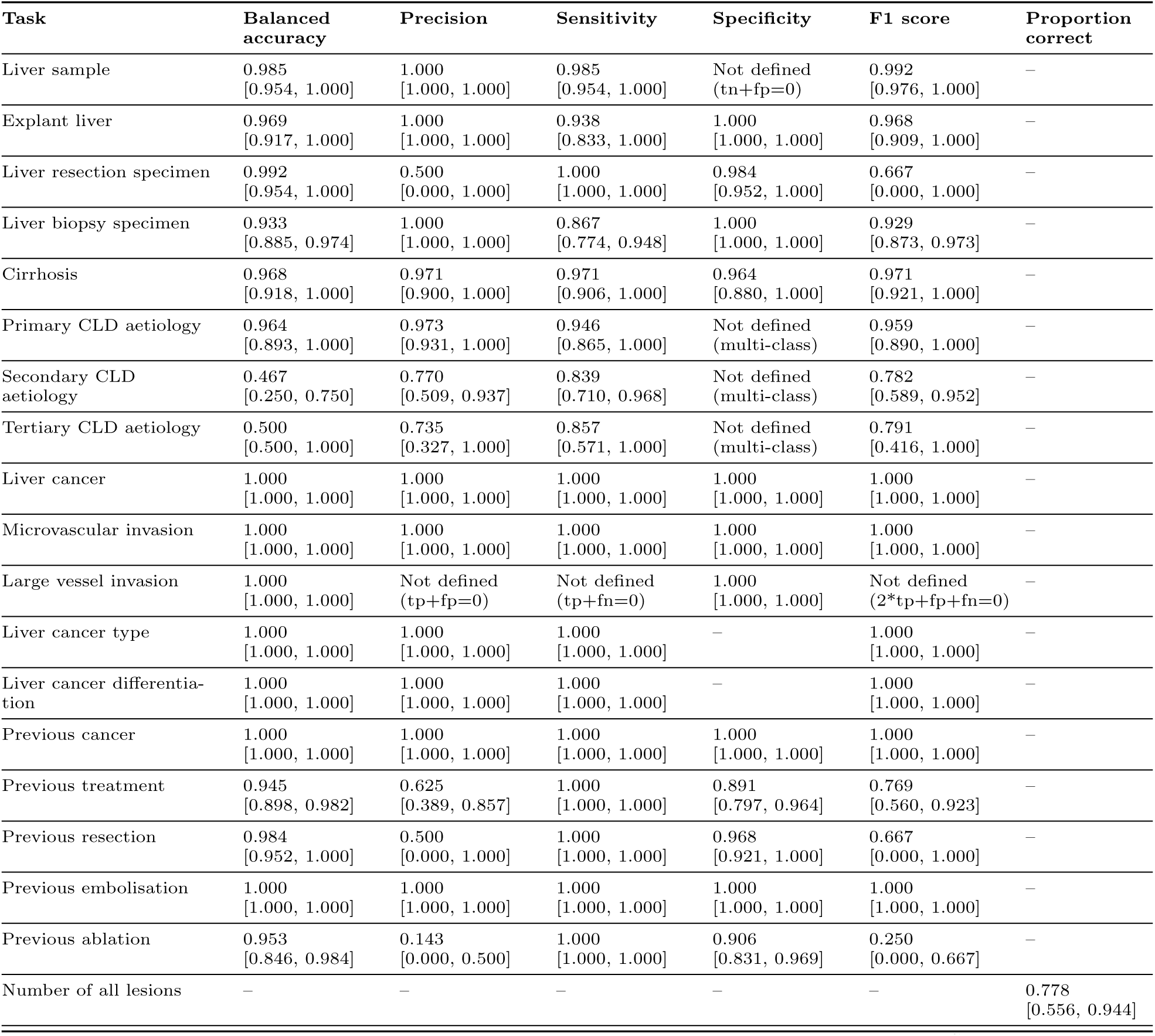
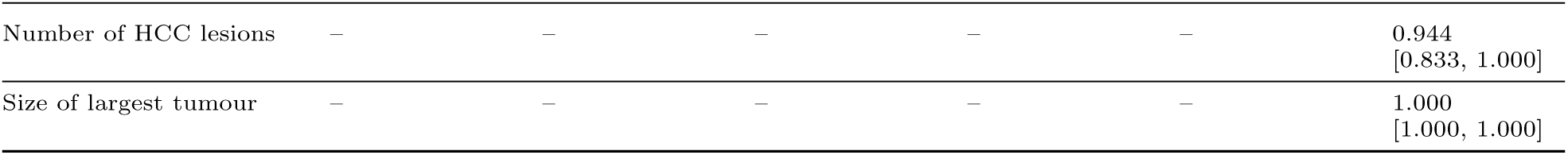
Performance metrics for pathology questions - Llama 3.3 70B (Guidance). (tp=true positive; tn=true negative; fp=false positive; fn=false negative).

**Supplementary Table 16:**
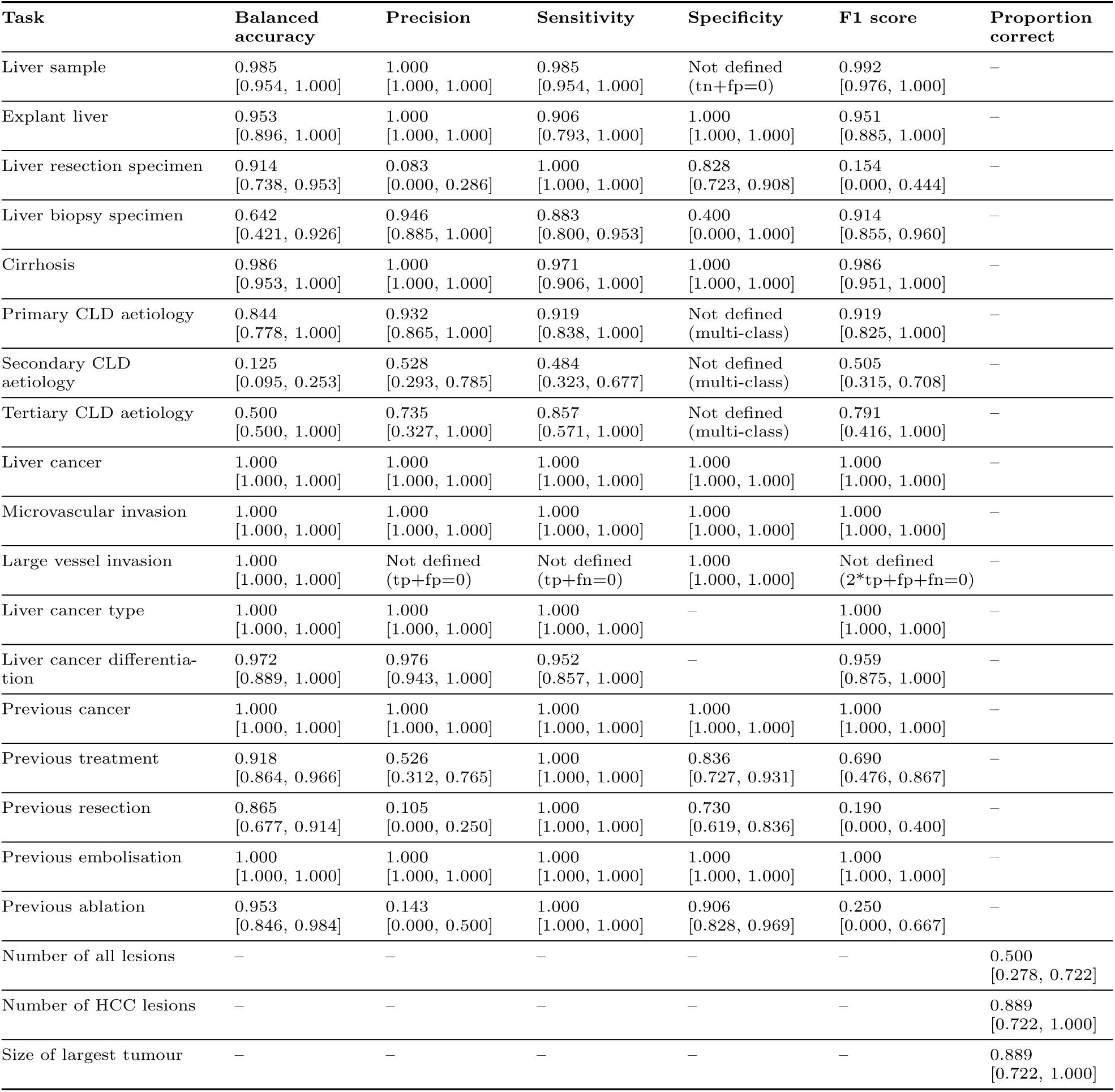
Performance metrics for pathology questions - Llama 3.1 8B (Guidance). (tp=true positive; tn=true negative; fp=false positive; fn=false negative).

| Task | Balanced accuracy | Precision | Sensitivity | Specificity | F1 score | Proportion correct |
| --- | --- | --- | --- | --- | --- | --- |
| Liver sample | 0.985<br>[0.954, 1.000] | 1.000<br>[1.000, 1.000] | 0.985<br>[0.954, 1.000] | Not defined<br>(tn+fp=0) | 0.992<br>[0.976, 1.000] | – |
| Explant liver | 0.953<br>[0.896, 1.000] | 1.000<br>[1.000, 1.000] | 0.906<br>[0.793, 1.000] | 1.000<br>[1.000, 1.000] | 0.951<br>[0.885, 1.000] | – |
| Liver resection specimen | 0.914<br>[0.738, 0.953] | 0.083<br>[0.000, 0.286] | 1.000<br>[1.000, 1.000] | 0.828<br>[0.723, 0.908] | 0.154<br>[0.000, 0.444] | – |
| Liver biopsy specimen | 0.642<br>[0.421, 0.926] | 0.946<br>[0.885, 1.000] | 0.883<br>[0.800, 0.953] | 0.400<br>[0.000, 1.000] | 0.914<br>[0.855, 0.960] | – |
| Cirrhosis | 0.986<br>[0.953, 1.000] | 1.000<br>[1.000, 1.000] | 0.971<br>[0.906, 1.000] | 1.000<br>[1.000, 1.000] | 0.986<br>[0.951, 1.000] | – |
| Primary CLD aetiology | 0.844<br>[0.778, 1.000] | 0.932<br>[0.865, 1.000] | 0.919<br>[0.838, 1.000] | Not defined<br>(multi-class) | 0.919<br>[0.825, 1.000] | – |
| Secondary CLD aetiology | 0.125<br>[0.095, 0.253] | 0.528<br>[0.293, 0.785] | 0.484<br>[0.323, 0.677] | Not defined<br>(multi-class) | 0.505<br>[0.315, 0.708] | – |
| Tertiary CLD aetiology | 0.500<br>[0.500, 1.000] | 0.735<br>[0.327, 1.000] | 0.857<br>[0.571, 1.000] | Not defined<br>(multi-class) | 0.791<br>[0.416, 1.000] | – |
| Liver cancer | 1.000<br>[1.000, 1.000] | 1.000<br>[1.000, 1.000] | 1.000<br>[1.000, 1.000] | 1.000<br>[1.000, 1.000] | 1.000<br>[1.000, 1.000] | – |
| Microvascular invasion | 1.000<br>[1.000, 1.000] | 1.000<br>[1.000, 1.000] | 1.000<br>[1.000, 1.000] | 1.000<br>[1.000, 1.000] | 1.000<br>[1.000, 1.000] | – |
| Large vessel invasion | 1.000<br>[1.000, 1.000] | Not defined<br>(tp+fp=0) | Not defined<br>(tp+fn=0) | 1.000<br>[1.000, 1.000] | Not defined<br>(2*tp+fp+fn=0) | – |
| Liver cancer type | 1.000<br>[1.000, 1.000] | 1.000<br>[1.000, 1.000] | 1.000<br>[1.000, 1.000] | – | 1.000<br>[1.000, 1.000] | – |
| Liver cancer differentiation | 0.972<br>[0.889, 1.000] | 0.976<br>[0.943, 1.000] | 0.952<br>[0.857, 1.000] | – | 0.959<br>[0.875, 1.000] | – |
| Previous cancer | 1.000<br>[1.000, 1.000] | 1.000<br>[1.000, 1.000] | 1.000<br>[1.000, 1.000] | 1.000<br>[1.000, 1.000] | 1.000<br>[1.000, 1.000] | – |
| Previous treatment | 0.918<br>[0.864, 0.966] | 0.526<br>[0.312, 0.765] | 1.000<br>[1.000, 1.000] | 0.836<br>[0.727, 0.931] | 0.690<br>[0.476, 0.867] | – |
| Previous resection | 0.865<br>[0.677, 0.914] | 0.105<br>[0.000, 0.250] | 1.000<br>[1.000, 1.000] | 0.730<br>[0.619, 0.836] | 0.190<br>[0.000, 0.400] | – |
| Previous embolisation | 1.000<br>[1.000, 1.000] | 1.000<br>[1.000, 1.000] | 1.000<br>[1.000, 1.000] | 1.000<br>[1.000, 1.000] | 1.000<br>[1.000, 1.000] | – |
| Previous ablation | 0.953<br>[0.846, 0.984] | 0.143<br>[0.000, 0.500] | 1.000<br>[1.000, 1.000] | 0.906<br>[0.828, 0.969] | 0.250<br>[0.000, 0.667] | – |
| Number of all lesions | – | – | – | – | – | 0.500<br>[0.278, 0.722] |
| Number of HCC lesions | – | – | – | – | – | 0.889<br>[0.722, 1.000] |
| Size of largest tumour | – | – | – | – | – | 0.889<br>[0.722, 1.000] |

**Supplementary Table 17:** Error analysis of Llama 3.3 70B on pathology report extraction tasks. The table shows up to three examples from the worst-performing tasks (accuracy or proportion correct <90%), including relevant excerpts from pathology reports.

| Task | Ground Truth | Llama 3.3 70B | Pathology report excerpt |
| --- | --- | --- | --- |
| Number of all lesions | 1 | 3 | "At the edge, there is fibroblastic proliferation and chronic inflammation and one small focus, approximately 2 mm of well differentiated hepatocellular carcinoma with pseudoglandular pattern and bile production. Less than 10% of cells are in cycle and there is no CK 19 expression. Vascular invasion is not seen. The smaller nodule is a macroregenerative nodule with prominent siderosis. A reactive lymph node without evidence of metastasis is identified at the hilum." |
| Number of all lesions | 2 | 3 | "Two atypical nodules are present as follows: slice 9 central mid right lobe 11mm pale yellow in colour and slice 12 posterior sub-capsular right lobe bile stained measuring 9mm. No further atypical nodules are identified." |
| Number of all lesions | 2 | 4 | "The two most atypical nodules are present in the caudate lobe where there is an intraparenchymal 9mm white lesion and in the mid medial left lobe, intraparenchymal lesion measuring 10mm with a soft green cut surface." |
| Secondary CLD aetiology | alpha 1 antitrypsin deficiency | not reported | "Establish micronodular cirrhosis with minimal steatosis and no ongoing steatohepatitis consistent with alcohol-related liver disease. Accumulation of alpha-1 antitrypsin material (PIZ to follow); no phenotype has been performed for this patient." |
| Secondary CLD aetiology | iron deposition | not reported | "background near inactive cirrhosis, in keeping with previous alcohol related injury although without ongoing steatohepatitis, and a grade 3 siderosis raising the possibility of an abnormality of iron metabolism." |
| Secondary CLD aetiology | alcohol related liver disease | not reported | "liver with ongoing steatohepatitic activity consistent with either non-alcoholic fatty liver disease or excess alcohol as the aetiology." |
| Tertiary CLD aetiology | alpha 1 antitrypsin deficiency | not reported | "Background cirrhosis with focal cholestasis and patchy, mild steatosis with minimally active steatohepatitis, consistent with a non-alcoholic fatty liver disease / alcohol aetiology; alpha-1 antitrypsin material is identified, in keeping with the known MZ phenotype; grade 2-3 hepatocellular siderosis also present. " |

**Supplementary Table 18:**
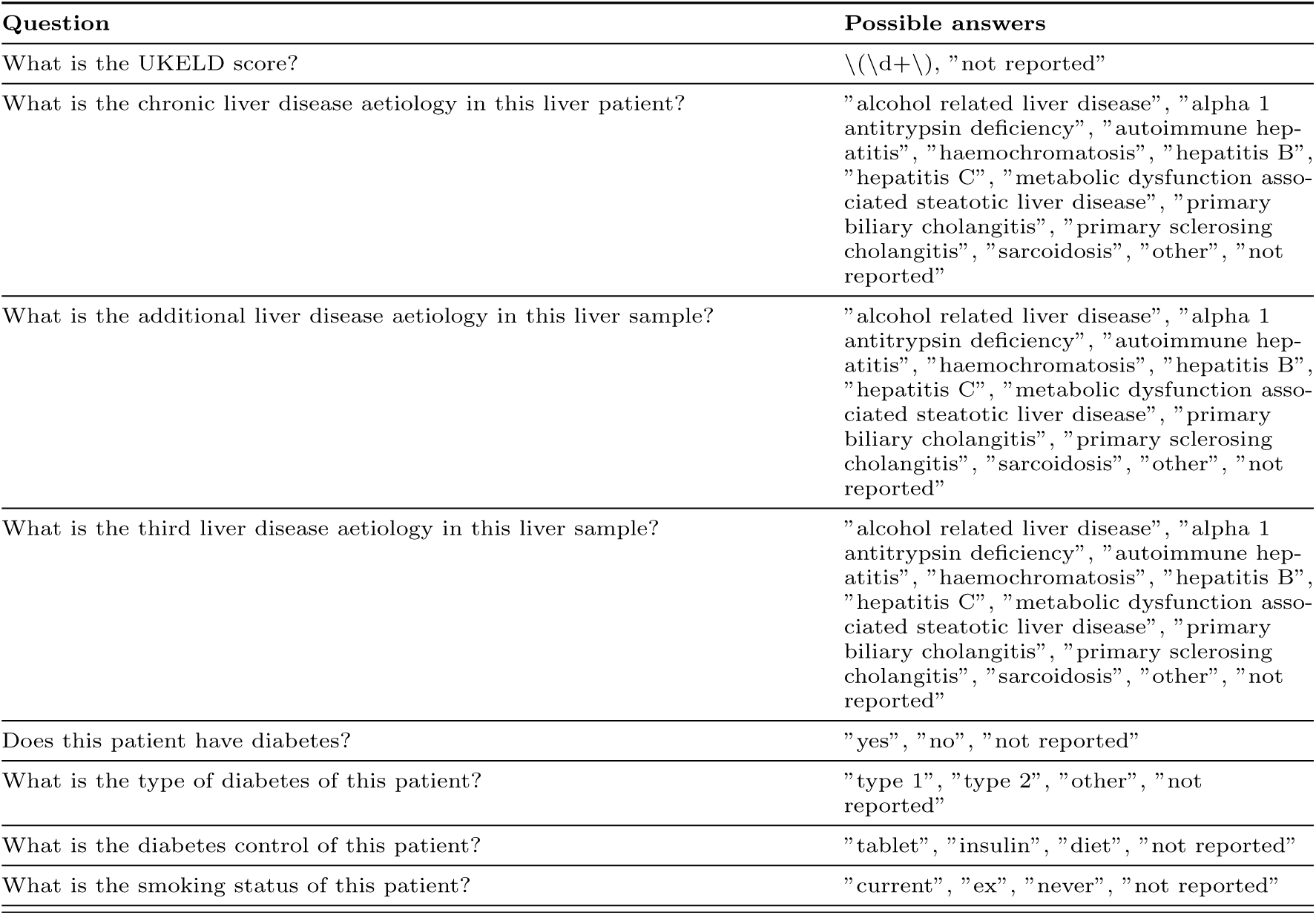
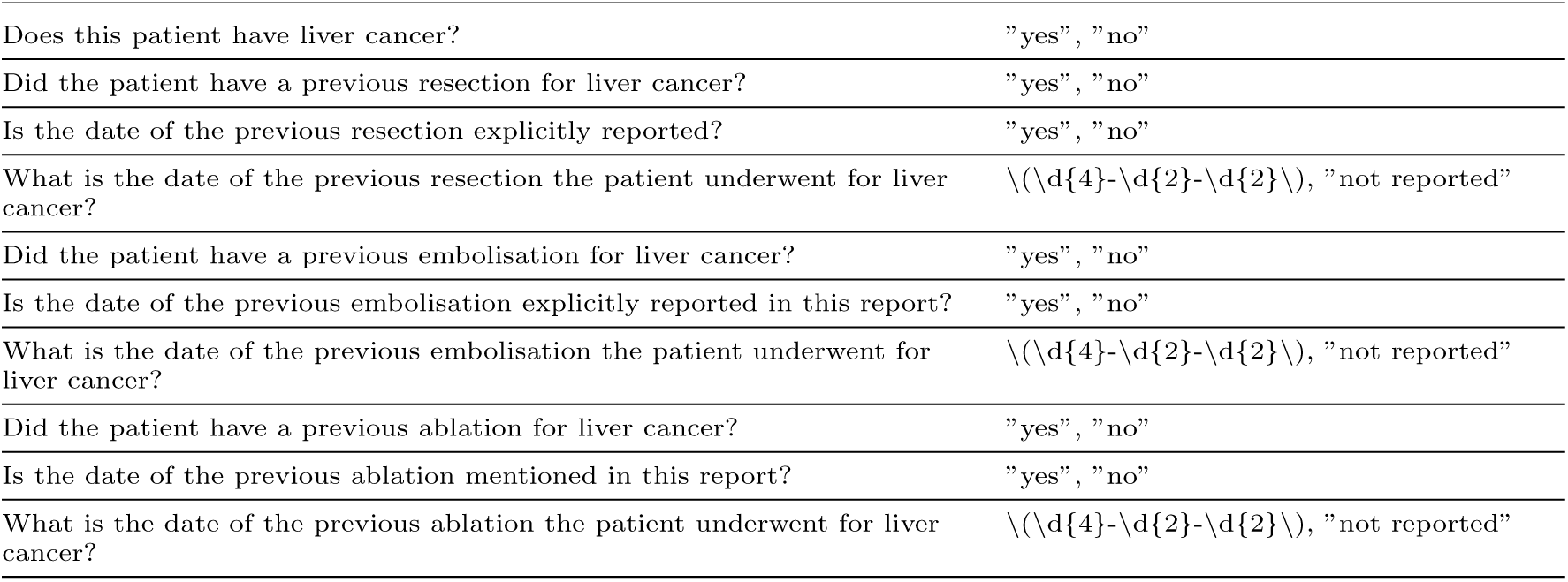
Liver transplant assessment questions.

**Supplementary Table 19:**
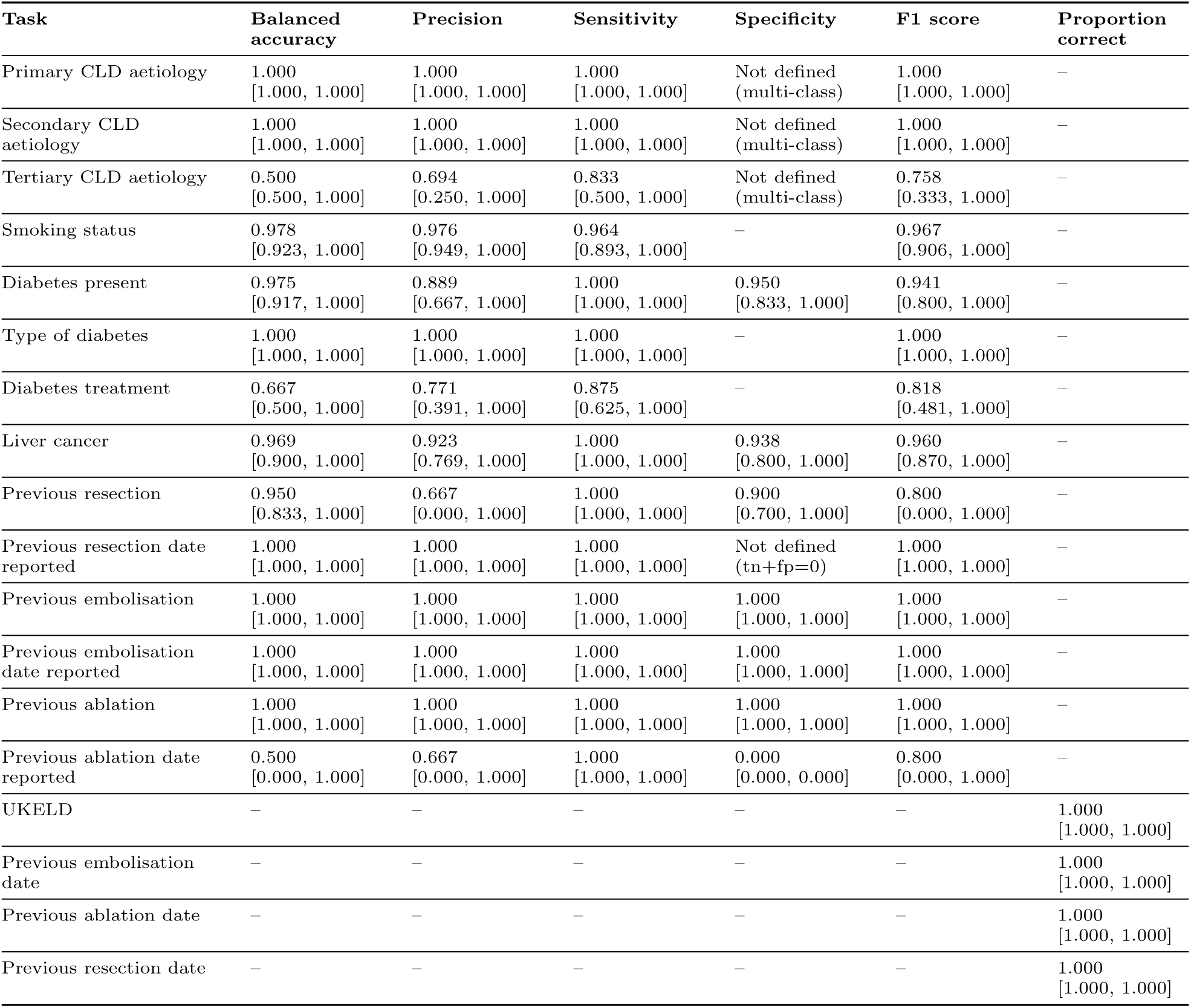
Performance metrics for LTA questions - Llama 3.3 70B (Guidance). (tn=true negative, fp=false positive).

| Task | Balanced accuracy | Precision | Sensitivity | Specificity | F1 score | Proportion correct |
| --- | --- | --- | --- | --- | --- | --- |
| Primary CLD aetiology | 1.000<br>[1.000, 1.000] | 1.000<br>[1.000, 1.000] | 1.000<br>[1.000, 1.000] | Not defined<br>(multi-class) | 1.000<br>[1.000, 1.000] | – |
| Secondary CLD aetiology | 1.000<br>[1.000, 1.000] | 1.000<br>[1.000, 1.000] | 1.000<br>[1.000, 1.000] | Not defined<br>(multi-class) | 1.000<br>[1.000, 1.000] | – |
| Tertiary CLD aetiology | 0.500<br>[0.500, 1.000] | 0.694<br>[0.250, 1.000] | 0.833<br>[0.500, 1.000] | Not defined<br>(multi-class) | 0.758<br>[0.333, 1.000] | – |
| Smoking status | 0.978<br>[0.923, 1.000] | 0.976<br>[0.949, 1.000] | 0.964<br>[0.893, 1.000] | – | 0.967<br>[0.906, 1.000] | – |
| Diabetes present | 0.975<br>[0.917, 1.000] | 0.889<br>[0.667, 1.000] | 1.000<br>[1.000, 1.000] | 0.950<br>[0.833, 1.000] | 0.941<br>[0.800, 1.000] | – |
| Type of diabetes | 1.000<br>[1.000, 1.000] | 1.000<br>[1.000, 1.000] | 1.000<br>[1.000, 1.000] | – | 1.000<br>[1.000, 1.000] | – |
| Diabetes treatment | 0.667<br>[0.500, 1.000] | 0.771<br>[0.391, 1.000] | 0.875<br>[0.625, 1.000] | – | 0.818<br>[0.481, 1.000] | – |
| Liver cancer | 0.969<br>[0.900, 1.000] | 0.923<br>[0.769, 1.000] | 1.000<br>[1.000, 1.000] | 0.938<br>[0.800, 1.000] | 0.960<br>[0.870, 1.000] | – |
| Previous resection | 0.950<br>[0.833, 1.000] | 0.667<br>[0.000, 1.000] | 1.000<br>[1.000, 1.000] | 0.900<br>[0.700, 1.000] | 0.800<br>[0.000, 1.000] | – |
| Previous resection date reported | 1.000<br>[1.000, 1.000] | 1.000<br>[1.000, 1.000] | 1.000<br>[1.000, 1.000] | Not defined<br>(tn+fp=0) | 1.000<br>[1.000, 1.000] | – |
| Previous embolisation | 1.000<br>[1.000, 1.000] | 1.000<br>[1.000, 1.000] | 1.000<br>[1.000, 1.000] | 1.000<br>[1.000, 1.000] | 1.000<br>[1.000, 1.000] | – |
| Previous embolisation date reported | 1.000<br>[1.000, 1.000] | 1.000<br>[1.000, 1.000] | 1.000<br>[1.000, 1.000] | 1.000<br>[1.000, 1.000] | 1.000<br>[1.000, 1.000] | – |
| Previous ablation | 1.000<br>[1.000, 1.000] | 1.000<br>[1.000, 1.000] | 1.000<br>[1.000, 1.000] | 1.000<br>[1.000, 1.000] | 1.000<br>[1.000, 1.000] | – |
| Previous ablation date reported | 0.500<br>[0.000, 1.000] | 0.667<br>[0.000, 1.000] | 1.000<br>[1.000, 1.000] | 0.000<br>[0.000, 0.000] | 0.800<br>[0.000, 1.000] | – |
| UKELD | – | – | – | – | – | 1.000<br>[1.000, 1.000] |
| Previous embolisation date | – | – | – | – | – | 1.000<br>[1.000, 1.000] |
| Previous ablation date | – | – | – | – | – | 1.000<br>[1.000, 1.000] |
| Previous resection date | – | – | – | – | – | 1.000<br>[1.000, 1.000] |

**Supplementary Table 20:** Performance metrics for LTA questions - Llama 3.1 8B (Guidance). (tn=true negative, tp=true positive, fp=false positive)

| Task | Balanced accuracy | Precision | Sensitivity | Specificity | F1 score | Proportion correct |
| --- | --- | --- | --- | --- | --- | --- |
| Primary CLD aetiology | 0.805<br>[0.689, 0.987] | 0.878<br>[0.760, 1.000] | 0.857<br>[0.714, 0.964] | Not defined<br>(multi-class) | 0.860<br>[0.712, 0.981] | – |
| Secondary CLD aetiology | 0.333<br>[0.333, 0.500] | 0.640<br>[0.413, 0.863] | 0.786<br>[0.607, 0.929] | Not defined<br>(multi-class) | 0.706<br>[0.491, 0.895] | – |
| Tertiary CLD aetiology | 0.500<br>[0.500, 1.000] | 0.694<br>[0.250, 1.000] | 0.833<br>[0.500, 1.000] | Not defined<br>(multi-class) | 0.758<br>[0.333, 1.000] | – |
| Smoking status | 0.933<br>[0.833, 1.000] | 0.916<br>[0.864, 1.000] | 0.893<br>[0.786, 1.000] | – | 0.893<br>[0.775, 1.000] | – |
| Diabetes present | 0.975<br>[0.917, 1.000] | 0.889<br>[0.667, 1.000] | 1.000<br>[1.000, 1.000] | 0.950<br>[0.833, 1.000] | 0.941<br>[0.800, 1.000] | – |
| Type of diabetes | 1.000<br>[1.000, 1.000] | 1.000<br>[1.000, 1.000] | 1.000<br>[1.000, 1.000] | – | 1.000<br>[1.000, 1.000] | – |
| Diabetes treatment | 0.667<br>[0.500, 1.000] | 0.771<br>[0.411, 1.000] | 0.875<br>[0.625, 1.000] | – | 0.818<br>[0.489, 1.000] | – |
| Liver cancer | 0.812<br>[0.687, 0.929] | 0.667<br>[0.450, 0.875] | 1.000<br>[1.000, 1.000] | 0.625<br>[0.375, 0.857] | 0.800<br>[0.621, 0.933] | – |
| Previous resection | 0.950<br>[0.833, 1.000] | 0.667<br>[0.000, 1.000] | 1.000<br>[1.000, 1.000] | 0.900<br>[0.700, 1.000] | 0.800<br>[0.000, 1.000] | – |
| Previous resection date reported | 1.000<br>[1.000, 1.000] | 1.000<br>[1.000, 1.000] | 1.000<br>[1.000, 1.000] | Not defined<br>(tn+fp=0) | 1.000<br>[1.000, 1.000] | – |
| Previous embolisation | 1.000<br>[1.000, 1.000] | 1.000<br>[1.000, 1.000] | 1.000<br>[1.000, 1.000] | 1.000<br>[1.000, 1.000] | 1.000<br>[1.000, 1.000] | – |
| Previous embolisation date reported | 0.500<br>[0.000, 0.500] | Not defined<br>(tp+fp=0) | 0.000<br>[0.000, 0.000] | 1.000<br>[1.000, 1.000] | 0.000<br>[0.000, 0.000] | – |
| Previous ablation | 0.889<br>[0.722, 1.000] | 0.600<br>[0.000, 1.000] | 1.000<br>[1.000, 1.000] | 0.778<br>[0.444, 1.000] | 0.750<br>[0.000, 1.000] | – |
| Previous ablation date reported | 1.000<br>[1.000, 1.000] | 1.000<br>[1.000, 1.000] | 1.000<br>[1.000, 1.000] | 1.000<br>[1.000, 1.000] | 1.000<br>[1.000, 1.000] | – |
| UKELD | – | – | – | – | – | 0.857<br>[0.714, 0.964] |
| Previous embolisation date | – | – | – | – | – | 0.833<br>[0.500, 1.000] |
| Previous ablation date | – | – | – | – | – | 1.000<br>[1.000, 1.000] |
| Previous resection date | – | – | – | – | – | 1.000<br>[1.000, 1.000] |

**Supplementary Table 21:** Error analysis of Llama 3.3 70B on LTA report extraction tasks. The table shows errors from the worst-performing tasks (accuracy or proportion correct <90%), including relevant excerpts from LTA reports.

| Task | Ground Truth | Llama 3.3 70B | LTA report excerpt |
| --- | --- | --- | --- |
| Diabetes treatment | diet | tablet | "Type 2 DBM - diagnosed [2024-07-09]. Previously on metformin but now diet controlled." and "T2DM - diagnosed 5 years ago. Previously on metformin. Stopped in [07 February]. Not on any medications at present." |
| Tertiary CLD aetiology | alcohol related liver disease | not reported | "Diagnosis: HCV cirrhosis/ NAFLD/ ETOH" |
| Previous ablation date reported | no | yes | "Subsequently found to have segment 6 lesion —> Ablation - Further LR3 lesion in segment 7/8 Most recent HCC MDT ([2022-10-24]) 30mm lesion Segment 5 (LR4) + 21mm lesion segment 5 (LR5) + 17mm lesion segment 7/8 (LR3) + 13mm lesion segment 7 (LR3) - Plan for TAE 21mm LR5 lesion + biopsy LR4 lesion +/- ablation but not visible on ultrasound scan so not performed Further TACE (4th) performed [24 November] for LR5 lesion" |

**Supplementary Table 22:**
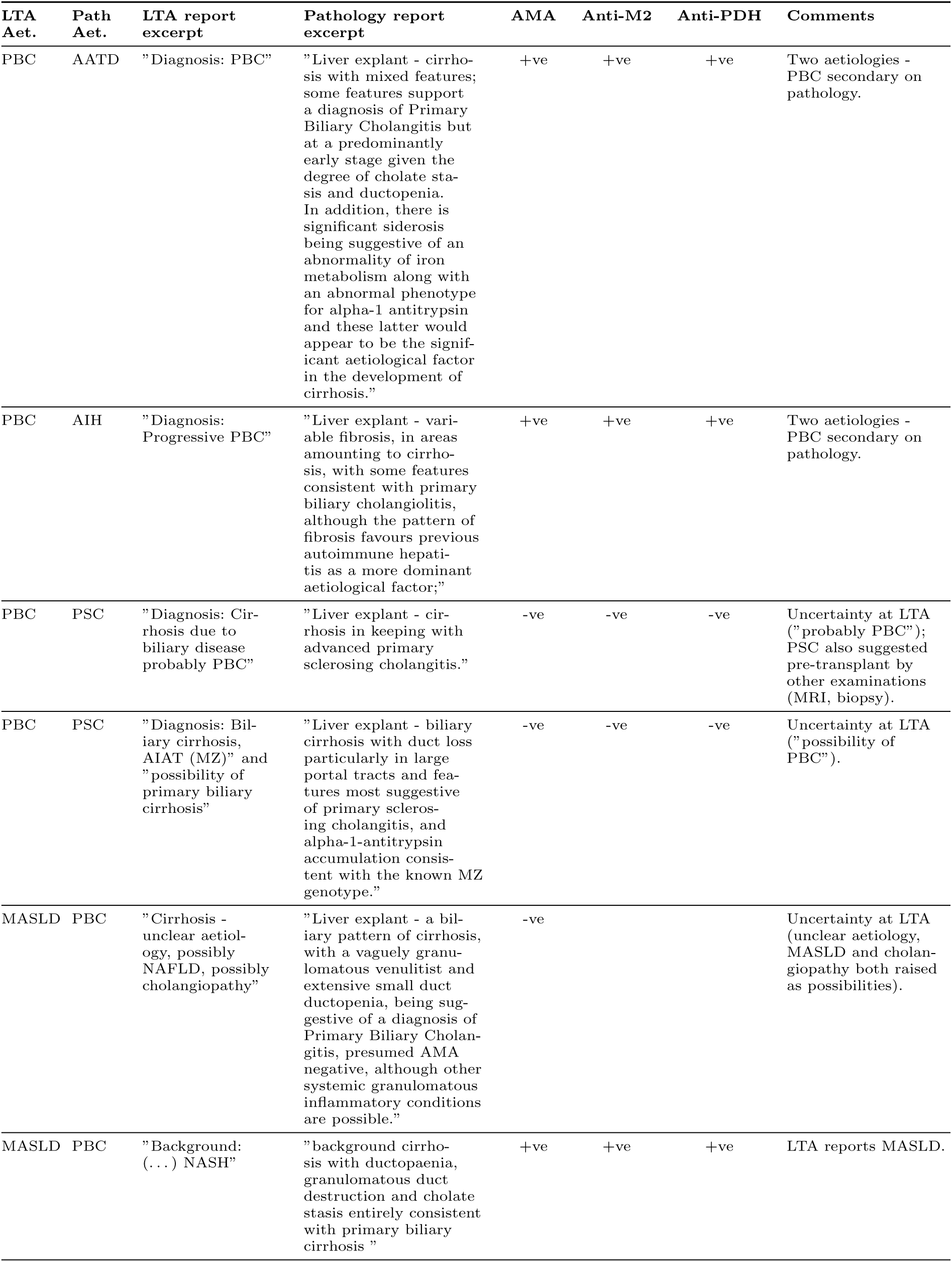
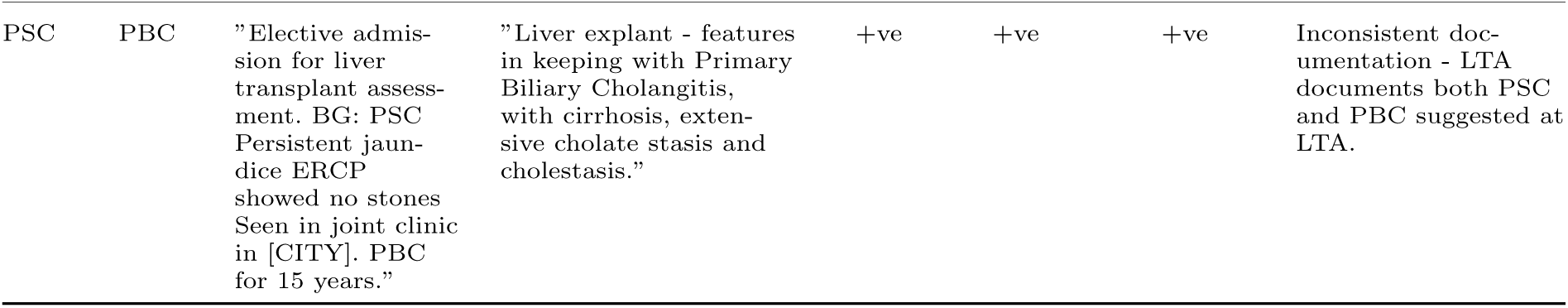
Examples of primary CLD aetiology re-classificaitons between LTA and pathology reports extracted by Llama 3.3 70B. Each row represents one patient. Results of serological tests were obtained by manual review of the patient records. Only relevant excerpt from the report is shown. [LTA aet. = Primary CLD aetiology extracted by Llama 3.3 70B from LTA reports; Path aet. = Primary CLD aetiology extracted by Llama 3.3 70B from pathology reports.]

**Supplementary Table 23:** Selected examples of errors in counting LR lesions for timelines reconstructed with Llama 3.3 70B. Only relevant excerpt from the reports is shown.

| Lesion type | Ground truth | LLM | Error type | Radiology report excerpt |
| --- | --- | --- | --- | --- |
| LR_3 | 2 | 1 | Missed lesion | "Segment 8 12 mm([REDACTED]) and 10 mm ([REDACTED]) LR3 observations with APHE but no washout ( [REDACTED]) appears stable allowing for differences in modality." |
| LR_TR_equivocal | 1 | 2 | Added lesion | "Segment 8 treated lesion (RFA to 20 mm LR-M, biopsy-proven HCC [2021-07-10], TAE to LR-TR viable component [2022-06-09]) - now equivocal 10 mm nodule of APHE at its medial margin (11, 33) LR-TR equivocal. More cranial segment 8 treated lesion (TACE at [REDACTED] to 16 mm LR-5 in [2022-10-10]) - LR-TR NE at follow-up [REDACTED] MRI and [REDACTED] CT due to image degradation - 6 mm LR-TR equivocal [REDACTED] MRI - now 8 mm nodule of APHE (11, 25), LR-TR viable." |
| LR_TR_nonviable | 1 | 2 | Added lesion | "MDT REVIEW [2023-05-24]: CT 27-8-22. Lung nodules up to 15 mm stable. LR-TR-NV at segment 8 ablation site. Seg 8 10 mm APHE anterior to ablation site (se3im132), LR 3. 11 mm APHE in segment 8, LR -3 se3im139. Wedge of APHE LLS stable, likely perfusional. MRI 25-8-22. LR-TR-NV seg 8 ablation site. Seg 8 (dome) TACE treated lesion = LR-NC as not well imaged at the dome, no large observation. 9 mm focus of APHE anterior to ablation site is stable, LR-3. Prior segment 7 observation not seen on current study. Segment 6, 9 mm, APHE and DWI correlate, stable, LR-3. Wedge of hyperenhancement (persistent) in segment 8 covering the 11 mm lesion seen on CT, no discrete lesion identified." |
| LR_4 | 0 | 1 | Added lesion | "Observation 2: LR-5 (previously LR-4) 16 mm S8 with APHE, washout and a capsule (s5 i47)." |
| LR_5 | 1 | 2 | Added lesion | "Focal lesions: Observation 1: Treated LR-5 observation. Good response to TAE. LR-TR equivocal (previously LR-TR viable). Equivocal nodule of arterial enhancement (s5 i86). No washout. (...). Observation 2: LR-5 (previously LR-4) 16 mm S8 with APHE, washout and a capsule (s5 i47). Increase in size from 14 mm. (...) Conclusion Cirrhosis and portal hypertension. LR-5 and LR-TR equivocal observations, both less than 3 cm." |
| LR_5 | 0 | 2 | Added lesion | "TREATED OBSERVATIONS: Observation #: 1 Treatment modality: TAE Location: Segment 7 Pretreatment category: LR-5 (...) Category: LR-TR Viable (...) Observation #: 2 Treatment modality: TAE Location: LMS Pretreatment category: LR-5 (...) LR-TR non-viable. ADDITIONAL OBSERVATIONS: Observation #: 3 Category: LR-3 - There are no further reportable Li-RADS observations (...) SUMMARY: - LR-TR viable segment 7 tumour, 15 mm enhancing component - LR-TR non-viable left medial section tumour - No evidence of metastatic disease within the abdomen or pelvis." |

**Supplementary Table 24:**
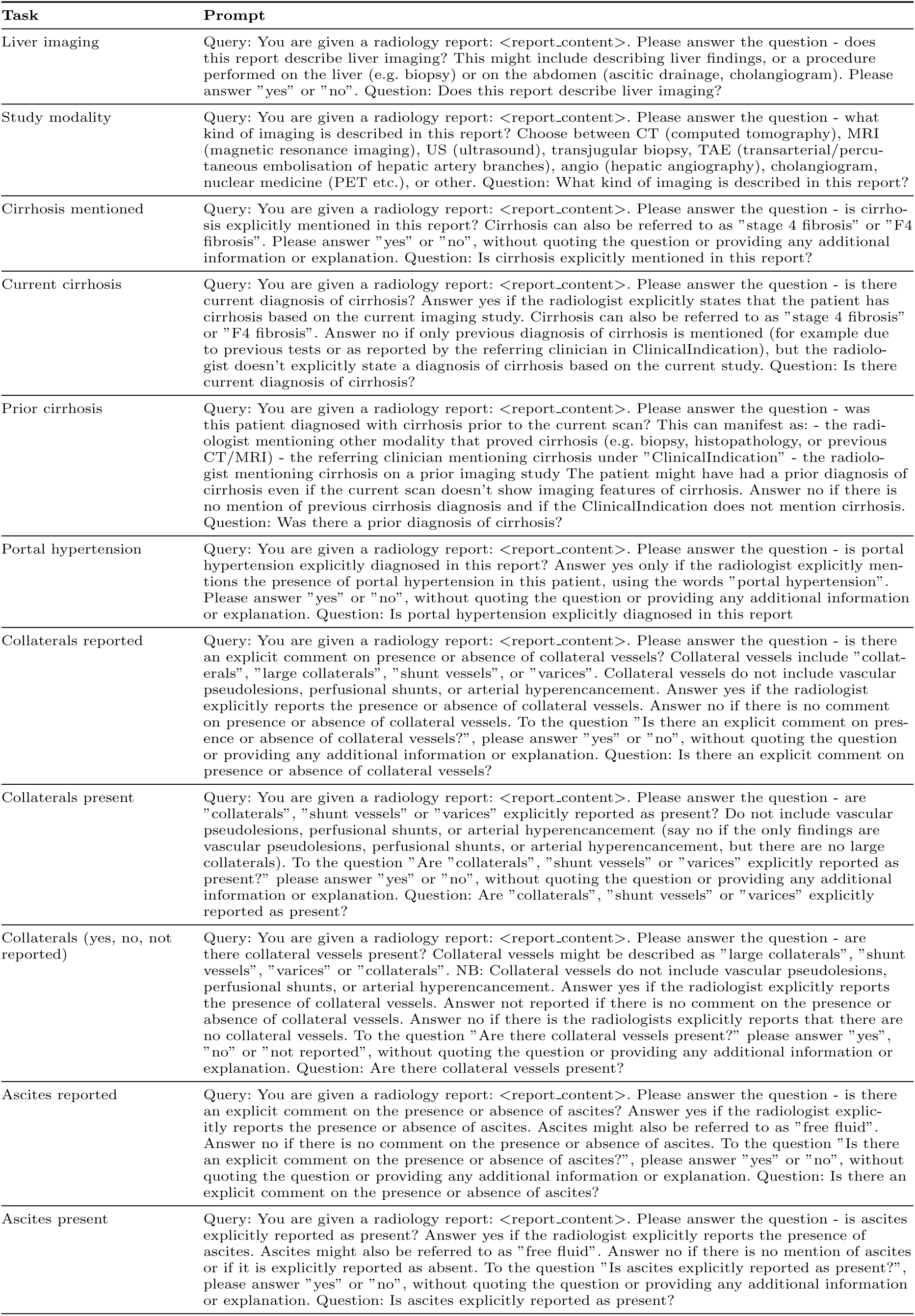
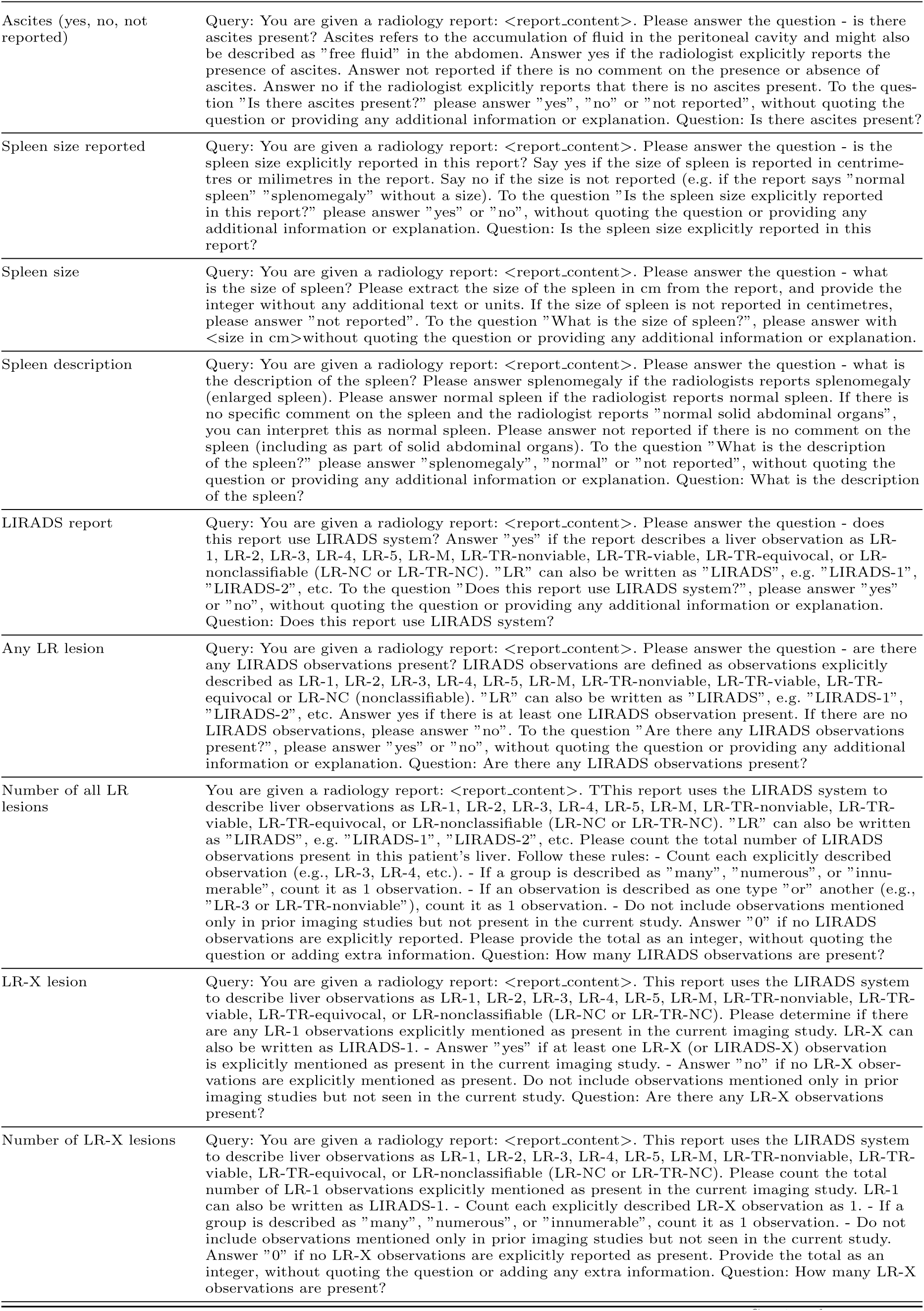
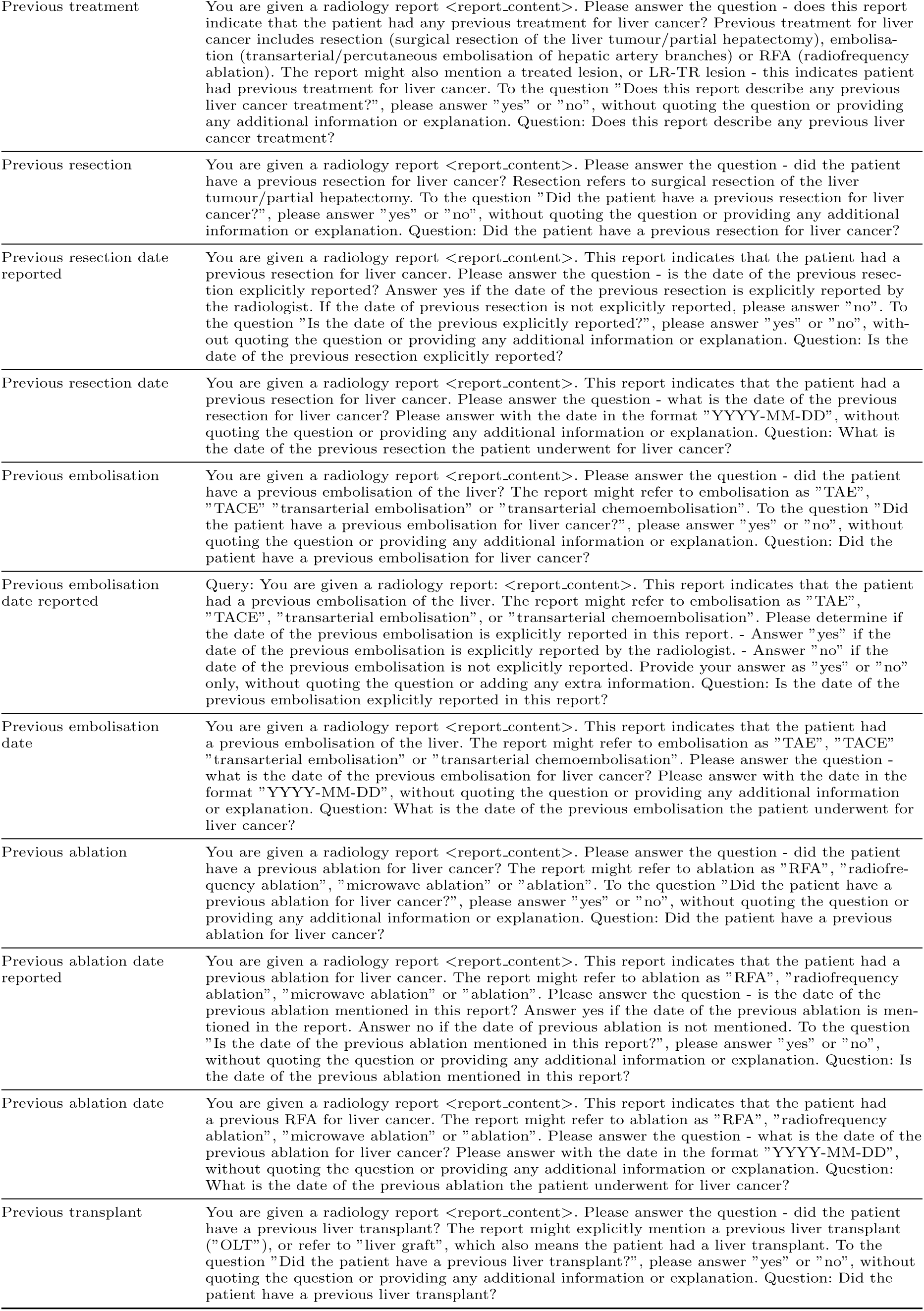
Radiology Prompts.

**Supplementary Table 25:** Pre-LIRADS radiology reports prompts.

| Task | Prompt |
| --- | --- |
| Liver lesion present | Query: You are given a radiology report: <report_content>. Your task is to identify whether there are any liver lesions which could develop into HCC (hepatocellular carcinoma) in this report. This might include liver lesions which are described as: - HCC - probable HCC, suspicious for HCC or similar - liver lesions which are equivocal, indeterminate, or needing further characterisation - previously treated HCC - liver lesions which are described as LIRADS 3, LIRADS 4, LIRADS 5, or similar Liver lesions which do not qualify include: - LIRADS 1 and LIRADS 2 lesions - cysts, haemangiomas, FNH, adenomas, or similar Based on this knowledge, please answer the question - are there any liver lesions which could develop into HCC in this report? Please answer "yes" or "no". Question: Are there any liver lesions which could develop into HCC in this report? |
| Number of liver lesions | Query: You are given a radiology report: <report_content>. Your task is to count the number of liver lesions which could develop into HCC (hepatocellular carcinoma) described in this report. This might include liver lesions which are described as: - HCC - probable HCC, suspicious for HCC or similar - liver lesions which are equivocal, indeterminate, or needing further characterisation - previously treated HCC - liver lesions which are described as LIRADS 3, LIRADS 4, LIRADS 5, or similar Liver lesions which do not qualify include: - LIRADS 1 and LIRADS 2 lesions - cysts, haemangiomas, FNH, adenomas, or similar Based on this knowledge, please answer the question - how many liver lesions which could develop into HCC are described in this report? Please answer with a positive integer. Question: How many liver lesions which could develop into HCC are described in this report? |
| Liver lesion description | Query: You are given a radiology report: <report_content>. Your task is to extract the description of liver lesions which could develop into HCC (hepatocellular carcinoma) described in this report. Here is a list of the descriptive categories and descriptions that fall into each category: - "HCC" - this includes descriptions such as "HCC", "definitive HCC" - "probable HCC" - this includes descriptions such as "probable HCC", "suspicious for HCC", "concerning for HCC" etc. - "equivocal" - this includes descriptions such as "indeterminate", "equivocal", "needs further characterisation" etc. - "not reported" - this category includes reports where no concrete description is provided by the radiologist (e.g. it states there is a lesion but with no further details) To the question "What's the description of the liver lesions which could develop into HCC in this report?", please answer with "HCC", "probable HCC", "equivocal", or "not reported". Question: What's the description of the liver lesions which could develop into HCC in this report? |

**Supplementary Table 26:**
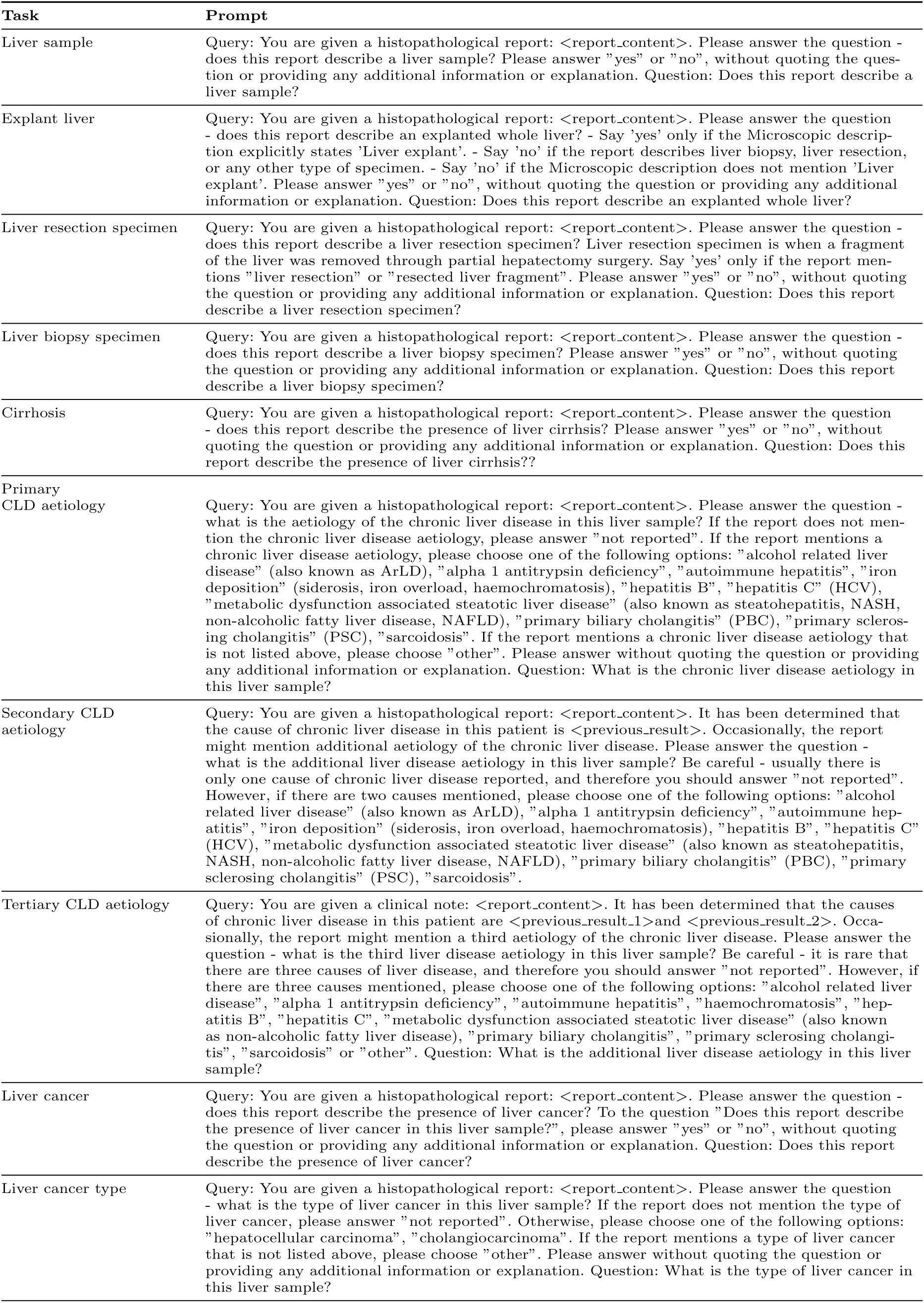
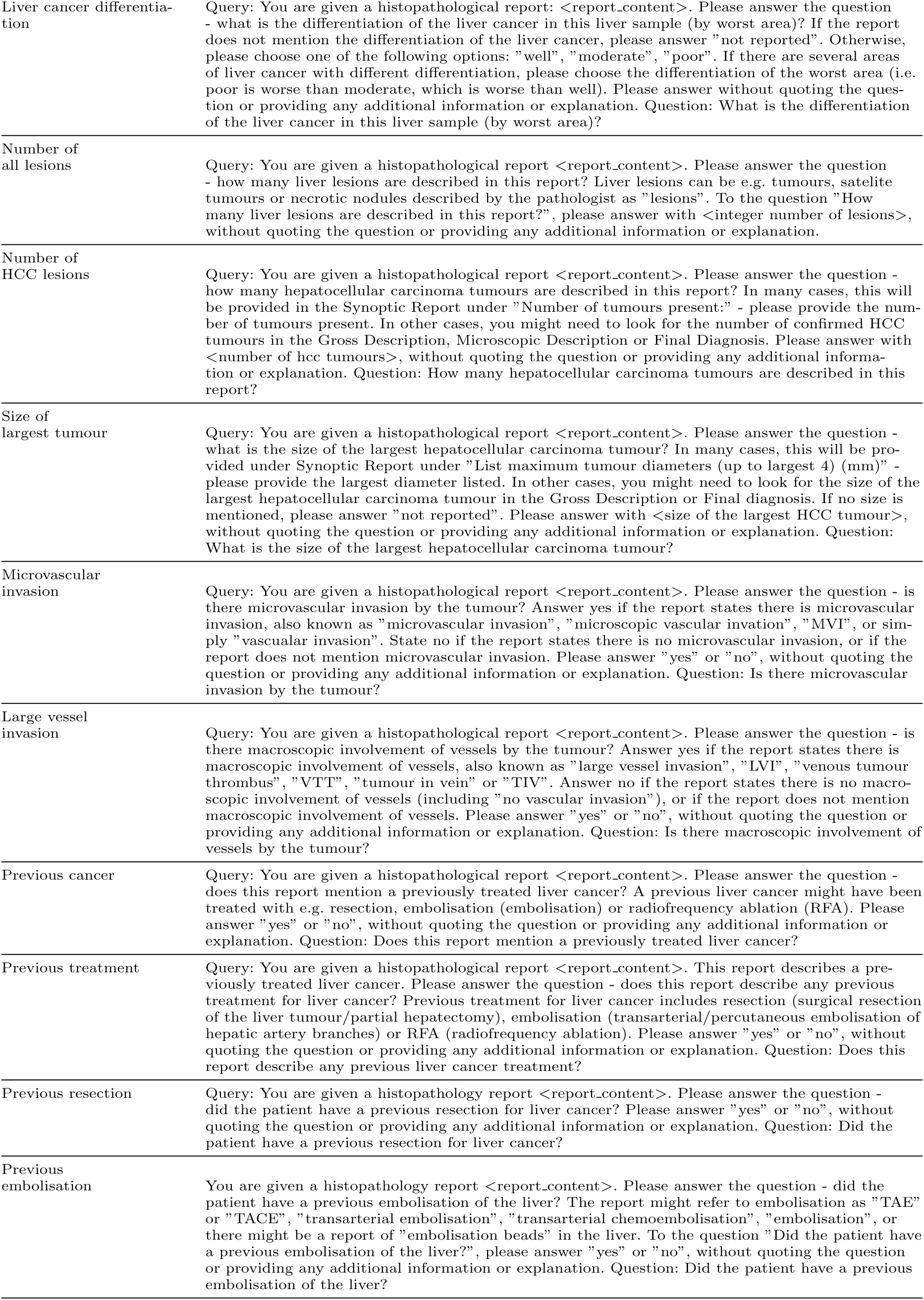
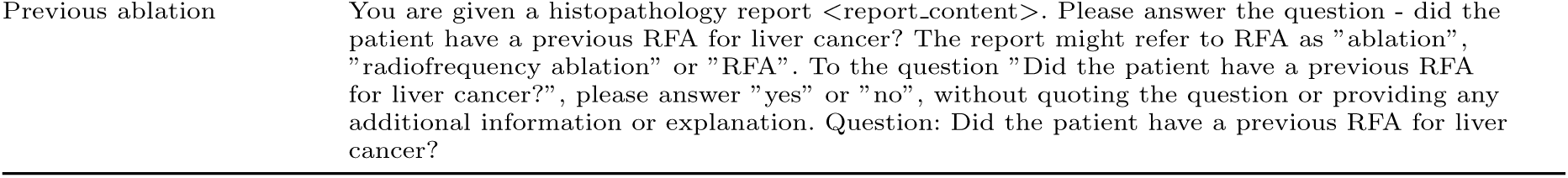
Pathology Prompts.

**Supplementary Table 27:**
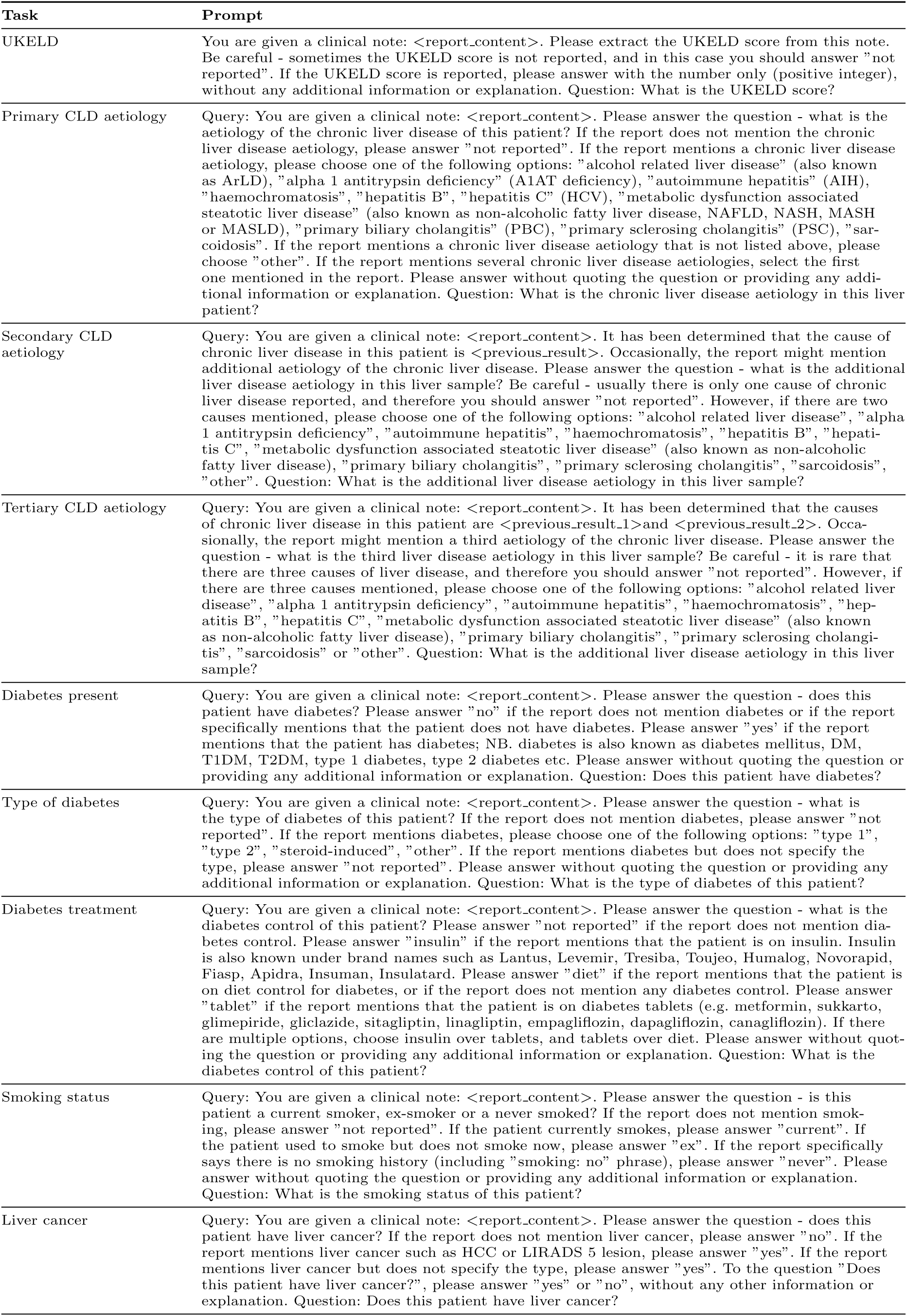
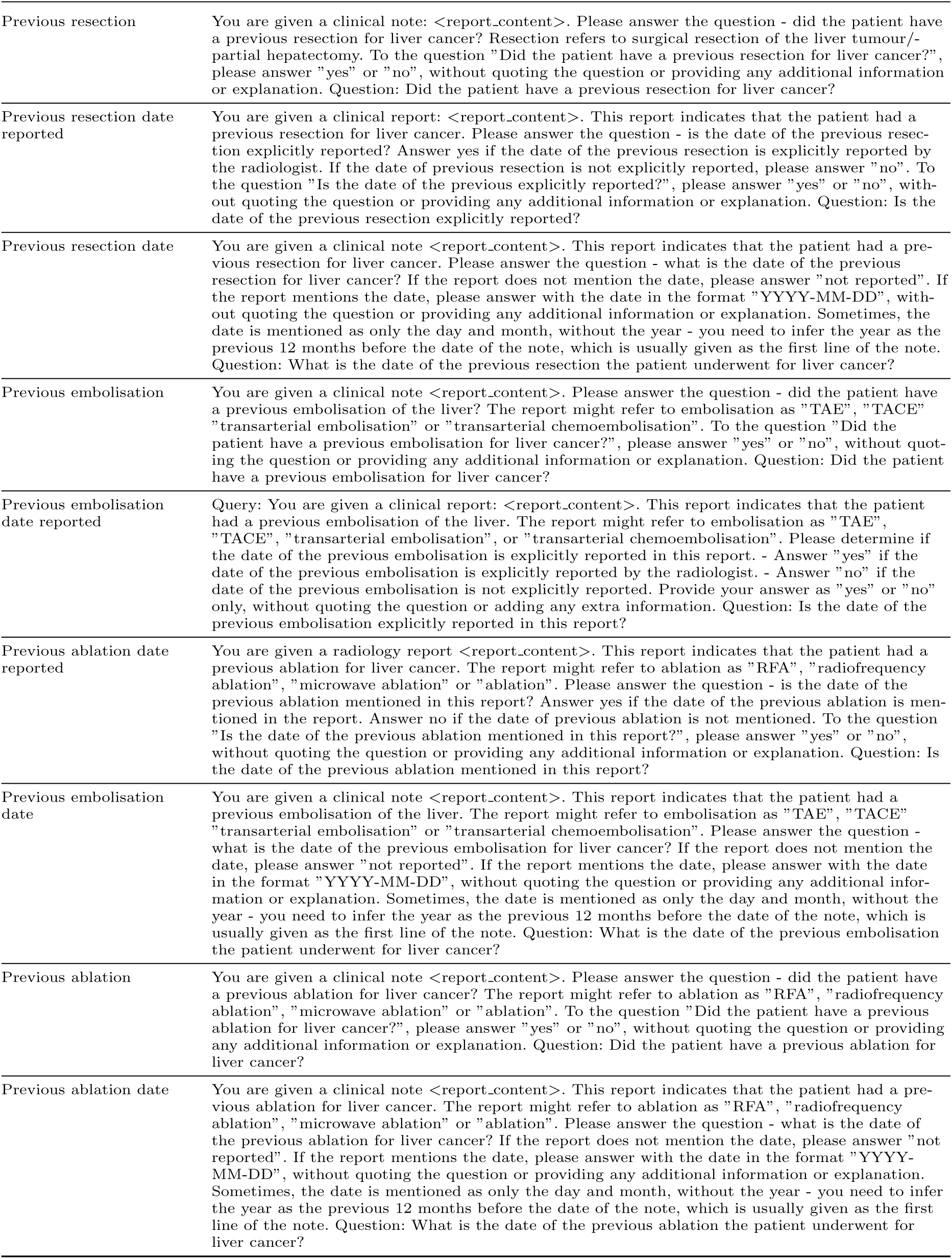
LTA Prompts.

